# Combinatorial clinically driven blood biomarker functional genomics significantly enhances genotype-phenotype resolution and diagnostics in neuromuscular disease

**DOI:** 10.1101/2021.01.14.21249850

**Authors:** Samya Chakravorty, Kiera Berger, Laura Rufibach, Logan Gloster, Sarah Emmons, Sreekala Shenoy, Madhuri Hegde, Ashok Reddy Dinasarapu, Greg Gibson

## Abstract

**Purpose:** 50-60% of neuromuscular-disease patients remain undiagnosed even after extensive genetic testing that hinders precision-medicine/clinical-trial-enrollment. Importantly, those with DNA-based molecular diagnosis often remain without known molecular mechanism driving different degrees of disease severity that hinders patient stratification and trial-readiness. These are due to: a) clinical-genetic-heterogeneity (eg: limb-girdle-muscular-dystrophies(LGMDs)>30-subtypes); b) high-prevalence of variants-of-uncertain-significance (VUSs); (c) unresolved genotype-phenotype-correlations for patient stratification, and (d) lack of minimally-invasive biomarker-driven-assays. We therefore implemented a combinatorial phenotype-driven blood-biomarker functional-genomics approach to enhance diagnostics and trial-readiness by elucidating disease mechanisms of a neuromuscular-disease patient-cohort clinically-suspected of Dysferlinopathy/related-LGMD, the second-most-prevalent LGMD in the US.

**Methods:** We used CD14+monocyte protein-expression-assay on 364 Dysferlinopathy/related-LGMD-suspected patient-cohort without complete molecular-diagnosis or genotype-phenotype correlation; and then combined with blood-based targeted-transcriptome-sequencing (RNA-Seq) with tiered-analytical-algorithm correlating with clinical-measurements for a subset of patients.

**Results:** Our combinatorial-approach significantly increased the diagnostic-yield from 25% (N=326; 18%-27%; 95%CI) to 82% (N=38; 69.08% to 84.92%; 95% CI) by combining monocyte-assay with enhanced-RNA-Seq-analysis and clinical-correlation, following ACMG-AMP-guidelines. The tiered-analytical-approach detected aberrant-splicing, allele-expression-imbalance, nonsense-mediated-decay, and compound-heterozygosity without parental/offspring-DNA-testing, leading to VUS-reclassifications, identification of variant-pathomechanisms, and enhanced genotype-phenotype resolution including those with carrier-range Dysferlin-protein-expression and milder-symptoms, allowing patient-stratification for better trial-readiness. We identified uniform-distribution of pathogenic-variants across *DYSF*-gene-domains without any hotspot suggesting the relevance of upcoming gene-(full-*DYSF*-cDNA)-therapy trials.

**Conclusion:** Our results show the relevance of using a clinically-driven multi-tiered-approach utilizing a minimally-invasive biomarker-functional-genomic platform for precision-medicine-diagnostics, trial-recruitment/monitoring, elucidating pathogenic-mechanisms for patient stratification to enhance better trial outcomes, which in turn, will guide more rational use of current-therapeutics and development of novel-interventions for neuromuscular-disorders, and applicable to other genetic-disorders.

## INTRODUCTION

Limb-girdle muscular-dystrophies (LGMDs) are one of the most prevalent and heterogeneous inherited neuromuscular-disorders (NMDs) with >30 monogenic clinically-overlapping subtypes[1]. Among them, Dysferlinopathy (MIM 254130, 253601, 606768), a recessively-inherited muscular dystrophy caused by variants in the *DYSF* (MIM 603009)[2, 3] gene, with variability in clinical presentations[4-6] is the second most prevalent LGMD[1, 7, 8]. Definitive molecular-diagnosis is typically a pre-requirement to enroll patients with such clinico-genetic heterogeneity into clinical-trials. Recently, in a large LGMD 35 gene-panel next-generation-sequencing (NGS) program, we achieved 27% (1259/4656 patients) diagnostic-yield. However, 72% of all clinically-reportable variants were variants of uncertain significance (VUSs) resulting in ∼50% of cohort, including at least 90 patients with *DYSF* VUSs or unresolved compound heterozygosity without known phasing, remaining undiagnosed hindering trial-enrolment[1].Importantly, with upcoming trials of gene-therapy (NCT02710500: https://clinicaltrials.gov/ct2/show/NCT02710500?term=NCT02710500&rank=1) and others on the horizon, improved understanding of genotype-phenotype correlations by identifying mechanism of pathogenicity of not only VUSs, but also pathogenic or likely pathogenic variants at the molecular level is essential to stratify patients appropriately for better readiness to clinical trials or precision medicine initiatives.

Rigorous VUS-reclassification and resolution of genotype-combinations per American College of Medical Genetics and Genomics (ACMG) guidelines[9] requires understanding disease mechanisms using an integrative approach combining functional-assays with phenotype-correlation[10-12]. Gene-based or other biomarker testing from easily-accessible tissue (eg: blood/urine) is needed since muscle-biopsies or skin-derived-transdifferentiated myotubes are invasive, costly, and adipocyte contamination can compromise quality. DYSF protein-expression in blood, although shown by us to be an effective Dysferlinopathy biomarker[13-16], was not able to provide definitive genotype-phenotype correlations, resolve carrier-range detection for patients clearly Dysferlinopathy-suspected, and was unable to reclassify VUSs. Alternatively, transcriptome-sequencing (RNA-Seq) using patient muscles or myotubes or fibroblasts or blood without in-depth focused clinical-correlation increased diagnostic-yield to a maximum of 36% in clinically-diverse cohorts [17-20]. We show here that blood-based targeted-RNA-Seq with clinical-correlation does have high resolution when the candidate gene, such as *DYSF*, is adequately expressed as suggested but not shown by two other groups [17, 18]. Recently, though blood-based whole transcriptome RNA-Seq on trios (proband and parents) provided 38% diagnostic yield in combination with exome or genome sequencing[21], for adult neuromuscular disorders that are late-onset such as Dysferlinopathy and many other neuromuscular disorders, parental DNA/RNA may not be available for segregation studies. Hence, using NMD-specific targeted higher depth blood-based RNA-Seq, we show here resolution of phases of previously unresolved compound heterozygous variants using allele-expression imbalance (AEI) as was suggested previously[22].

Using a phenotype-driven combinatorial blood-biomarker approach (Fig. 1), we show here that diagnostic-yield increase to 82% (31/38) can be attained drawing from a large cohort of 364 patients clinically-suspected of Dysferlinopathy or related-LGMD without complete molecular diagnosis. Our results illustrate the importance of phenotype-driven biomarker-based functional genomics to improve understanding of variant-gene-disease relationships.

**Fig. 1.**
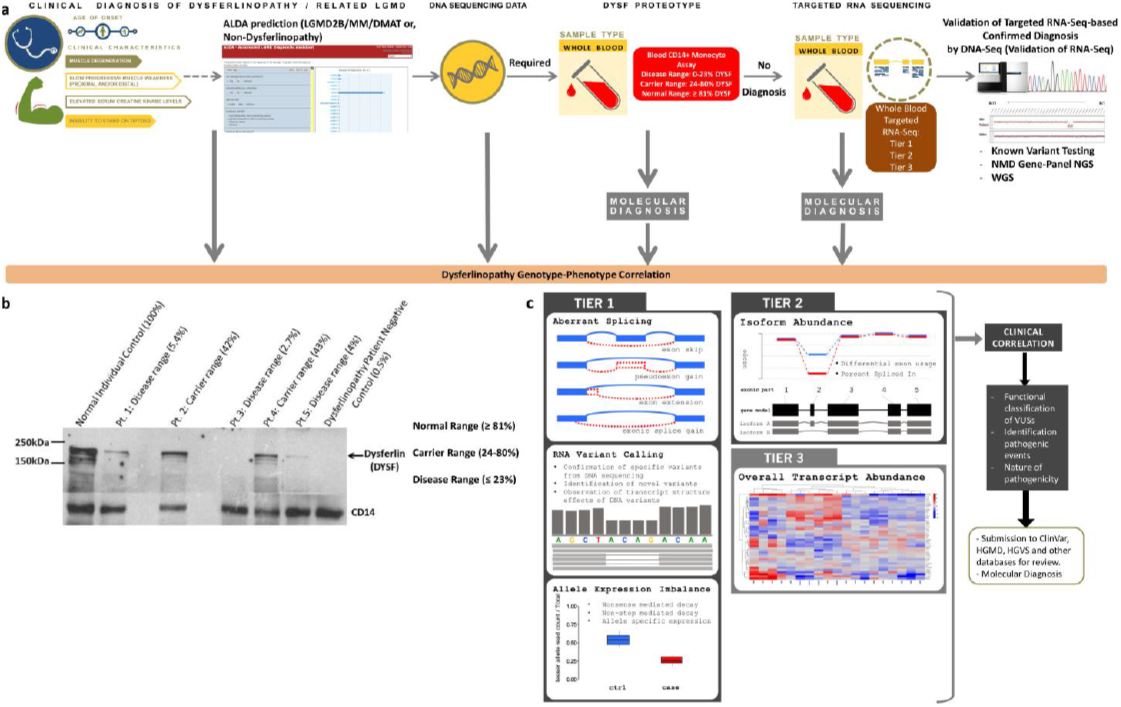
Study design and methods pipeline. (**a**) Study design and methods pipeline where patients with a clinical suspicion of Dysferlinopathy [limb-girdle muscular dystrophy 2B (LGM2B) or miyoshi myopathy (MM) or distal myopathy with anterior tibial onset (DMAT)] were recruited in the study for definitive molecular diagnosis with analysis of genotype-phenotype correlation. Then, the automated LGMD diagnostic assistant (ALDA) prediction was performed on cases for which there was sufficient clinical data. Any available prior CLIA-certified genetic testing (NGS panel, DYSF Sanger testing, exome, or array CGH based on respective physician discretion) report was collected. Functional assays of blood based monocyte assay and targeted RNA-Seq was used to definitively diagnose the patients. To validate results of RNA-Seq, subsequent DNA-Seq [known variant testing, neuromuscular disease (NMD) panel, CNV analysis, or genome sequencing (GS)] was performed. (**b**) Example of CD14+ monocyte assay immunoblot showing stratification of patient samples (unaffected normal ≥81%, carrier-range 24%-80%, and disease-range ≤23%) based on DYSF protein expression (with CD14 as loading and housekeeping gene control, ∼55kDa) in blood. (**c**) The tiered approach in targeted RNA-Seq analysis for molecular diagnostics and reclassification of VUSs. Tier 1 focused on the analysis of aberrant splicing, variant calls and allele expression imbalance identification; Tier 2 analyzed isoform pattern and exon usage, and Tier 3 evaluated gene expression analysis of the *DYSF* gene first followed by the remaining genes (if required and relevant) in the targeted neuromuscular disease (NMD) panel.

## MATERIALS AND METHODS

### Study design

A total of 364 patients of diverse ethnicities (including Americas and Europe) with clinical-suspicion of Dysferlinopathy (LGMD2B/Miyoshi Myopathy/Distal Myopathy with Anterior Tibial Onset/related LGMD) or related-LGMD and 15 normal-control samples from unaffected individuals were recruited between 2016 and 2019 in this study at Emory University based on the following inclusion- and exclusion-criteria. Inclusion-criteria: Patients with Dysferlinopathy or related LGMD clinical-suspicion were selected after comprehensive clinical-evaluation. Exclusion-criteria: Patients with definitive clinical-diagnosis of other unrelated muscular-dystrophy types were excluded in order to target a focused Dysferlinopathy-suspected cohort. Written informed-consents were obtained from all participants of the study according to Institutional Review Board approval. A combinatorial approach of a) focused clinical-data, b) a clinical-grade-assay for DYSF protein-quantification in CD14+ monocytes (Fig. 1A and 1B), and c) a clinically-driven whole-blood targeted-NMD-associated 274 gene-panel (Table S1) RNA-Seq (Fig. 1A and 1C) was used. We used a tiered (Tiers1-3) RNA-Seq-analysis (Fig. 1C) to identify novel pathogenic-variants, reclassify VUSs, and elucidate disease-pathomechanisms by evaluating splicing, allelic/gene-expression, and mapped the *DYSF* variant-landscape. Subsequent DNA-testing using genome sequencing (GS) or neuromuscular-disease gene-panel-testing with deletion-duplication-analysis, or sanger-testing was performed to confirm the pathogenic-variants identified (Fig. 1). For details of study design including sample-size, data-exclusions, replication, randomization, blinding, clinical-evaluation, patient-recruitment including patient-enrollment-questionnaires, see Supplementary Materials and Methods.

### Combinatorial approach of ALDA-prediction, genotype, functional assays

The Jain Foundation had previously developed an algorithm to predict LGMD-subtypes based on patient clinical-features known as the Automated LGMD Diagnostic Assistant (ALDA: https://www.jain-foundation.org/lgmd-subtyping-diagnosis-tool)[1, 16]. We used clinical-data that was available from patients recruited in this study to run ALDA-prediction (Table S2) of their LGMD/NMD-subtype. Genotype information from prior CLIA-CAP-certified genetic-testing reports for all 364 patients who underwent DNA-testing to identify disease genetic-basis was also collected where available. These genetic tests were heterogeneous, ranging from exome or array-comparative-genomics-hybridization to known variant Sanger-sequencing based on respective physician’s discretion. Subsequently, 326 out of 364 patients underwent blood CD14+ monocyte-assay (MA-assay) for minimally-invasive DYSF protein-profile-analysis using immunoblotting. ALDA-prediction was used for clinical-correlation of MA-assay results. Thereafter, targeted RNA-Seq using whole blood with a tiered-analytical-approach was performed on a subset of 51 consenting patients either without complete molecular-diagnosis (38 patients) or without resolution of genotype-phenotype correlation even with confirmed genetic diagnosis (13 patients). This combinatorial sequential approach enabled understanding the Dysferlinopathy genotype-phenotype landscape (Fig. 1). For details of a) MA-assay protocol, b) whole-blood targeted-RNA-Seq including library-preparation, sequencing, tiered-analysis-approach, alignment and quality-control, variant-calling, analysis of splicing, allele-expression imbalance, gene-expression; c) GS analysis; d) neuromuscular-disease 131 gene-panel list and sequencing with deletion-duplication-analysis; and e) sanger-testing, see Supplementary Materials and Methods. Custom R-Scripts and Codes used for RNA-Seq data-analysis are made publicly available through the Github repository: https://github.com/kiera-gt/rnaseq-nmd.

### Patient outcome: enrolment in Dysferlin Registry

All patients of our cohort who received confirmed molecular-diagnosis of Dysferlinopathy and proper stratification based on genotype-phenotype-correlation for better trial-readiness from this study were invited and included with informed-consent to the Dysferlin Registry (https://dysferlinregistry.jain-foundation.org/), an international global patient-registry. The Dysferlin Registry includes an interactive format that provides forums and educational information to the Dysferlinopathy-confirmed patients participating in the registry. The inclusion of Dysferlinopathy patients to the registry enhances the trial-readiness of the community and aids in the recruitment for the International Clinical Outcome Study for Dysferlinopathy (COS; NCT01676077:https://clinicaltrials.gov/ct2/show/NCT01676077?term=NCT01676077&rank=1) the upcoming follow-up COS2 study (https://www.jain-foundation.org/dysferlinoutcomestudy), as well as promising therapeutic clinical-trials. The Dysferlin Registry also provides de-identified data to researchers after obtaining the appropriate approval.

### Statistical analysis

Details of statistical-analysis for all comparisons are provided in places of respective data in main manuscript, figure legends, and Supplementary Information.

## Results

### CD14+ monocyte assay improves patient-stratification

The MA-assay was able to stratify patients as either normal-range (≥81%DYSF), carrier-range (24-80%DYSF), or disease-range (≤23%DYSF) based on their DYSF protein-expression (%DYSF) (Fig. 1B). Among the 219 disease-range cases, 76% (163/219) had two *DYSF* pathogenic variants (Table S3), whereas among 64 carrier-range and 59 normal-range cases, only 8% (5/64) and 2% (1/59) respectively had two *DYSF* pathogenic variants indicating the MA-assay’s robustness and that Dysferlin protein absence is highly suggestive of Dysferlinopathy. Eight carrier-range %DYSF patients were genetically confirmed of a different muscular-dystrophy (*FKRP*(MIM 606596)/*MYOT*(MIM 604103)/*CAPN3*(MIM 114240)/*ANO5*(MIM 608662)), and 25 mostly carrier-range %DYSF multi-genic patients with variants in both *DYSF* and ≥1 other gene(s) (Table S2 and S4) were identified, suggesting a possible genetic-interaction or secondary-effect on %DYSF caused by other muscular-dystrophies that needs to be considered, similar to previous reports[23]. Altogether, %DYSF as a blood-biomarker provides a robust tool for diagnosing Dysferlinopathy, and helps determine those for which additional analysis, like RNA-Seq, is needed.

### Combinatorial biomarker approach enhances genotype-phenotype resolution and molecular diagnostics

Our combinatorial-biomarker-approach aims to not only significantly enhance molecular-diagnostics but importantly also elucidate *DYSF* variant pathogenic-mechanisms at both RNA and protein levels so that genotype-phenotype correlation can be elucidated for better trial-readiness. Definitive molecular diagnosis was determined by a positive-correlation between results of each available confirmed data category used (Table S2) i.e., for example diagnosis will be achieved only if there two pathogenic *DYSF* variants and a disease-range %DYSF protein. Diagnostic-yield was 47% (40%-53%; 95% CI; N=348) based on correlation between confirmed genotype (two *DYSF* pathogenic-variants) only, but decreased to 25% (18%-27%; 95% CI; N=326) based on correlation between confirmed genotype and %DYSF, suggesting that %DYSF although can be a first-pass clinical biomarker is not sufficient enough to resolve both diagnostics and genotype-phenotype correlation.

We reasoned that whole-blood targeted-RNA-Seq of 274 NMD-associated genes expressed in skeletal muscle (Table S1)[24] with high read-depth in a tiered-analysis (Tiers1/2/3; Fig. 1C), in combination with genotype, %DYSF and clinical-data could enhance diagnostics by identifying splice-defects (PVS1/PS3-criteria), confirming phasing of variants (*cis/trans*;PM3-criteria), and uncovering novel-mechanisms, per ACMG-guidelines[9]. The Genotype-Tissue Expression (GTEx: https://gtexportal.org/home/) shows *DYSF*-overexpression in whole-blood. *DYSF* isoform-expression differences between blood and muscle are unlikely to hinder analysis since alternate exons are in-frame and well-expressed in blood except for exon 17 (Table 3 of [25]). 51 Dysferlinopathy-suspected patients who provided consents underwent RNA-Seq, among whom 38 patients’ %DYSF and/or DNA-sequencing were unable to provide a complete molecular diagnosis and in 13 patients the pathomechanism of the identified *DYSF*-variants were unclear. High average coverage of >500X for *DYSF* mRNA allowed for high-confidence analyses. Combining targeted RNA-Seq with genotype, ALDA, and %DYSF resulted in the molecular diagnosis of 31 out of the 38 previously undiagnosed and incomplete cases significantly increasing the diagnostic-yield to 82% (N=38; 69.08% to 84.92%; 95% CI) (Fig. 2A), and resolved the genotype-phenotype correlation and nature of pathogenicity of variant combinations in 13 patients (Table S5).

**Fig. 2.**
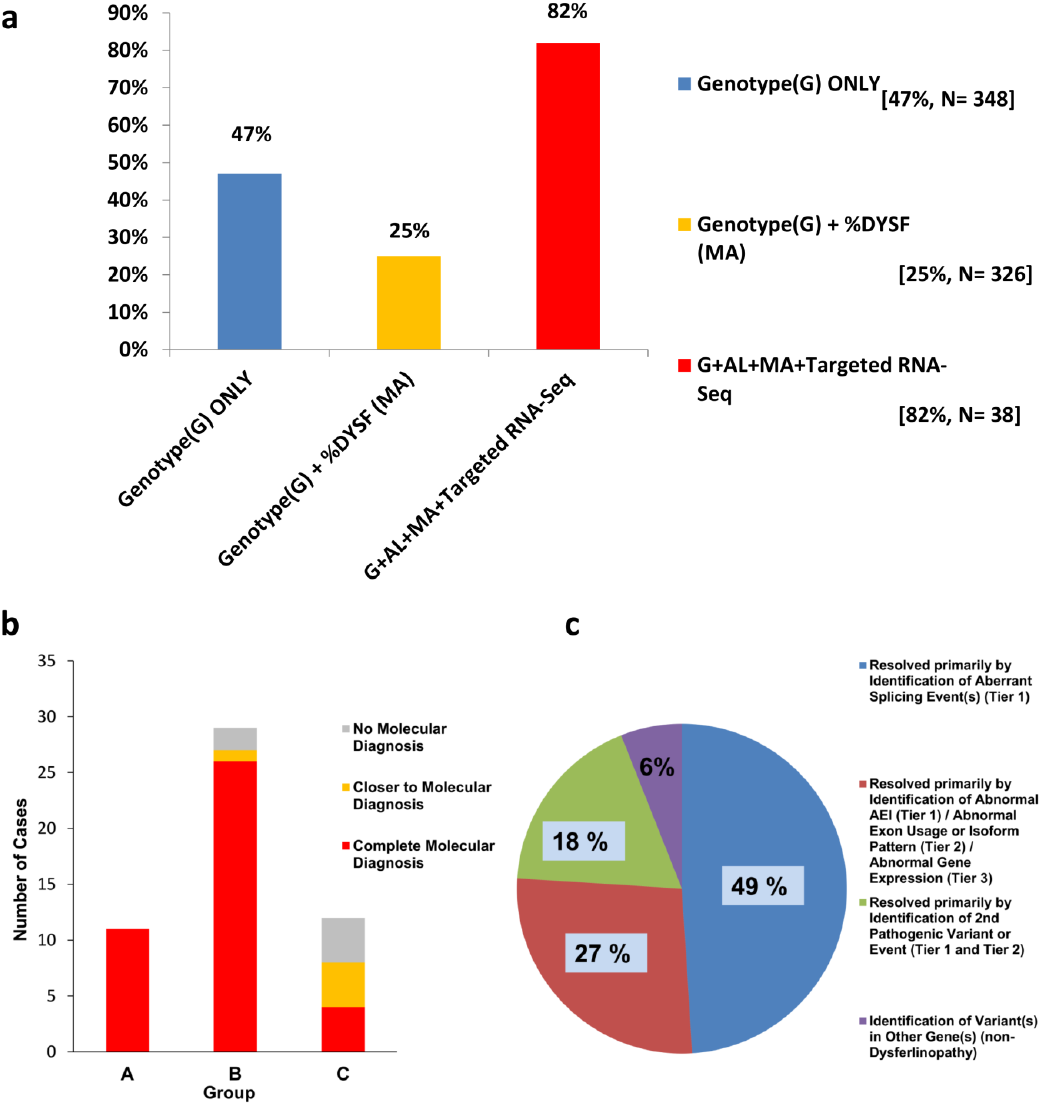
Significant increase in diagnostic yield using blood-based targeted RNA-Seq. (**a**) The diagnostic yields based on correlation between available genotype information (G) only (47%; N=348; 40%-53%; 95% CI); G and %DYSF protein expression in CD14+ monocytes (MA) (25%; N=326; 18%-27%; 95% CI); and after combining G, MA, ALDA prediction (AL) with targeted RNA-Seq (82%; N=38; 69.08% to 84.92%; 95% CI). (**b** and **c**) shows the overview of samples that underwent targeted RNA-Seq and their status after analysis. (**b**) Out of 51 samples with a suspected Dysferlinopathy or related LGMD that underwent RNA-Seq with patient informed consent (Table S5); eleven samples (Group A) had disease-range (<23%) Dysferlin protein expression in monocytes and two pathogenic (P) or likely pathogenic (LP) variants already identified via DNA sequencing and where RNA-Seq validated the mechanism of the variants’ pathogenicity and reclassified LP to P to complete the diagnosis; identification of a single (second) P/LP event or reclassification of a VUS to P/LP completed the molecular diagnosis for 28 cases (Group B). The remaining 12 cases (Group C) required two such instances to be found for a complete molecular diagnosis. (**c**) Breakdown of the RNA-Seq results for samples closer to or with a complete molecular diagnosis of Dysferlinopathy or another type of muscular dystrophy resulting from variants in a gene other than *DYSF*. 49% (22/45) of the patient cases were resolved primarily by identification of aberrant splicing event(s) using RNA-Seq Tier 1; 27% (13/45) were resolved primarily by combined identification of allele expression imbalance (AEI) (RNA-Seq Tier 1), through abnormal exon usage (RNA-Seq Tier 2), and/or through abnormal *DYSF* mRNA expression (Tier 3) ; 18% (8/45) were resolved primarily by the identification of a previously unidentified second pathogenic variant or event (RNA-Seq Tier 1 and Tier 2); and 6% (3/45) were potential non-Dysferlinopathy cases identified through the identification of variant(s) in gene(s) other than *DYSF*.

### Clinical utility of blood-based RNA-Seq

All 51 RNA-Seq samples (Table S5) were grouped based on: disease-range %DYSF with two P/LP variants where RNA-Seq validated variants’ pathogenicity mechanism and/or reclassified LP to P (Group A); identification of a single (second) P/LP event and/or a VUS-reclassification as P or LP (Group B); and lastly those that required identification of two such events described for Groups A and B (Group C) (Fig. 2B).

We confirmed the 2 *DYSF* variants are *in trans* supporting the reclassification of LP variants to P using allele-expression imbalance analysis (AEI-analysis) and *DYSF* mRNA abundance for all 14 Group A cases, 6 of which involved two nonsense/frameshift-variants, and 8 involved one non-synonymous/non-frameshift-variant. In Group B, 22 patients received confirmed diagnoses; one patient was brought closer to a confirmed diagnosis. In Group C, 4 patients received confirmed diagnoses; 4 patients were brought closer to complete diagnoses with either *DYSF* (patient C9) or another muscular-dystrophy [*DNAJB6* (patient C1), *CAPN3/VCP* (patient C4), or *COL6A2* (patient C10)].

Among the 44 cases diagnosed (31) or geno-phenotype relationship resolved (13), aberrant-splicing alone accounted for 50% (22/44) (Fig. 2C) (Figures S1-S3). Seven *DYSF* VUSs [in IVS 9 (NM_003494.3:c.907-3C>A), in exon12 (NG_008694.1(DYSF_v001):c.1171_1180+4dup14), in IVS25 (NM_003494.3:c.2643+5G>A), in exon43 (NM_003494.3:c.4794G>T), exon37-40deletion (NG_008694.1(NM_003494.3):c.3904-4410del), in exon49 (NM_003494.3:c.5503A>G), in IVS49 (NM_003494.3:c.5526-7T>G)], and 6 variants previously LP or with conflicting classification [in IVS11 (NM_003494.3:c.1053+1G>A), in IVS18 (NM_003494.3:c.1639-6T>A), in IVS22 (NM_003494.3:c.2163-2A>G), in IVS25 (NM_003494.3:c.2643+1G>A), in IVS26 (NM_003494.3:c.2810+1G>A), in IVS50 (NM_003494.3:c.5668-7G>A)] were found to alter splicing, thereby reclassified as pathogenic (PVS1,PS3-criteria)[9]. Analysis using all RNA-Seq-Tiers 1-3 (AEI/isoform-pattern/exon-usage/gene-expression) diagnosed another 26% (12/44) of cases. Using Tiers 1/2, or both, 18% (8/44) of cases were diagnosed by identifying a second pathogenic variant that was missed during prior DNA-testing. We also diagnosed 6% (3/44) cases with other forms of muscular dystrophy (*DNAJB6* (MIM 611332), *CAPN3 / VCP* (MIM 601023), and *COL6A2* (MIM 120240)). Interestingly, within Groups B and C (Fig. 2B) (Table S5), 78% (29/37) of the confirmed diagnoses came via identification of a pathogenic variant in an exonic region whereas only 19% (7/37) and 2.7% (1/37) were intronic or copy-number-variations (CNVs) and untranslated (UTR)-region variants, respectively.

Importantly, RNA-Seq also allowed better patient stratification who expressed carrier-range %DYSF. In patients A11, A12, B9, and C6 (Table S5), RNA-Seq helped explain carrier-range expression despite presence of two P/LP variants. The carrier-range %DYSF seen in patients A11 and A12 is due to the natural in-frame skip of exon 17 in majority of blood *DYSF*-transcripts, which skips around their exon 17 or IVS16-branchpoint variants in blood predicted to cause premature stop (Figure S1D). However, presence of exon17 in majority of muscle-tissue RNA-transcripts results in premature stop and lack of Dysferlin protein expression, leading to Dysferlinopathy. Overall, these results show the utility of RNA-Seq in identifying novel functional variants and aid in our understanding of the nature of their pathogenicity to understand clinical severity.

### Identification of aberrant splicing resolving geno-phenotype relationship

All *DYSF* splice aberrations identified through RNA-Seq are presented in Figures S1-S3. Three patients (B1, B4, and B5), all with disease-range %DYSF but only one previously identified P/LP variant, were diagnosed by the identification of varying-sized large-deletions all spanning entire *DYSF* exon 52 (Fig. 3A and S2A-C), causing exon 52 complete skip, and also a small-percentage of transcripts skipping both exons 52 and 53. Nonsense-mediated decay (nmd) was also observed likely contributing to the reduced %DYSF seen in all three patients. Though exon 52 deletions were previously identified[26], this is the first report of the three novel deletions causing similar splicing defects. AEI analyses demonstrated that these deletion variants are *in trans* with the second exonic *DYSF* pathogenic variants found in all three cases (Table S5 and Figure S2A-C).

**Fig. 3.**
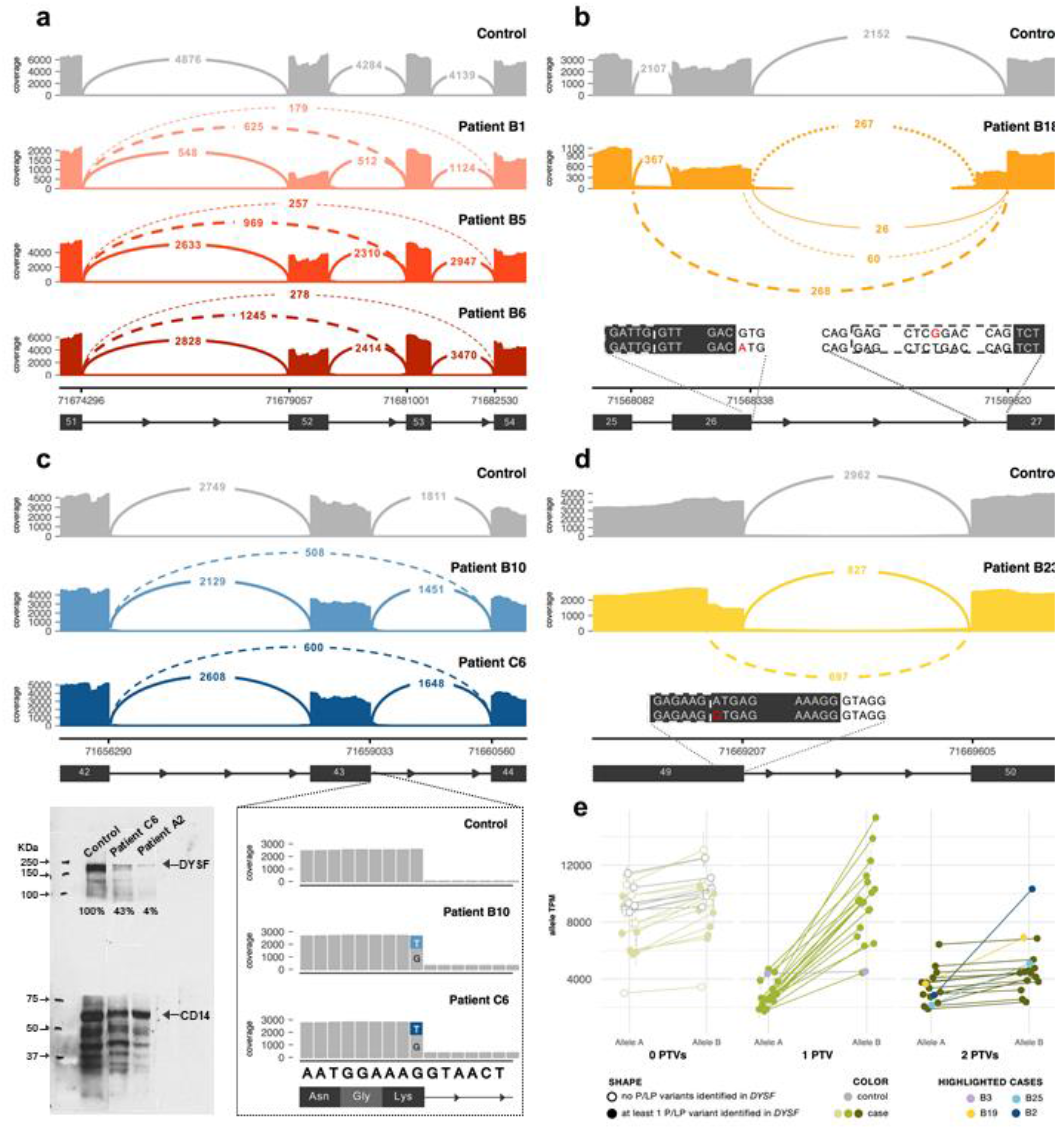
Aberrant splicing and allele expression in RNA. (**a**) Sashimi plot of the exon-skipping event seen in three patients (B1, B4, and B5). Subsequent genome sequencing (GS) identified the cause as a gross deletion encompassing exon 52. Numbers within continuous and dashed lines indicate number of spliced transcripts. (**b**) In patient B18, RNA-Seq identified exon 26 skipping caused by destruction of the splice donor site by the essential splice site IVS26 variant (left inset) and the exon 27 extension caused by a novel branch point variant (right inset). Splice events shown in (A and B) introduce a premature stop codon in the transcript. (**c**) Upper: Sashimi plot of exon-skipping caused by a leaky splice variant in exon 43 (Patients B9 and C6) that is reclassified as pathogenic. Inset: Variant expression in a minority of reads, which show a normal splice pattern. Lower Left: monocyte assay showing reduced DYSF protein expression in patient C6 (patient A2: disease-range reference). (**d**) Sashimi plot of cryptic splice site variant NM_003494.3:c.5503A>G (inset) in patient B22 which leads to a premature stop codon in the transcript. In sashimi plots in panels A through D, solid lines indicate reference splicing, while dashed lines show observed aberrant splicing. **(e)** Allele Expression Imbalance caused by nonsense-mediated decay of transcripts containing a protein truncating variant (PTV). Samples are grouped by the number of PTVs observed in DYSF mRNA. Allele “A” ratio was calculated as the smaller percent of total reads at every high quality, heterozygous, exonic SNV site in DYSF (and the opposite for Allele “B”; see Methods for further details). In each sample, the average percent and standard deviation were taken for Allele A and Allele B and mapped onto the sample’s overall gene abundance to estimate the abundance of each DYSF allele copy. Cases with one PTV show normal expression of Allele B and reduced expression of Allele A (a result of nonsense-mediated decay acting upon the transcript copy containing the PTV). Cases with 2 PTVs show a reduction in both alleles, the effect of nonsense-mediated decay acting upon both transcript copies. Highlighted cases (lavender, yellow, teal, navy) are examples of the need to use caution in the interpretation of AEI (see text).

RNA-Seq can also differentiate between multiple splice events occurring at the same locus. Patient B17 (Table S5) had only one *DYSF*-variant (NM_003494.3:c.2810+1G>A) reported before[3] and an absence of muscle Dysferlin. RNA-Seq revealed an additional aberrant splicing event using an alternate splice acceptor site in intron 26 that resulted in a 67 bp extension of exon 27 (Fig. 3B and S2J). Because these reads use the canonical exon 26 splice donor site, it is unlikely they are a consequence of NM_003494.3:c.2810+1G>A variant. Rather, we identified that another intronic variant (NM_003494.3:c.2811-20T>G) seen only in the aberrantly-spliced mRNA-reads, disrupts branch-point sequence and leads to preferential use of the alternate splice-acceptor site. This data indicates that the previously identified variant (c.2810+1G>A) and the novel branch-point variant (NM_003494.3:c.2811-20T>G) are *in trans*. Nonsense-mediated decay due to frameshift and premature protein-truncation of both splice-events agrees with the observed absence of Dysferlin protein.

Though extreme deficits in DYSF protein expression are common in Dysferlinopathies[15], some patients with a clinically-consistent Dysferlinopathy phenotype show only moderate loss of DYSF protein expression[13]. Patients B9 and C6, both with carrier-range %DYSF, were found to share a VUS in *DYSF* exon 43 (NM_003494.3:c.4794G>T, p.Lys1598His), which has been reported previously in a case with amyloid deposits[27]. RNA-Seq showed that NM_003494.3:c.4794G>T is a “leaky” splice variant, resulting in exon 43 complete skip in approximately half of the affected reads (Fig. 3C), disrupting the DYSF C2F domain and likely causing DYSF mis-folding or deposition in the amyloids seen in this patient muscle (Table S5), and since DYSF and amyloid-deposit co-localization is known to be associated with Dysferlinopathy[28, 29]. Also, these patients showed milder/slower clinical presentation/progression (muscle-weakness onset after age 45yrs, considerably later than typical onset between 17-20yrs) compared to other Dysferlinopathy patients (Table S5). Clinical course difference is possibly due to the residual 56-57% of normally spliced *DYSF*-transcripts, allowing mechanistic understanding, patient stratification and better trail-readiness.

RNA-Seq allowed us to observe mRNA structural effects that DNA-sequencing alone can miss. Patient B22 RNA-Seq showed that exon 49 NM_003494.3:c.5503A>G *DYSF* variant, predicted to be missense variant, in fact activates a cryptic splice site, resulting in a 23 bp deletion (Fig. 3D). Similarly, in patient C1 we identified a *DNAJB6* 5’UTR VUS (NM_058246.4:c.-85G>T) activating a cryptic splice site and hence reclassified as likely pathogenic suggesting LGMD1D diagnosis (Table S5). We identified 15 patients (Table S5: A13, A14, B1, B3, B4, B5, B8, B9, B11, B13, B16, B17, B24, C6, C11) with pathogenic exon-skip events due to a *DYSF* variant, suggesting exon skipping is a prevalent Dysferlinopathy mechanism.

In patient A7 (Table S5) and an additional 7 patients in the total cohort (Table S2), we identified a pseudoexon insertion (PE50.1) caused by a new splice acceptor site creation due to presence of a pathogenic deep-intronic *DYSF* IVS50 variant NM_003494.3:c.5668-824C>T (Table S2 and S5, Figure S1C), also seen recently by us in other Dysferlinopathy patients[30], causing 57% of normally spliced transcripts while 43% of transcripts spliced from PE50.1 to exon 51. This level of resolution for allelic-expression ratios is needed to better stratify patients and evaluate treatment strategy efficacy such as recent use of antisense oligonucleotides (AONs) to correct PE50.1[30].

We elucidated aberrant splicing mechanisms in four patients (A1, A3, A7, A11, Figure S1A-D), and identified 31 new splicing events in 17 patients allowing for reclassification of VUSs, novel variants, LP or P/LP variants without any previous functional evidence (Figure S2A-Q). We also identified 2 new splicing events in 2 patients (patients C6 and C11: Figure S3A-B), where two pathogenic events were needed to obtain a complete diagnosis.

### Allele expression imbalance (AEI) as a tool to phase variants in adult NMD

In previous studies, nonsense-mediated decay has prevented some protein truncating variants (PTVs) from being called in mRNA[31]. However, the extremely high read-depth by our targeted RNA-Seq panel, even without using any nonsense-mediated-decay inhibitor, allowed us to not only confidently call PTVs, but also use it to phase *DYSF* variants without parental or offspring sequencing which are not readily available for later-onset adult neuromuscular diseases such as Dysferlinopathy. AEI-analysis was able to phase *DYSF* variants in 26 cases (Table S5: A2, A5, A8, A9, A10, A11, A12, A13, A14, B1, B4, B5, B8, B11, B12, B13, B16, B17, B18, B20, B21, B23, C6, C10, C11, C12) aiding in mechanistic resolution and diagnosis. Within each sample, the allele ratio was found to be consistent for single nucleotide variants (SNVs) across *DYSF* gene. When SNVs were grouped by number of PTVs found in *DYSF*, we found that the lesser-expressed nucleotide expression (lesser allele expression) in patients with one PTV was significantly reduced to ∼25% (p=7×10^−13^) as a result of nmd. In patients with biallelic PTVs, both transcripts were subject to nmd and SNV allele-ratio generally returned to 0.5 (Fig. 3E).In patient-B18 with disease-range %DYSF, RNA-Seq determined that the NM_003494.3:c.5296G>A (p.Glu1766Lys) VUS (PM2, PP3, PP4 criteria[9]) is in *trans* with the nonsense pathogenic NM_003494.3:c.4090C>T (p.Gln136*) variant based on the inverse relationship of AEI ratios, and hence reclassified the NM_003494.3:c.5296G>A variant as pathogenic (PS3, PM3 criteria) (Fig. 3F) (Table S5).

In sample C12, phasing of *DYSF* variants had the opposite result. Reduced Dysferlin staining was seen in muscle. NM_003494.3:c.5022delT and NM_003494.3:c.401C>T (p.Pro134Leu) *DYSF* variants, previously reported as associated with reduced Dysferlin staining[32] were found to be in *cis*, indicating that NM_003494.3:c.401C>T is not pathogenic (Figure S4 and Table S5). RNA-Seq identified a different missense *DYSF* VUS [(NM_003494.3:c.6196G>A(p.Ala2066Thr)) previously reported in homozygous-state in patient with <5%DYSF[31]], in *trans* with the NM_003494.3:c.5022delT deletion, leading us to reclassify NM_003494.3:c.401C>T as benign, NM_003494.3:c.6196G>A as pathogenic, and complete Dysferlinopathy diagnosis.

### *DYSF* mRNA Expression

While muscle Dysferlin protein expression has been found to correlate well with that in blood CD14+ monocytes (Fig. 1B and [13]), the same is not true for *DYSF* mRNA-expression from whole-blood (Fig. 4A) which correlates rather better with the number of *DYSF* PTVs in the sample (Fig. 4B). PTV variants lead to nmd thus decreasing overall gene expression. As a group, samples containing biallelic PTVs exhibited >2-fold decrease in *DYSF* mRNA expression compared to those with no PTVs (Fig. 4B, p=1×10^−10^). Samples containing just one-PTV also exhibited a decrease in expression, though to a lesser degree (Fig. 4B, p=1×10^−6^). The discordance between mRNA- and protein-expression is likely explained by pathogenic missense variants and non-frameshift splicing events leading to protein non-functionality or degradation rather than mRNA-decay. For example, patient B7 (Table S5 and Figure S2D), with <10%DYSF, and homozygous intronic extended splice-site VUS (IVS49:c.5526-7T>G) which causes an in-frame insertion of two serine residues that does not affect *DYSF* mRNA expression but abolishes DYSF protein expression. This type of resolution to know at what biological level (RNA or protein in this case) the pathogenicity of the variant acts is important to use for patient stratification to reduce variability in responses to clinical trials.

**Fig. 4.**
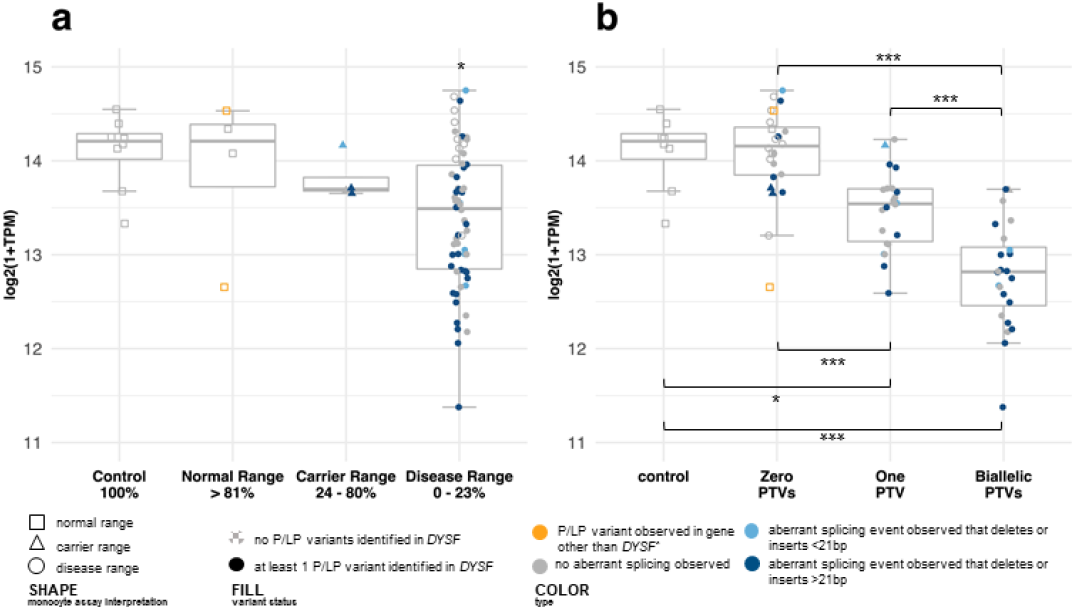
*DYSF* mRNA Expression. DYSF mRNA abundance in Transcripts per Million (TPM) categorized by (a) protein expression in monocytes and (b) number of protein truncating variants (PTVs) observed in the sample. (a) mRNA abundance does not correlate well with protein abundance in monocytes. N=78; control=8, normal range=4, carrier range=4, disease range=62. One-way ANOVA: F=3.3, p=0.03. (b) mRNA abundance correlates well with the number of DYSF PTVs observed in a sample, showing that nonsense-mediated decay is the single largest factor acting post transcription and pre-mRNA processing but prior to translation. Of interest, gross insertions or deletions caused by aberrant splicing (marked by navy blue, whether protein truncating or not), do not appear to affect mRNA abundance differently than other variants of a similar type (PTV vs PTV, non-PTV vs non-PTV). N=78; control=8, zero PTVs=24, one PTV=23, biallelic PTVs=23. One-way ANOVA: F=33.3, p=1e-13. Post-hoc comparisons (t-tests with Bonferroni correction for multiple tests): p(control vs 1 PTV)=0.006, p(control vs 2 PTVs)=0.0008, p(0 PTVs vs 1 PTV)=9.2e-05, p(0 PTVs vs 2 PTVs)=1.1e-09, p(1 PTV vs 2 PTVs)=0.0002.

Gene- and allele-expression did not match what was expected based on the identified *DYSF* variants for samples A12, and B2 (Table S5). Patient B2 had <5%DYSF with only one *DYSF* variant (NM_003494.3:c.6216delC (p.Met2073*)) in exon 55. But average allele expression ratio for SNVs across *DYSF* was 0.49, which is abnormal compared to all other samples with one-PTV (Fig. 3E). It is possible that DYSF p.Met2073* is so close to the reference-stop (Fig. 5) that the transcript is not targeted for nonsense-mediated decay. However, the *DYSF* mRNA-expression is extremely low, suggesting the presence of biallelic PTVs. Subsequent GS found several rare deep-intronic *DYSF* variants, most notably an ultra-rare NM_003494.3:c.4509+1586dupG that could possibly cause a regulatory effect, but pathogenicity confirmation could not be achieved. However, based on the combination of one clearly pathogenic *DYSF* variant, potentially pathogenic deep-intronic *DYSF* variant, disease-range %DYSF, and related clinical features, a Dysferlinopathy diagnosis was achieved.

**Fig. 5.**
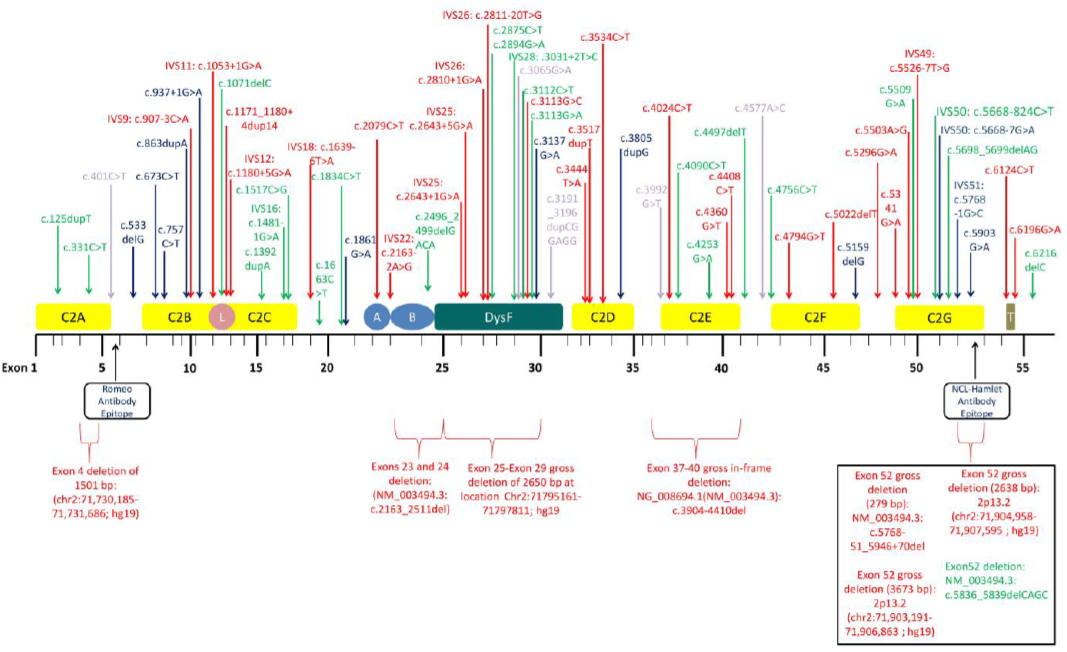
Mapping of Reclassified Variants by RNA-Seq on Dysferlin Domains Relative to Exons. Dysferlin contains seven calcium-dependent C2 lipid binding domains (C2A-C2G), a ferl domain (L), FerA and FerB domains (A and B, respectively), a unique DysF domain specific to Dysferlin, and a transmembrane domain (T). The C2 and transmembrane domains are known to have a function in membrane repair. The NCL-Hamlet antibody epitope towards the C-terminal region and the Romeo antibody epitope towards the N-terminal region of Dysferlin are shown. Variants of uncertain significance (VUSs) that have been reclassified by RNA-Seq to pathogenic (P) or likely pathogenic (LP) are red color coded variants and those reclassified as benign (B) or likely benign (LB) by RNA-Seq are light-purple color coded variants; blue color coded variants indicates those with conflicting or non-confirmatory interpretations (P/LP or LP) that have been reclassified as P by RNA-Seq; and green color coded ones are pathogenic variants whose mechanisms of pathogenicity was revealed by RNA-Seq. All variant positions correspond to *DYSF* transcript NM_003494.3.

## DISCUSSION

We have clearly demonstrated the use of phenotype-driven blood-biomarker functional genomic combinatorial approach to provide better genotype-phenotype-resolution and high diagnostic-yield for a neuromuscular-disorder type. In this new genomics era, intersection of clinical genetics and research genetics based tools needs to be considered to identify more efficient indicators for disease mechanisms, diagnostics, biomarkers and therapy. While muscle is the main target tissue for NMD evaluation, it is possible to use a tiered combinatorial approach, such as done for this study, for NMD genes that are also adequately expressed in blood (Table S1) or urine or to find other biomarkers that may expedite the diagnostic odyssey NMD patients face.

*DYSF* is a large gene comprising 55 exons and spanning a genomic region of >150 kb[33]. Although variant detection provides the most definitive diagnosis, it is highly expensive considering the large number of exons involved. Moreover, the recessively inherited disease is often seen to be caused by single nucleotide changes with no previous substantial evidence of recurrence[33, 34]. Therefore, protein expression analysis is preferable and aids in diagnostics. Currently most neuromuscular physicians do not prefer protein expression analysis as a first phase of diagnosis since it requires a muscle biopsy, which is painful, invasive, and costly to perform. A less invasive, inexpensive, and rapid method involving a non-muscle tissue was developed for protein analysis, specifically for dysferlin protein[34] in patient peripheral blood mononuclear cells (PBMCs), which we and colleagues have previously shown correlates well with expression in the disease target skeletal muscle tissue in both healthy and Dysferlinopathy conditions[14-16]. We further refined and modified this assay to a more clinically relevant novel method by specifically isolating CD14+ monocytes which allowed us to better stratify patient samples to unaffected, carrier- or disease-range (Fig. 1B) based on DYSF protein expression, but was not as diagnostic as desired (Fig. 2A). Higher level of resolution was needed to create a more robust carrier testing platform and genotype-phenotype correlation, especially since recently we have shown that Dysferlinopathy is the second most prevalent LGMD in the US with the highest percentage of homozygous variants indicating the greater need for carrier testing[1].

The current diagnostic pathway for Dysferlinopathy typically only assesses Dysferlin protein expression and *DYSF* DNA-sequencing[35], but here we show that by using a tiered analytical approach we can tease apart all possible aberrations at RNA-transcript structural-, exonic-, allelic-, gene-, and protein-expression levels that will lead to not only a higher diagnostic yield but also important help enable patient stratification by connecting molecular underpinnings to clinical severities. One limitation of this study is that the prior DNA-testing were done non-uniformly (Exome/Gene-Panel/Sanger) based on respective physician discretion, but it does reflect the real world heterogeneous scenario of clinical genetics diagnostic requisitions from our experience. Our combinational phenotype-driven approach helps one understand if the variant pathogenicity mechanisms are acting at mRNA expression level, or protein structural or functional level. This mechanistic information is important for diagnostics, better trail-readiness, and the evaluation of efficacy measurements in clinical trials that are based on gene- / mRNA- / AON- / protein- / chemical-therapy. Hence, the monocyte assay and/or targeted RNA-Seq correlated with clinical data could also potentially be used for efficacy testing and monitoring of trials that are based on targeting the DNA or RNA. Such biomarker-based testing, such as the evaluation of human urine extracellular mRNA to identify splice variant biomarkers of myotonic dystrophy (DM1) and Duchenne muscular dystrophy (DMD)[36] are upcoming. Combining the power of functional omics platforms with biomarker strategies can significantly increase efficiency of NMD diagnostics, clinical-trial readiness, effectiveness, and outcome measurements, as well as novel therapy discoveries[10, 24].

In the DYSF mRNA of our cohort, we observed allele expression imbalance driven almost entirely by nonsense-mediated decay (Figure 4e). The high read depth achieved by our targeted sequencing combined with the consistency of the observed effect of nonsense-mediated decay allowed us to reliably phase exonic DYSF SNVs in samples with a single protein truncating variant (PTV) identified in DYSF. In each of these samples, the PTV was seen in approximately 25% of the total reads at that site and the reference allele made up the remaining ∼75% (Figure 4e, second panel). Because nonsense-mediated decay acts upon the entire length of the transcript, all other exonic SNVs across DYSF exhibited a similar AEI ratio such that we could determine whether each SNV was in cis or trans with the PTV. For example, in patient A2, the nonsense variant c.331C>T (p.Q111X) in exon 4 is called in 168 out of 993 total reads at that site (∼17%). Three other variants were called in the mRNA of patient A2: synonymous SNP c.1827T>C (p.D609D) in exon 20 (1410 of 1680 total reads, 84%), synonymous SNP c.2583A>T (p.S861S) in exon 25 (1465 of 1839 total reads, 80%), and the pathogenic/likely pathogenic missense variant c.6124C>T (p.R2042C) in exon 54 (1535 of 2000 total reads, 77%). Based on the observed AEI, we can be confident that all three of the other variants are in trans with the exon 4 nonsense variant. This method of phasing has the advantage of not requiring trio sequencing but is of course not perfect and should be used cautiously. Only one PTV was identified in Patient B3 (highlighted lavender in Figure 4e), but the observed AEI more closely matches that of samples with zero or two PTVs. Because the truncating variant seen is located in the last exon of DYSF, it is possible that this variant does not lead to nonsense-mediated decay. Another possibility is that there is a second PTV that we were unable to identify, since the overall DYSF mRNA abundance is also reduced compared to controls.

When a sample contains two PTVs the average AEI ratio of heterozygous SNVs returns to around 50/50, similar to the ratio seen in samples and controls with no PTVs (Figure 4e, first and third panels). In these samples we cannot phase each individual SNV in DYSF but can still reliably determine that the PTVs are in trans with one another. Because insertions and deletions, a main type of pathogenic PTVs, are known to be more difficult than SNVs to align and call accurately1,2, we cannot rely on the AEI seen for these variants but instead use the AEI of the samples other DYSF SNVs and extrapolate what we observe in samples with one PTV. If the PTVs were in cis, we would expect the sample’s AEI ratio to be similar or more extreme than that of cases with one PTV. Instead, we see both Allele A and Allele B to be reduced in abundance at similar levels to Allele A in cases with one PTV, supporting that nonsense-mediated decay is acting on both transcript copies and the PTVs are in trans. Again, some samples were observed not to follow this pattern. In two of these cases (highlighted teal and navy in figure 4e), the deviation can be clearly attributed to the location of one PTV. Patient B25 has a splice variant in IVS39 that can be seen in mRNA to result in multiple aberrant splicing events. Some aberrantly spliced reads skip both exon 40 and the alternate isoform exon 40a, which we observed to be naturally excluded in around half of blood transcripts (average percent-spliced-in: 41%), meaning we are likely not seeing the full extent of this aberrant splicing event in blood.

With this context, the slight bias seen in the AEI of this patient is expected. Patient B2 contains a nonsense variant in exon 17, an exon used variably by isoforms and only included in less than 10% of transcripts expressed in blood (average percent-spliced-in: 9%). As a result, the AEI observed in patient B2 is driven by the nonsense variant in exon 40 and resembles the AEI seen in samples with only one PTV. The natural skipping of exon 17 in the majority of blood transcripts also explains why this patient was found to have carrier range protein levels in the monocyte assay.

The cause of the AEI bias seen in patient B16, however, is not clear (Figure 4e, highlighted yellow). The protein truncating variants in this sample are an insertion located in exon 2 and a deletion located in exon 46. Both of these exons are constitutively expressed in all isoforms, and we are unable to ascertain whether the AEI bias is due to one of the variants, a sample quality or sequencing issue, or another factor. In this sample we cannot be confident that the PTVs are in trans, an example of the caution that should be used in the interpretation of AEI.

Allele Expression Imbalance relies on the quality of variant calls in mRNA, and a number of technical factors must be considered. One major caveat to variant phasing using this method is SNVs near the end of exons. In order to call variants in mRNA using the GATK pipeline (originally developed for DNA sequencing reads), reads must be split at the splice junction points determined during alignment. As a result, variants at the ends of exons are found mostly at what the GATK tools view as the “end” of the read, which is often where sequencing errors are found. Questionable variant proportions should be checked by comparing the HaplotypeCaller bamout to the input bam. Another area to exercise caution in AEI interpretation is with aberrant splicing events. The majority of mRNA reads no longer contain the underlying causative intronic variant due to intron excision, so read counts for these events should be obtained from splice junction mapping counts. Because these variants frequently result in gross deletions or splicing that appears to be non-canonical to alignment tools, leading to difficulty mapping these reads, AEI balance should be interpreted with the same caution as exonic indels would be (https://bmcbioinformatics.biomedcentral.com/articles/10.1186/s12859-019-2928-9 https://academic.oup.com/bib/article/18/6/973/2562816)

For *DYSF*, there are no distinct variant hotspots and the pathogenic variant distribution is found to be uniform on the exon-domain landscape (Fig. 5). This finding highlights the challenges of pursuing therapeutic options that target particular *DYSF* regions (e.g. exon-skipping[37, 38]) or correct individual variants (e.g. CRISPR) and instead points to the direction of therapies that are independent of specific variants (e.g. gene-therapy) such as the dual-vector strategy for whole *DYSF* cDNA-transfer or modified steroid-therapy which have both shown promising results in dysferlin-deficient animal models[39, 40]. Moreover, most, if not all, of the nonsense variants we evaluated showed significant nonsense-mediated decay which has implications on the potency of nonsense-mediated read-through drugs like Ataluren[41] in NMDs since the drug target (i.e. the *DYSF*-mRNA with the nonsense variant) is significantly reduced.

In conclusion, we have clearly shown the clinical utility of our combinatorial multi-level biomarker-driven functional genomics approach to elucidate variant pathological mechanisms, to understand genotype-phenotype correlations, and significantly enhance diagnostic yield in Dysferlinopathy. Importantly, such analysis enables greater patient stratification which in turn increases readiness for clinical trials and precision medicine initiatives for neuromuscular and other genetically based disorders.

## Data Availability

The original contributions presented in the study are included in the article/supplementary material; further inquiries can be directed to the corresponding author.

## SUPPLEMENTARY INFORMATION

Supplemental Information include Supplementary Materials and Methods, four Supplementary Figures [Figures S1 (A-D), S2 (A-Q), S3 (A-B), and S4] and four Supplementary Tables (Tables S1-S5).

## ACKNOWLEDGEMENTS

We thank all the patients and their families and individuals who provided samples to be used for normal controls for consenting to enroll in our study. We also thank all the Clinical Outcome Study (COS) PIs and members of the JAIN COS consortium for the recruitment and collection of samples and data from patients enrolled in COS, as well as the Newcastle MRC Centre for Neuromuscular diseases biobank and Dr. Hanns Lochmüller of University of Newcastle Upon Tyne (former director of the MRC and currently of University of Ottawa) for providing the COS specimens with informed consent. We thank the Georgia Institute of Technology Molecular Evolution Core Sequencing Facility. S.S. and L.G. were supported by the Jain Foundation Focused Research Grant to S.C. G.G. is an investigator of the Georgia Institute of Technology. B.R.R.N., A.K., and M.H. are employees of Perkin Elmer Genomics. The content of this manuscript is solely the responsibility of the authors and does not necessarily represent the official views of the funding agencies.

