## Supplementary Materials for "Combinatorial clinically driven blood biomarker functional genomics significantly enhances genotype-phenotype resolution and diagnostics in neuromuscular disease"

### SUPPLEMENTARY INFORMATION

#### TABLE OF CONTENTS

| **SUPPLEMENTARY MATERIALS AND METHODS** | **Page Numbers** |
|---|---|
| Sample Size | 6 |
| Data Exclusions | 6-7 |
| Replication | 7 |
| Randomization | 7 |
| Blinding | 7-8 |
| Clinical Evaluation | 8 |
| Patient Recruitment | 8-9 |
| Patient Enrollment Questionnaires | 9-16 |
| CD14+ Monocyte Assay and Data Analysis | 17 |
| Protocol of CD14+ Monocyte Assay | 18-29 |
| Whole Blood Targeted RNA-Seq Library Preparation and Sequencing | 30 |
| Targeted RNA-Seq Tiered Analysis Approach | 30-32 |
| RNA-Seq Alignment and Quality Control | 32-33 |
| RNA-Seq Variant Calling | 33-34 |
| RNA-Seq Splicing Analysis | 34-35 |
| RNA-Seq Allele Expression Imbalance | 35 |
| RNA-Seq Gene Expression Analysis | 35 |
| Whole Genome Sequencing (WGS or GS) and Analysis | 35-37 |
| Neuromuscular disease (NMD) 131 gene-panel used for NGS DNA sequencing | 37-39 |

Neuromuscular Disease (NMD) Panel DNA-Sequencing with
Deletion-Duplication Analysis 39

Sanger Confirmation of Genomic DNA 40

**SUPPLEMENTARY FIGURES**

**Figure S1 (A-D).** Sashimi and/or IGV plots of cases with new *DYSF* splicing mechanisms identified due to previously reported variant with prior pathogenic (P) or likely pathogenic (LP) classification confirming molecular diagnosis 41-44

**Figure S2 (A-Q).** Sashimi and/or IGV plots of cases with *DYSF* splicing events identified related to a single pathogenic event or reclassification of VUS or discovery of a single pathogenic mechanism that completes molecular diagnosis 45-66

**Figure S3 (A-B).** Sashimi and/or IGV plots of cases with *DYSF* splicing events identified related to two pathogenic events or reclassification of VUSs or discovery of two pathogenic mechanisms that completes molecular diagnosis 67-69

**Figure S4.** IGV plot of patient C12 showing identification of a third new *DYSF* variant not previously reported in genetic testing, *in trans* based on AEI confirming molecular diagnosis 70-71

**SUPPLEMENTARY TABLES**

**Table S1 (Excel spreadsheet).** Neuromuscular Disease (NMD) Gene Panel for Targeted RNA-Seq.

**Table S2 (Excel spreadsheet).** List of all de-identified patient cases (364 patients and 15 normal individual controls) and their molecular diagnosis based on correlation between genotype, %DYSF protein interpretation in monocyte assay, and ALDA before RNA-Seq.

**Table S3 (Excel spreadsheet).** Correlation between Genotype and %DYSF Range in Monocyte Assay (refer Table S2)

**Table S4 (Excel spreadsheet).** List of all Multi-genic patient cases with initial clinical Dysferlinopathy suspicion showing reduced Dysferlin protein expression

**Table S5 (Excel spreadsheet).** Diagnosis by RNA-Seq: DYSF genotype, proteotype, RNA-Seq result, and clinical data of all 51 patient samples RNA-Sequenced.

**SUPPLEMENTARY REFERENCES** 72-75

#### SUPPLEMENTARY MATERIALS AND METHODS

##### Sample Size

Our total sample size is 364 patient samples and 15 normal control samples from unaffected individuals. The number of samples was based on the number of patients who we were able to recruit based on inclusion/exclusion criteria described below and in main text. The sample size in the study is substantial (364 individual patient samples) and fulfills power analysis requirement of a large cohort of multiple ethnicities in order to understand the spectrum of genotype-phenotype correlation in the disease, based on our previous studies and experience. All the covariates related to our patient population including gender, genotype information, initial diagnosis during recruitment and final diagnosis based on our study are provided (Table S2, and S4), whenever the data is available or provided by the patient or confirmed in this study. Gender of the patient population was roughly male:female=50%:50% based on gender data that were available to us. All patients were initially clinically suspected of Dysferlinopathy (LGMD2B or Miyoshi Myopathy or Distal Myopathy with Anterior Tibial Onset or related LGMD) or related myopathy. We used normal control samples from 15 unaffected individuals of different ethnicities without any history of any neuromuscular or other genetic disease symptoms, sufficient for internal controls to confirm molecular diagnostics, and for genotype-phenotype correlation.

##### Data Exclusions

Samples were not processed if reached us after 24 hrs from blood draw based on phlebotomist signed time on top of blood collection tubes in order to remove any discrepancy for RNA or protein degradation. RNA-Seq data from any specific patient sample was excluded if outlier status based on tissue composition or contamination was found based on principle component analysis (PCA) in initial Quality Control step of RNA-Seq data analysis.

**Replication**

Replicates were run at the same time for all western blots and for each sample. Two blots were run each time and each blot contained the samples from the same group of patients, as described in the Supplementary Materials and Methods “Protocol of CD14+ Monocyte Assay” section below. Normal control samples were collected for processing from same 15 normal control individuals after every two months till study completion to remove any possibility of protein or RNA degradation during storage. No discrepancy occurred during any assay repeat of any sample.

**Randomization**

Patients from different ethnicities were recruited based on specific inclusion/exclusion criteria as described above and in main text, and were the experimental group, without any bias. Normal unaffected individuals with different ethnicities were the control group.

**Blinding**

Blinding was performed by keeping the genotype and clinical data of patients in a locked cabinet of first corresponding author's office during CD14+ monocyte assay and analysis of results and by giving internal IDs for each patient sample. Similar approach was performed for RNA-Seq analysis. Similarly, for DNA-Seq analysis such as that of known variant testing, neuromuscular disease panel testing or whole genome sequencing, monocyte assay and RNA-Seq data was not available during analysis for blinding purpose, and only used to interpret once a reportable genetic variant was identified. These are following the approved Institutional Review Board protocol.

**Clinical Evaluation**

All patients in this study underwent comprehensive clinical evaluation by their respective physicians and Dysferlinopathy or a closely related LGMD or myopathy was suspected. For any participant who agreed, the patient and/or their respective physician were asked a full list of relevant questions (see Supplementary Materials and Methods “Patient Enrollment Questionnaires” section below) regarding their clinical symptoms, clinical history, and ethnicity in order to assess the patient’s phenotype. Clinical, ethnic and family history data were collected whenever the information was available to us. Clinical history including age of onset, initial symptoms, region in which weakness first started, and functional status of the patient was collected where detailed clinical notes were available. Pattern of weakness including upper limbs or lower limbs, or both, proximal or distal or both, symmetrical or asymmetrical patterns of weakness, and differential weakness, was collected when available. Detailed pedigree analysis was also performed when possible.

**Patient Recruitment**

All the recruited cases were suspected to have Dysferlinopathy or related LGMD/myopathy based on clinical presentation. Patients for this study were recruited with the help of Jain Foundation, who contacted the foundation for diagnostic support and from the Clinical Outcome

Study for Dysferlinopathy (COS) and thereafter were recruited in this study by first corresponding author. For the COS cases (Serial Number 326-364 in Table S2), the de-identified RNA and DNA used in the study was obtained from the Euro biobank in Newcastle, UK. These samples were given to the biobank after proper consent was obtained as part of the Clinical Outcome Study for Dysferlinopathy (COS) and transferred to this study using Material Transfer Agreement between institutions.

**Patient Enrollment Questionnaires**

1. **Is there anyone in the patient's families with similar symptoms? If so, are their symptoms the same, milder, or more severe than the patient's?**

NOTE: list all family members and how their symptoms compare

- Brother
- Sister
- Father
- Mother
- Son
- Daughter
- Maternal Uncle
- Other aunt or Uncle
- Male first cousin
- Female first cousin
- Other

**2. Ancestry (select all that apply)**

- Finnish

- Other Northern European

- Japanese

- Not listed (specify)

- Unknown

**3. Have any LGMD subtypes or other conditions been ruled out for this patient by genetic testing? NOTE: It is recommended that you initially run the tool without any exclusions**

- Yes

- No

- Unknown

If yes, check all the diseases that apply

2A 2B SG's 2G 2H 2I 2J DG's 2L 2Q

1A 1B 1C 1D 1E 1F 1G 1H Nonaka/HIBM

Tibal Becker DMD Manifesting Carrier FSH EDMD

Pompe Bethlem

**4. Age of onset of symptoms**

**NOTE: Onset of symptoms should not include high CK without weakness symptoms or subclinical changes e.g. on MRI.**

- <5 years

- 5-12

- 13-25

- >25

- Unknown

**5. If you had an electromyogram (EMG), what were the results? This is the test in which needles are stuck into muscles to record nerve impulses. Go to http://www.nlm.nih.gov/medlineplus/ency/article/003929.htm for a description of EMG**

- The EMG showed a muscle problem (myopathic pattern)
- The EMG showed a nerve problem (neurogenic pattern)
- I have never had an EMG or I don't remember the results

**6. Distal Weakness or Proximal Weakness?**

- Primarily Distal
- Comparable to or less than proximal weakness
- None, or much less than proximal weakness with much later onset
- Unknown

**7. Facial weakness**

**NOTE: Examples of this could be difficulty chewing, whistling, making normal facial expression. Also, onset is particularly early in the disease course prior to having severe generalized weakness.**

- Yes
- No
- Unknown

**8. Did you have nervous system symptoms (for example: headaches, tiredness, sleep disorders) which began within one year of the onset of muscle weakness? If yes, do you have respiratory difficulties?**

- I have some of these symptoms, and I have respiratory difficulties.
- I have some of these symptoms, but I do not have respiratory difficulties.

- I do not have any of these symptoms.
- I don't know

**9. Scapular Winging**

**NOTE: If the answer is NO, then you need to determine for how long they have had muscular dystrophy symptoms. If it has been for less than a couple of years, then the answer should be UNKNOWN because we don’t know how soon patients start to show scapular winging**

- Pronounced
- Mild
- No
- Unknown

**10. Calf Hypertrophy**

**Note: Answer PRONOUNCED or MILD if calf muscles were enlarged earlier in the disease course, even if they aren’t now**

- Pronounced
- Mild
- No
- Unknown

13. Pain/Cramps

**11. Asymmetry of Weakness/Wasting**

- Pronounced (persistent)
- Mild
- No

- Unknown

**12. Rate of progression**

**NOTE: descriptions of each rate can be found below**

- Rapid: significant increase in weakness within a few years, causing loss of function and/or walking and/or involvement of other muscle groups beyond the first ones affected.

- Moderate: increase in weakness over several years, but only a slight increase in weakness from year to year. No loss of walking or great difficulty walking within 10 years of onset.

- Slow: very gradual or no decrease in strength for several years following diagnosis. Function appears stable or nearly so.

- Unknown: answer this if a detailed history isn't available, or if patient has only recently experienced symptoms (<3 years) and there has not been noticeable progression.

**13. Biopsy: Dystrophic**

**NOTE: Does the patient's muscle biopsy show a dystrophic appearance (such as fiber size variation, central nucleation, focal necrosis, fiber replacement by fatty or connective tissue)?**

- Yes

- No

- Unknown

**14. Biopsy: Inflammation –**

**Note: Does patient's biopsy show inflammatory characteristics (e.g., substantial levels of mononuclear cell infiltration?**

- Yes

- No

- Unknown

**15. Biopsy: Vacuoles/Inclusion Bodies –**

**NOTE: Are rimmed vacuoles observed (in substantial numbers) on patient's biopsy?**

- Yes
- No
- Unknown

**16. CK results**

**NOTE: If the patient has more than one CK measurement, use the one taken closest to the onset of the patient's muscle symptoms**

- Unknown/Not Measured
- <200
- 200-500
- 500-2000
- 2000-7000
- >7000

**17. Does the patient's biopsy show aggregation of desmin or other myofibrillar proteins (conditions with this biopsy appearance are often referred to as a Myofibrillar Myopathy)?**

- Yes
- No
- Unknown

**18. Finger contractures**

- Yes
- No

- Unknown

**19. Toe walking/Achilles tendon tightness**

- Yes

- No

- Unknown

**20. Foot drop**

**Do you trip often or has the doctor ever said you have a foot drop?**

**Explanation: This is where you trip over your own foot because you can't pull your foot through the walking motion (lack of dorsiflexion).**

- Yes

- No

- Unknown

**21. Inability to stand on toes**

- Unable

- Able

- Unknown

**22. Quadriceps Strength (early in disease course if patient now has severe generalized weakness)**

- More affected than other proximal leg muscles

- Affected comparably to other proximal leg muscles

- Selectively spared compared to other proximal leg muscles

- Unknown

**23. Neck weakness**

- Yes

- No

- Unknown

**24. Legs or arms more affected**

- Legs

- Arms

- Arms and legs comparable

- Unknown

**25. Cardiac conduction defect**

**Do you have a pacemaker, or abnormal heart rhythm?**

- Yes

- No

- Unknown

**26. Dilated or Hypertrophic Cardiomyopathy**

**Has your doctor ever told you that you have a "dilated cardiomyopathy" or "hypertrophic cardiomyopathy"?**

- Yes (dilated)

- Yes (hypertrophic)

- No

- Unknown

**CD14+ Monocyte Assay and Data Analysis**

Peripheral blood was collected into EDTA-treated collection tubes by a trained phlebotomist and maintained at 4$^{o}$C during shipping as well as storage. Isolation of CD14+ monocytes was performed using EasySep direct Human monocyte isolation kit (Cat #19669, Stem cell technologies). Cell pellets obtained by centrifugation were lysed with membrane protein extraction reagent (M-PER) and whole protein lysate was prepared. All lysates were quantified for total protein by BioRad Bradford assay. Immunoblotting analysis was performed using known amounts of protein lysate by SDS-gel electrophoresis. Monoclonal dysferlin antibodies (NCL-Hamlet or Romeo) were used at 1:3000 (NCL-Hamlet) or 1:1000 (Romeo) working concentration for primary antibody incubation. Horseradish peroxidase (HRP)-linked IgG antibody (Cat. No. NXA931; GE Healthcare) was used at 1:2500 (if primary is NCL-Hamlet) or Goat Anti-Rabbit IgG H&L (HRP) (Cat. No. ab205718; abcam) was used at 1:2000 (if primary is Romeo) working concentration for secondary antibody incubation. Chemiluminescence detection was performed using an ECL Plus system (Cat. No. RPN2132; BioRad). Parallel immunoblotting was performed for both the target DYSF protein as well as the CD14 protein as loading and a housekeeping protein control. Densitometry analysis on signal intensity for Dysferlin was done and further normalized using the CD14 protein intensity mentioned above, using Image Studio™ Lite Version 5.2 (LI-COR Biosciences) (https://www.licor.com/bio/image-studio-lite/). For detailed protocol see Supplementary Materials and Methods "Protocol of CD14+ Monocyte Assay" section below. All monocyte assay results were reported back to patients and/or respective physicians as research reports according to guidelines of the approved Institutional Review Board protocol.

**Protocol of CD14+ Monocyte Assay (significant modulation and refinement for clinical utility from our previous studies[1-3])**

**Summary:** A blood-based screen for defective dysferlin protein expression by immunoblot analysis using blood monocytes.

**Procedural Overview:**

A) Assigning ID to samples ………………….. 10min
B) Isolation of Monocytes………………………3 hrs
C) Lysate Preparation……………………………1 hr
D) Protein Quantification……………………… 1 hr
E) SDS-PAGE …………………………………1 hrs
F) Immunoblotting…………… ……………… 6.0 hrs
G) Developing the film…………………………...1hr
H) Scanning images……………….……………….30min
I) Quantification………………………………….3hrs

**A. Assigning ID to samples**

1. Once the sample arrives, a Number is assigned to the sample. The EDTA sample tubes from the same patient is marked as number A and number B (For example: - 1A and 1B).
2. The number is also noted on the consent form along with the D.O.B of the person.
3. The number, patient's name and date of birth is added into the internal database

**B. Isolation of blood monocytes.**

**Required Reagents and materials.**

- EasySep direct Human monocyte isolation kit. Cat #19669 by Stem cell technologies. (to negatively isolate blood CD14+ monocytes)
- Easy eights easy sep magnet. Cat #18103 by Stem cell technologies.
- Corning Cellgro D-PBS (without calcium and magnesium). Cat # 21-040-CV.
- Ultra-Pure 0.5M EDTA, pH 8. Cat # 15575-038 by Invitrogen.
- 14 ml Falcon round- bottom disposable tubes by Corning. Cat #352057.
- 10 ml Sterile serological pipettes.
- Disposable transfer pipettes.

<u>Procedure.</u>

1.Mix the blood in the EDTA tube by inverting. Carefully pipette 3ml of blood from each of the EDTA tubes using a disposable transfer pipette and add to two labelled 14ml round-bottom disposable tubes.

2.Vortex the rapid spheres for 30seconds.

3.Add isolation cocktail (50µl/ml), 150 µl to each of the 3 ml sample tube. Add rapid spheres (50µl/ml), 150 µl to each of the sample tubes. Mix and incubate at RT for 5min **(On tube holder not on magnet).**

4. Add 9ml of 1XPBS containing 1mMEDTA, (3x Volume of original sample), to each of the tubes and pipette up and down 2-3 times.
5. Place the tube without lid into the magnet and incubate at RT for 5min.
6. Carefully pipette the cell suspension (**using 10ml pipette and do not pour**) into a new labelled 14ml round- bottom tube. Don’t forget to include some RBC solution **(10%, i.e., around 1ml, not more than that)**.

7. Again add 150 µl of rapid spheres, to each of enriched samples contained in new tubes. Mix and incubate for 5min, RT.
8. Remove the old tubes from the magnet and place the new tubes, without lid, containing enriched cells and rapid spheres into the magnet. Incubate for 5min at RT.
9. Carefully pipette the clear enriched solution, into a new labelled round-bottom tube (all at once using 10ml pipette, **do not pour**. At this step, collect only clear solution, **no RBC solution must be collected. (No rapid spheres must be added at this step)**.
10. Remove the old tubes from the magnet and place new tube containing enriched cells and without lid into the magnet. Incubate at RT for 5min.
11. Carefully pipette the enriched cell suspension into a new, labelled 15ml centrifuge tube (all at once using 10ml pipette, **do not pour**). Collect only clear solution.
12. Centrifuge at 800rpm (120 X *g*) for 10min, RT.
13. Remove the supernatant and resuspend the pellet of one tube of a sample in 8ml PBS containing EDTA, pipette all the solution including cells. Then resuspend the pellet of 2$^{nd}$ tube of the same patient using 1$^{st}$ tube's re-suspended solution. Centrifuge at 1200rpm (300 X *g*) for 10min and remove supernatant.
14. The pellet does not have to be re-suspended if protocol has to be continued. Go to step 2 of next protocol.
15. If protocol has to be stopped, remove the supernatant and re-suspend the washed pellet in 50µl 1XPBS containing 1mM EDTA containing 1 mM EDTA (store at 4°C for 1-2 days).

**C. Lysate preparation**. *(Adapted and modified from Pierce's M-per protocol*).

Required reagents

M-PER .CAT #78503 from PIERCE.

Roche complete EDTA –free protease inhibitor cocktail. Roche Cat no#11836170001

1. If using stored, Pellet the monocyte cell suspension by centrifuging @2,500 X *g* for 5min at RT and carefully removing supernatant.
2. Add 200µl of MPER solution to wet pellet/suspension, with a 200µl micropipette and shake the mixture.
3. Immediately add 34µl of 7X protease inhibitor cocktail. Shake and vortex the mixture vigorously for 10min, RT (till pellet is broken).
4. Transfer all contents into 1.7ml eppendorf tubes.
5. Remove cell debris by centrifugation at 10,000rpm, 10min and 4˚C (cold centrifuge).
6. Collect the supernatant into a 0.6ml tube, tap it, quick spin and then aliquot 8µl of sample into a 0.6ml tube for BCA assay, aliquot 23µl of the supernatant into 5, 0.6ml tubes and retain the rest in the same 0.6ml tube. Store at -20˚C.

**D. SDS PAGE**

<u>Required Reagents</u>

Bio-Rad Mini Protean TGX gel- (10%, 30µl/10-lane) Cat no: #456-1033.

Bio-Rad Kaleidoscope ladder –precision protein standard. Cat no: #161-0375.

Bio-Rad B-Mercaptoethanol- Cat no: # 1610710.

TEKnova Tris-Hcl (0.5M, pH 6.8)- Cat no: # T1068

<u>Reagents to be prepared in lab.</u>

1. Preparation of in house 4x Lammeli SDS buffer.

| |
|---|
| 4X Lammeli SDS buffer |

| | |
|---|---|
| Glycerol | 5ml |
| SDS | 1g |
| B-Mercaptoethanol (Or 0.25 m DTT) | 2.56ml |
| 0.5M Tris-Hcl pH 6.8+0.4%SDS | 2.13ml |
| Bromophenol blue | Traces |

Aliquot 0.5ml into 1.5ml eppendorf tubes and store at -20˚C. While running gels take out 1 tube, thaw, add to lysate and discard the rest. Do not store again at -20˚C or reuse the next day.

2. Preparation of Electrophoresis Running buffer (10X, pH-8.3)

| Electrophoresis Running buffer (10X, pH-8.3) | 1 Litre | 500ml |
|---|---|---|
| Tris base | 30.3g | 15.15g |
| Glycine | 144g | 72g |
| SDS | 10g | 5g |
| DI water | 1000ml | 500ml |

Store 10X buffer at 4°C and don't adjust pH. Before running gel, dilute 100ml of 10x Electrophoresis running buffer with 900ml of DDI water to make 1X running buffer, 1L.

<u>Procedure:</u>

1. Turn on the heating block and set at 100°C.Before using, fill the heating block holes with 1ml of DI water.
2. Take the protein lysate from -20°C and keep it on ice. If the volume of lysate is 23 µl, add 7µl of 4X Lammeli SDS buffer and quick spin the lysate. If the lysate is more than

23 µl, then thaw, quick spin the lysate and aliquot 23µl from the lysate and store the rest at -20°C. Then add 7µl of 4X Lammeli SDS buffer to the 23µl lysate and quick spin.

3. Boil the lysate in the heating block containing water for 5min.
4. Meanwhile prepare 1L of 1X Electrophoresis running buffer.
5. Remove the readymade gels from the pack; remove the sealed green tape at the bottom of the gels.
6. Place the 2 gels on gel holder having protruding conduction rods and remove the comb. Place the gels in the gasketed gel holder with smaller plate facing inwards. Place the whole set up at the back of the tank, if using single gel, place the 'Buffer dam' on the other side.
7. Fill the wells first, then the space between the two gel plates and then finally fill the main tank up to 2-gel mark.
8. Quick spin the samples after boiling. Load 12 µl of protein marker in lane 1, 20µl of samples in all other lanes including controls.
9. Run the sample at 80V, 0.3A, for half an hour and then at 100v for 1hrs or until loading dye leaves the gel.

**E. Immunoblotting**

<u>Required reagents</u>

Bio-Rad Blotting Grade Blocker (Non-fat milk powder). Cat #170-6404.

Amersham Protran 0.1um Nitrocellulose membrane. Cat #10600010.

Bio-Rad filter paper. Cat #1703968.

Dysferlin antibody. NCL Hamlet (Leica Biosystems) – Lyophilized concentrated monoclonal antibody. Clone no: HAM1/7B6.

CD14 antibody. Monoclonal mouse Clone no: 134603. Cat #MAB3831.

Clarity Western ECL substrate, 200ml. Cat # 1705060.

1. 1X Towbin buffer (prepare in lab).

| Towbin transfer buffer 1X | Liter | 500ml |
|---|---|---|
| Tris base | 3.03g | 1.515g |
| Glycine | 14.4g | 7.2g |
| Methanol | 200ml | 100ml |
| DI water | 800ml | 400ml |

Store at 4°C and don't adjust pH

2. Nitrocellulose membrane 0.1µM

   Cut a membrane piece of size 8.5cm x 6.0cm using clean scissors. Hold with sterile forceps, without touching the membrane. Mark the number (corresponding to gel number on the top and bottom, left hand side of the membrane (corresponding to the gel no:) to identify the membrane when they are cut into halves. One cut membrane for a gel.

3. Filter papers

   Cut two pieces of filter papers of the size 8.5cm x 6.0cm, each. Two pieces are required for a gel.

4. 1X TBST

   For each western, dilute 100ml of 10x Stock TBST with 900ml of DI water.

   10x Stock of TBST preparation.

| TBST Buffer(10X) | 1Litre | 500ml |
|---|---|---|

| | | |
|---|---|---|
| Tris HCl | 12.11g | 6.055g |
| NaCl | 87.66g | 43.83g |
| DI water | 1000ml | 500ml |

Store at 4°C.Dont adjust pH.

Make fresh 1X TBST for each day by diluting 100ml of 10X TBST and 900ml of DI water. Add 0.5ml of Tween20 and stir on a magnetic stirrer.

5. Blocking buffer (5% nonfat dried milk powder in TBST).

   Take 50 ml of freshly prepared 1X TBST in 125 ml tissue culture bottle and add 2.5g of blotting grade blocker (nonfat dried milk powder). Drop a magnet and stir it on a magnetic stirrer until a homogenous mixture is formed without any clumps.

6. Monoclonal Dysferlin Antibody (NCL-Hamlet or Romeo)- Primary Ab (1:3000 for NCL-Hamlet; 1:1000 for Romeo) prepare fresh. 10 µl of NCL Hamlet primary antibody in 30ml 1X TBST for 1 tray. Or, 15 µl of Romeo primary antibody in 15ml 1X TBST for 1 tray
7. Monoclonal CD14 Antibody – (1:1000)- Prepare fresh. 15 µl of CD14 primary antibody in 15ml 1X TBST for 2 trays.
8. 2°Ab (1:2500 if primary NCL-Hamlet or 1:2000 if primary Romeo) 12 µl of Secondary antibody in 30ml 1XTBST (if primary NCL-Hamlet) or 15 µl of Secondary antibody in 30ml 1XTBST, for 2 trays.

<u>Procedure</u>

1. Remove gel holder containing the gels from buffer tank. Remove spacer plates using the notch remover.

2. Cut off the wells and the gel bump at the bottom of the gels using a gel scraper.

3. Equilibrate by placing the gels in 30ml 1X Towbin Transfer buffer for 15min.

4. Always hold the filter paper and membrane with clean forceps only. Soak the nitrocellulose membrane in 30ml Towbin transfer buffer for 10min in different wash box, simultaneously with the gel.

5. Soak the filter papers in Towbin transfer buffer, in a third wash box for the last 2 minutes.

6. Wipe lightly, the semi- dry transfer cell with the Towbin transfer buffer and then wipe dry.

7. Tap the filter paper on to a clean paper towel, to remove excess Towbin transfer buffer and place on the semi-dry transfer cell and roll using a sterile pipette to remove bubbles.

8. Place the membrane onto filter paper and roll gently.

9. Place the gel onto membrane and roll gently to remove bubbles.

10. Lastly, place another filter paper carefully from lower end to upper end and roll well.

11. Wipe any excess buffer near the sandwich and run the blot for 28min, 20V, and 0.5A.

12. Prepare the blocking buffer according to the protocol.

13. Remove the membrane and make sure that the ladder is transferred. We need the top blue band (250kDa) for dysferlin and 75kDa pink band for the CD14.

14. Block unreacted binding sites on the membrane by incubation with 50 ml blocking buffer for 1hr, with gentle agitation.

15. After an hour pour off the blocking buffer and wash with TBST quickly and then pour off TBST.

16. Cut the membrane into two halves, blue band corresponds to dysferlin and pink band to CD14 band, place them in separate boxes and add 30ml of dysferlin 1˚Ab for upper half of blot and CD14 1˚Ab for lower half, incubate in 4˚C fridge standing, overnight.

17. The next day, pour off the 1˚Ab, add 30ml of freshly prepared 1X TBST containing Tween 20 and keep at RT, rocking for 15min for first wash. Each blot should be washed in a separate tray.

18. For the second wash, prepare 30ml of blocking buffer for 2 trays and keep both the upper halves of the 2 blots in one tray and lower halves in a second tray. Keep at RT with gentle agitation for 30min.

19. Third and fourth washes should be done with TBST, 15min each, RT, agitation and each blot half separately.

20. During fourth wash, make 2˚Ab as per the protocol, 30ml for 2 trays. Add 15ml to each tray and keep at RT, 1hr standing without agitation.

The dysferlin membrane halves and CD14 membrane halves can be kept together while 2˚Ab incubation but must be separated while washing with 1XTBST.

| Wash box 1-Dysferlin | Wash box 2- CD14 |
|---|---|
| Membrane 1 - top half<br>Membrane 2- top half | Membrane 1 - bottom half<br>Membrane 2- bottom half |

21. Pour off 2˚Ab and wash with 1XTBST- 4times, 15min each, rocking.

22. After the fourth wash, while blot is in TBST, prepare ECL by mixing 1 ml Peroxide reagent and 1 ml enhancer reagent in a light sensitive 15ml centrifuge tube. Pour off TBST and add Biorad ECL evenly on the membrane, swirl immediately well to mix and keep it for 2 min incubation (or 3 min).

23. With forceps and wiping excess ECL on the sides of the box, place both the halves of the membrane together onto a transparent folder kept inside the cassette such that the halves align well, leaving a space between the two halves.

24. Close the cassette and take it to dark room.

**F. Developing film**

1. In the dark room, switch off the lights, take out a film, fold it on the top right hand corner.

2. Place the X-ray sheet on to the transparent sheet with blot inside and close the cassette, and expose for 2min and then drop it to developer. Similarly, expose for 1min and 30sec and then drop it to developer.
3. In case, the CD14 bands or Dysferlin bands are not clear, keep the transparent sheet with blot inside, on benchtop for 30min or more.
4. Take it to dark room and expose for 2min and drop into the developer. Similarly, expose for 1min, 30sec, 10sec, 5sec and 3sec and develop it.

**G. Scanning the images and saving the data**

1. Scan all the films in the Konica scanner machine

2. Place the film upside down, with the right hand to corner fold now in the left side top corner of the scanner surface.

3. Scan settings are as follows:

- Resolution – highest resolution – 600x600dpi- press ok.
- Color- gray scale- press ok.
- File type- JPEG- press ok.

4. Press Scan and choose the direct input mode, enter your email address and press ok.

**H. <u>Quantification</u>**

1. Use image studio to compare full-length dysferlin signal strength against housekeeping protein, CD14 signal for each sample.

2. Ratios for each samples are derived by comparing the dysferlin signal to the same exposure CD14 signal, for example, 5sec Dysferlin (DYSF) signal to 5sec CD14 signal, 10sec DYSF to 10 sec CD14 signals, 30sec DYSF to 30 sec CD14 signals. Likewise, any best DYSF signals can be compared to the CD14 signal values of the same exposure time, which are not weak or unsaturated value. The ratio of individual sample is divided by ratio value of positive control to derive the percentage of DYSF for each patient sample. The percentage values of all exposures of each patient are averaged to get a final value of %DYSF.

3. Choosing the positive control – The positive control sample should be a sample of a normal sequenced (without any DYSF variant) individual without any history of any neuromuscular disease that shows visually and quantitatively raw %DYSF/CD14 ratio of $\geq$ 80%.

4. Choosing the negative control - The negative control sample should be a previous sample of a sequenced (with confirmed DYSF pathogenic variant(s)) patient who cannot produce any DYSF protein (0%) with visual and quantitative confirmation.

**Whole Blood Targeted RNA-Seq Library Preparation and Sequencing**

High quality (RNA Integrity Number; RIN>7) RNA was extracted from whole blood of the patients and control individuals using QIAamp RNA Blood Kit (cat # 52304, Qiagen) following the manufacturer's protocol. Only blood specimens in EDTA tubes shipped to us within 24hrs from blood draw time based on time log on top of the EDTA vial were used for RNA extraction to control for any RNA degradation effect. Library preparation was performed using SureSelect$^{XT}$ RNA Target Enrichment for Illumina Multiplexed Sequencing kit (cat# G9691-9000) following manufacturer's protocol. Targeted RNA-Seq was performed to have a more focused clinically relevant platform for neuromuscular disease (NMD) diagnostics and to achieve greater read depth and coverage of the target NMD genes. We used a custom-designed target library probe to capture 274 genes (Table S1) that are known to be NMD-associated and are known to have skeletal muscle expression (≥1TPM) as retrieved from The Genotype-Tissue Expression (GTEx) portal (https://gtexportal.org/home/)[4]. These 274 genes were curated initially based on the associations to NMDs (Types of Neuromuscular Diseases, http://muscle.ca/wp-content/uploads/2019/08/Disorder_List_ENG_May2017.pdf) as recently done by us[5]. Strand-specific paired-end 150bp sequencing was performed on an Illumina NextSeq instrument to obtain high output paired-end by 150bp reads at a depth of more than 15 million reads per sample.

**Targeted RNA-Seq Tiered Analysis Approach**

We used a novel tiered approach of targeted RNA-Seq analysis for molecular diagnosis which included Tiers 1, 2 and 3 (Fig. 1C). Taking together the results of RNA-seq analysis, available clinical information, %DYSF protein expression in CD14+ monocytes, and DNA-Seq data, we performed phenotype-genotype correlations. In our Tier 1, we analyzed for aberrant splicing including exon(s) skip(s), pseudoexon gain, exon extension or exonic splice gain. We performed RNA variant calling in Tier 1 including confirmation of specific variants found previously in DNA-Seq and identification of novel reportable and causal variants not found by DNA-Seq. Moreover, in Tier 1, we analyzed for the presence of allele expression imbalance (AEI) due to nonsense mediated decay, nonstop mediated decay, or allele-specific expression. In Tier 2, we analyzed for differential isoform patterns or abundance including differential exon usage by percent spliced in (PSI). In Tier 3, we first investigated *DYSF* mRNA expression levels and compared to normal controls, and then if needed expanded expression analysis to the rest of the panel. The results were clinically correlated to reclassify VUSs, identify pathogenic events at the mRNA level, and understand the pathogenic nature of the variants as per ACMG-AMP guidelines, in order to submit to public databases such as ClinVar (https://www.ncbi.nlm.nih.gov/clinvar/), Human Genome Mutation Database (HGMD: http://www.hgmd.cf.ac.uk/ac/index.php), Human Genome Variation Society (HGVS: https://www.hgvs.org/) and others. All RNA-seq assay results were reported back to patients and/or respective physicians as research reports according to guidelines of the approved Institutional Review Board protocol.

Previous literature using RNA-Seq to diagnose neuromuscular disorders analyzed muscle biopsies and used the publicly available GTEx data as controls for comparison[6,7]. Although GTEx samples are an important resource for comparison, their use as either proxy tissue or as normal controls in RNA-Seq is debatable due to a) potential differences in the methods pipeline and sequencing platform settings, b) possible sample differences in sex, age, and storage conditions since most GTEx sample are from individuals >40 years age[8]. Therefore, to make our pipeline more clinically cautious and relevant, and to improve analyses, we used internal normal control blood specimens. These samples came from individuals of different ethnicities without any symptoms or individual/family history of neuromuscular or neurological disease, showed ≥100% DYSF protein expression in CD14+ monocytes, and underwent the same sample collection/storage conditions, sequencing platform and overall methods pipeline as patient samples.

**RNA-Seq Alignment and Quality Control**

Raw FASTQ files were checked for quality using FastQC (https://www.bioinformatics.babraham.ac.uk/projects/fastqc/)[9]. Reads were not trimmed beyond removal of adapter sequences [10] using Trimmommatic (http://www.usadellab.org/cms/?page=trimmomatic)[9,11] to prepare for alignment. Human reference genome GRCh38 (NCBI) (https://www.ncbi.nlm.nih.gov/assembly/GCF_000001405.39) and NCBI *Homo sapiens* Annotation Release 106 (https://www.ncbi.nlm.nih.gov/genome/annotation_euk/Homo_sapiens/106/) were obtained from Illumina iGenomes (https://support.illumina.com/sequencing/sequencing_software/igenome.html) and sequenced reads were aligned using the splice-aware alignment program STAR version 2.5.2b (https://code.google.com/archive/p/rna-star/ https://github.com/alexdobin/STAR) in 2-pass mode [12] to improve novel splice junction discovery.

Quality metrics for all samples were obtained by running QoRTs v1.2.42 (http://hartleys.github.io/QoRTs/)[13], and principal component analysis (PCA) on gene expression was performed to check for outlier status based on tissue composition or contamination. We used PCA of the 274 gene expression of the patient samples for quality control which showed better clustering towards whole blood RNA-Seq compared to lower quality and coverage, and 3'-bias for RNA-Seq using CD14+ monocytes or PBMCs (data not shown). Uniquely mapped, non-duplicate read counts for genes and splice junctions were obtained by removing duplicate reads from STAR-aligned BAM files using Picard MarkDuplicates (http://broadinstitute.github.io/picard), converting to FASTQ files, and realigning with STAR using the same parameters as before.

**RNA-Seq Variant Calling**

To confirm RNA expression of variants identified by DNA-Seq and check for sequence variants not reported from DNA, we followed the protocol outlined in GATK Best Practices for Variant Calling in RNA (https://software.broadinstitute.org/gatk/documentation/article?id=4067)[14-16]. Filtered VCF files were annotated using ANNOVAR (http://annovar.openbioinformatics.org/en/latest/)[17] and high quality variants in *DYSF* were extracted for evaluation using ACMG-AMP Guidelines for Variant Interpretation[18]. High quality loss of function (LOF) variants and variants with an allele frequency of <5% in the Genome Aggregation Database (gnomAD: https://gnomad.broadinstitute.org/) [19] were also pulled for evaluation from the remaining 273 genes in the panel. All potentially causative variants were manually evaluated using the Integrative Genomics Viewer (IGV: https://software.broadinstitute.org/software/igv/) [20] to ensure they were not a result of mis-mapping or noise. We also used *in silico* prediction algorithms, namely Polyphen2 (Polymorphism Phenotyping v2: http://genetics.bwh.harvard.edu/pph2/)[21,22], SIFT (Sorting

Intolerant From Tolerant: https://sift.bii.a-star.edu.sg/)[23-25], MutationTaster (http://www.mutationtaster.org/)[26], FATHMM (Functional Analysis through Hidden Markov Models: http://fathmm.biocompute.org.uk/)[27], and Transcript-inferred Pathogenicity (TraP: http://trap-score.org/)[25] score for further tentative understanding of the variant pathogenicity.

**RNA-Seq Splicing Analysis**

Counts for splice junctions (annotated and unannotated) that overlapped *DYSF* were extracted from STAR output files from all samples. Splice events were considered "annotated" if they matched a known transcript. "Unannotated" events were further analyzed to see if either junction matched an exon/intron boundary in a known transcript. Events fitting this criterion were only kept for analysis if they had read support totaling more than 5% than that of the matched junction. For events where neither junction matched a known exon/intron boundary, they were still considered for further analysis if the read support was >10% than that of the matched junction. The filtered group of unannotated splice events was further curated by the number of samples each was observed in. If an event had >5% read support in more than half of our control samples, it was automatically eliminated as a potential pathogenic event. This extremely conservative cutoff was chosen because the goal at this stage was merely to narrow the list of splice events undergoing manual evaluation in later steps. Each of the unannotated splice events from the curated list was observed using the Integrative Genomics Viewer, prioritizing the events occurring in just one sample. This led to the identification of 38 pathogenic aberrant splicing events, for which 27 causative variants had been reported from DNA sequencing either as a pathogenic or likely pathogenic variant or VUS or not found in DNA testing but found by RNA-

Seq. Expanding the analysis to events in a handful of samples found an additional 3 pathogenic splicing events.

**RNA-Seq Allele Expression Imbalance (AEI)**

To evaluate allele expression across *DYSF*, the allele ratio for each individual high confidence single nucleotide variant (SNV) in each sample was calculated by dividing the read count of the lesser-expressed nucleotide (lesser allele) by the total number of reads at the variant position. Read counts for SNVs passing all filters were obtained from the RNA-Seq variant calling VCF file. Each SNV was grouped by the number of PTVs in the sample it belonged to. Because the data did not pass tests for normality or homogeneity of variance, significant differences between groups were calculated using Wilcoxon rank sum tests and p-values were adjusted using the Benjamini-Hochberg method.

**Allele Expression Imbalance (AEI) Calculation Method**

Only exonic heterozygous SNVs in DYSF located in constitutively expressed exons, called at >50X per allele and passing all variant quality filters, were considered in the analysis. Samples that were observed to be outliers by gene expression PCA or did not contain any heterozygous SNVs in DYSF were excluded from AEI analysis. Further criteria for sample inclusion was a requirement that the sample contained at least two heterozygous SNVs located more than 150 coding bases apart, to show that the observed AEI is consistent across the entire length of the transcript and that any effect seen is not local to any single variant. A total of 50 samples including 6 controls met the criteria for inclusion. In each sample, allele ratios were calculated for every SNV meeting the stated criteria by taking the lower number of allele-supporting reads divided by the total available reads at that site for Allele A and the greater number of allele-supporting reads divided by the total site reads for Allele B. In this manner, allele expression is divorced from the concept of "reference" or "alternate" allele. Both AEI and overall gene abundance were correlated with the observation of PTVs in a sample and we attempted to keep the gene abundance observation in the visualization of AEI. Variant call depth is not normalized and varied widely within individuals since depth of coverage is not consistent across all exons, so the calculated allele ratios were instead applied to the overall gene TPM for plotting as a more stable representative of abundance. Each sample is represented by two plotted points showing the average of Allele A and B connected by a line. Error bars represent one standard deviation.

**RNA-Seq Gene Expression Analysis**

Non-duplicate read counts for all genes in the panel were obtained from STAR output files. Transcripts Per Million (TPM) Normalization was performed to control for sequencing depth and to make samples directly comparable. Comparisons of gene expression were performed using Welch's t-test followed by pairwise t-tests with non-pooled SD. P-values were adjusted using the Benjamini-Hochberg method.

**Whole Genome Sequencing (WGS or GS) and Analysis**

When needed, WGS/GS was performed in a CLIA-CAP certified facility as performed by us previously[28]. Subsequent WGS was performed to validate the RNA-Seq results and novel findings. The analyzed region of genes included the coding exons and intronic region on both sides of each exon. Select pathogenic deep intronic sites were also targeted. In some cases, due to the complexity of the sequence, not all variants in the flanking intronic sequence were able to be analyzed. Variants were evaluated by their reported frequency in databases such as the Genome Aggregation Database (gnomAD), Human Gene Mutation Database (HGMD), and ClinVar. Variants that have a population frequency greater than expected given the prevalence of the disease in the general population were considered to be benign variants. Silent variants of uncertain significance and intronic variants of uncertain significance beyond +/-3 were not reported back unless known to be pathogenic or other evidence suggests potential disruption of splicing. The interpretation of variants was based on the individual's clinical phenotype. On average % fully covered disease causative gene target bases was 99.5%; average % fully covered disease causative gene exons was 98.8%; and average coverage per target base was 30.6. WGS was performed on genomic DNA using 2X150bp reads on Illumina NovaSeq 6000 System at a mean coverage of 30X in the target region. The target region includes coding exons and 10bp of flanking intronic sequence of the known protein-coding RefSeq genes. This sequencing provides >97% coverage of the 22,000 genes in the genome at >30X. This includes 100% coverage (>20X) of all exons of 3000 disease-associated genes. A base was considered to have sufficient coverage at 20X and an exon is considered fully covered if all coding bases plus three nucleotides of flanking sequence on either side were covered at 20X or more. Low coverage regions, if any, were limited to ~1% or less of the nucleotides included in this panel unless a pathogenic variant explaining the phenotype was discovered. Alignment to the human reference genome (hg19 https://www.ncbi.nlm.nih.gov/assembly/GCF_000001405.13/) was performed and annotated variants were identified in the targeted region. Variants were called at a minimum coverage of 8X and an alternate allele frequency of 20% or higher. High-quality single nucleotide variants (SNVs), which pass quality filters, were not confirmed by Sanger sequencing. Reportable SNVs that do not pass the quality filters were confirmed using Sanger sequence analysis. This assay could not detect variants in areas containing large numbers of tandem repeats. Copy number variation (CNV) and absence of heterozygosity (AOH) analysis was assessed using BioDiscovery NxClinical 4.3 (https://www.biodiscovery.com/products/NxClinical) (BioDiscovery, El Segundo, CA). All variants were annotated and classified based on ACMG-AMP guidelines[18].

**Neuromuscular Disease (NMD) 131 Gene-Panel used for NGS DNA sequencing**

*ACTA1* (NM_001100.3), *AGRN* (NM_198576.3), *ALG14* (NM_144988.3), *ALG2* (NM_033087.3), *ANO5* (NM_213599.2), *ATP2A1* (NM_173201.3), *B3GALNT2* (NM_152490.4), *B4GAT1* (NM_006876.2), *BAG3* (NM_004281.3), *BICD2* (NM_001003800.1), *BIN1* (NM_139343.2), *CACNA1S* (NM_000069.2), *CAPN3* (NM_000070.2), *CAV3* (NM_033337.2), *CCDC78* (NM_001031737.2), *CFL2* (NM_021914.7), *CHAT* (NM_020549.4), *CHKB* (NM_005198.4), *CHRNA1* (NM_000079.3), *CHRNB1* (NM_000747.2), *CHRND* (NM_000751.2), *CHRNE* (NM_000080.3), *CLCN1* (NM_000083.2), *CNTN1* (NM_001843.3), *COL12A1* (NM_004370.5), *COL6A1* (NM_001848.2), *COL6A2* (NM_001849.3), *COL6A3* (NM_004369.3), *COLQ* (NM_005677.3), *CPT2* (NM_000098.2), *CRYAB* (NM_001885.2), *DAG1* (NM_004393.5), *DES* (NM_001927.3), *DMD* (NM_004006.2), *DNAJB6* (NM_058246.3), *DNM2* (NM_001005360.2), *DOK7* (NM_173660.4), *DPAGT1* (NM_001382.3), *DPM1* (NM_003859.1), *DPM2* (NM_003863.3), *DPM3* (NM_153741.1), *DYNC1H1* (NM_001376.4), *DYSF* (NM_003494.3), *EMD* (NM_000117.2), *FHL1* (NM_001449.4), *FKBP14* (NM_017946.3), *FKRP* (NM_024301.4), *FKTN* (NM_001079802.1),

*FLNC* (NM_001458.4), *GAA* (NM_000152.3), *GFPT1* (NM_002056.3), *GMPPB* (NM_013334.3), *GNE* (NM_001128227.2), *HNRNPA2B1* (NM_031243.2), *HNRNPDL* (NM_031372.3), *IGHMBP2* (NM_002180.2), *ISPD* (NM_001101426.3), *ITGA7* (NM_002206.2), *KBTBD13* (NM_001101362.2), *KCNJ2* (NM_000891.2), *KLHL40* (NM_152393.3), *KLHL41* (NM_006063.2), *LAMA2* (NM_000426.3), *LAMB2* (NM_002292.3), *LAMP2* (NM_002294.2), *LARGE1* (NM_004737.4), *LDB3* (NM_001080116.1), *LIMS2* (NM_001136037.2), *LMNA* (NM_170707.3), *LMOD3* (NM_198271.4), *LRP4* (NM_002334.3), *MATR3* (NM_199189.2), *MEGF10* (NM_032446.2), *MTM1* (NM_000252.2), *MUSK* (NM_005592.3), *MYF6* (NM_002469.2), *MYH2* (NM_017534.5), *MYH7* (NM_000257.3), *MYL2* (NM_000432.3), *MYOT* (NM_006790.2), *MYPN* (NM_032578.3), *NEB* (NM_001271208.1), *PHKA1* (NM_002637.3), *PLEC* (NM_000445.4), *PLEKHG5* (NM_020631.4), *PMP22* (NM_000304.3), *PNPLA2* (NM_020376.3), *POMGNT1* (NM_017739.3), *POMGNT2* (NM_032806.5), *POMK* (NM_032237.4), *POMT1* (NM_007171.3), *POMT2* (NM_013382.5), *PREPL* (NM_006036.4), *PYGM* (NM_005609.2), *RAPSN* (NM_005055.4), *RYR1* (NM_000540.2), *SCN4A* (NM_000334.4), *SELENON* (NM_020451.2), *SGCA* (NM_000023.2), *SGCB* (NM_000232.4), *SGCD* (NM_000337.5), *SGCG* (NM_000231.2), *SIL1* (NM_022464.4), *SMCHD1* (NM_015295.2), *SNAP25* (NM_003081.3), *SQSTM1* (NM_003900.4), *STAC3* (NM_145064.2), *STIM1* (NM_003156.3), *SUN1* (NM_001130965.2), *SUN2* (NM_015374.2), *SYNE1* (NM_033071.3), *SYNE2* (NM_182914.2), *TAZ* (NM_000116.4), *TCAP* (NM_003673.3), *TIA1* (NM_022173.2), *TMEM43* (NM_024334.2), *TMEM5* (NM_014254.2), *TNNI2* (NM_003282.3), *TNNT1* (NM_003283.5), *TNPO3* (NM_012470.3), *TOR1AIP1* (NM_001267578.1), *TPM2* (NM_003289.3), *TPM3* (NM_152263.3), *TRAPPC11* (NM_021942.5), *TRIM32* (NM_012210.3), *TRPV4*

(NM_021625.4), *TTN* (NM_133378.4), *UBA1* (NM_003334.3), *VCP* (NM_007126.3), *VMA21* (NM_001017980.3), *VRK1* (NM_003384.2).

**Neuromuscular Disease (NMD) Panel DNA-Sequencing with Deletion-Duplication Analysis**

When needed, NMD 131 gene-panel used for NGS DNA sequencing was performed to validate the RNA-Seq results and novel findings. Direct sequencing of the amplified captured regions was performed using 2X100bp reads on Illumina systems. A base was considered to have sufficient coverage at 20X and an exon was considered fully covered if all coding bases plus three nucleotides of flanking sequence on either side are covered at 20X or more. Low coverage regions, if any, were limited to ~1% or less of the nucleotides in the test unless a pathogenic variant explaining the phenotype is discovered. Copy number variation (CNV) and absence of heterozygosity (AOH) analysis was assessed using BioDiscovery NxClinical 4.3 (BioDiscovery, El Segundo, CA). CNV analysis was designed to detect deletions and duplications of three exons or more; in some instances, due to the size of the exons or other factors, not all CNVs may be analyzed. This assay was not designed to detect small copy number variants (<200 Kb) unless there was a significant copy number change (homozygous deletion or duplication ≥4X). Genomic coordinate numbering was based on GRCh37/hg19. All variants were annotated and classified based on ACMG-AMP guidelines[18].

**Sanger Confirmation of Genomic DNA**

When needed, PCR-amplified genomic DNA and RT-PCR amplified cDNA products of the *DYSF* gene were Sanger sequenced for confirmation of the variants. An automated primer design script, developed and validated in-house, was used to design primers[29].

#### SUPPLEMENTARY FIGURES

**Figure S1 (A-D). Sashimi and/or IGV plots of cases with new *DYSF* splicing mechanisms identified due to previously reported variant with prior pathogenic (P) or likely pathogenic (LP) classification confirming molecular diagnosis (also see Table S5 for reference and details of each patient case)**

**A) Patient A1**

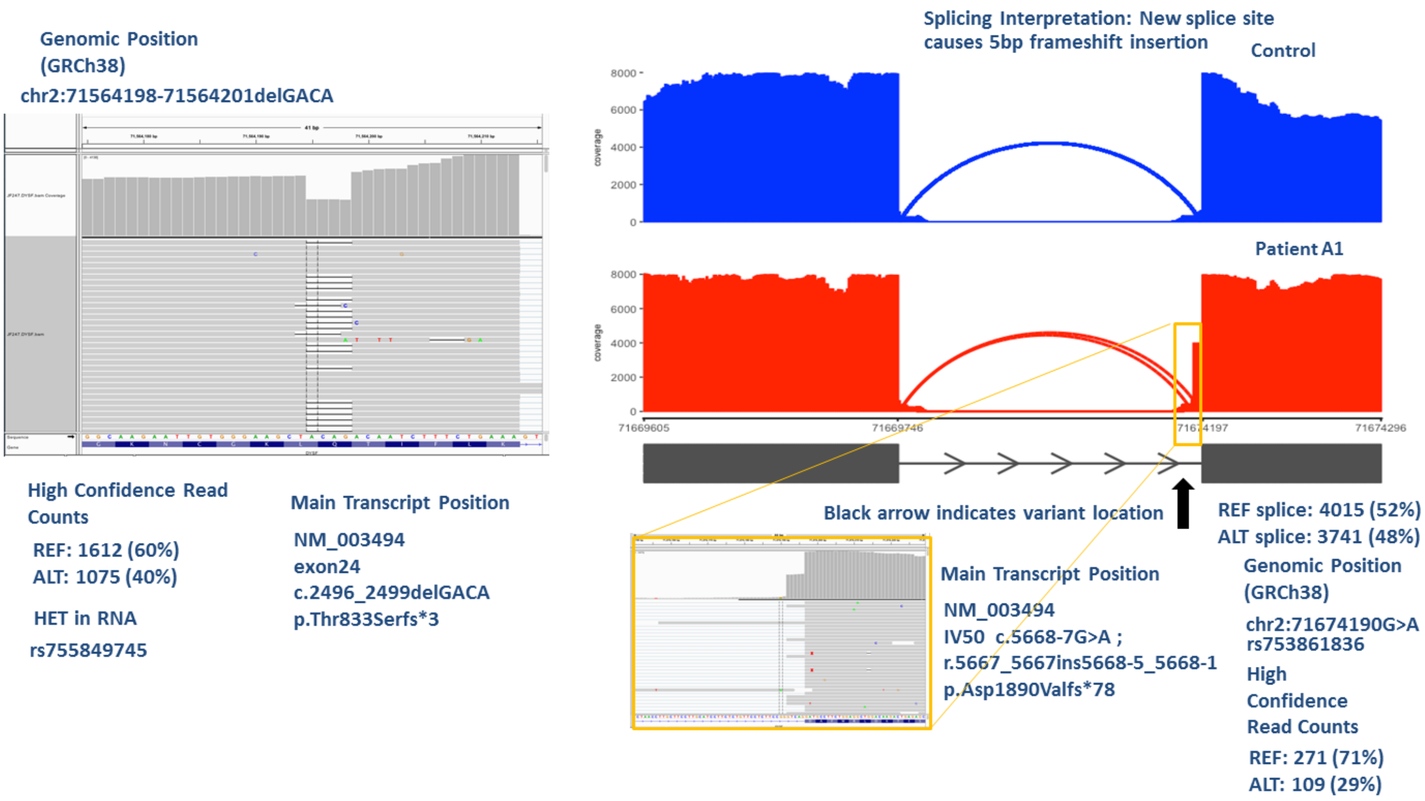

**(A) Patient A1:** Confirmation of heterozygous c.2496_2499delGACA in RNA (left). Right: IVS50: c.5668-7G>A (likely pathogenic) causes creation of new splice acceptor site in IVS50 causing 5bp exon extension resulting in insertion at the RNA level of r.5667_5668ins5668-5_5668-1 that in turn causes a frameshift and a downstream stop codon (p.Asp1890Valfs*78). DYSF mRNA expression significantly reduced (p=1.5e-05). Phasing of the second pathogenic variants in this case (c.2496_2499delGACA) is not possible using the AEI value.

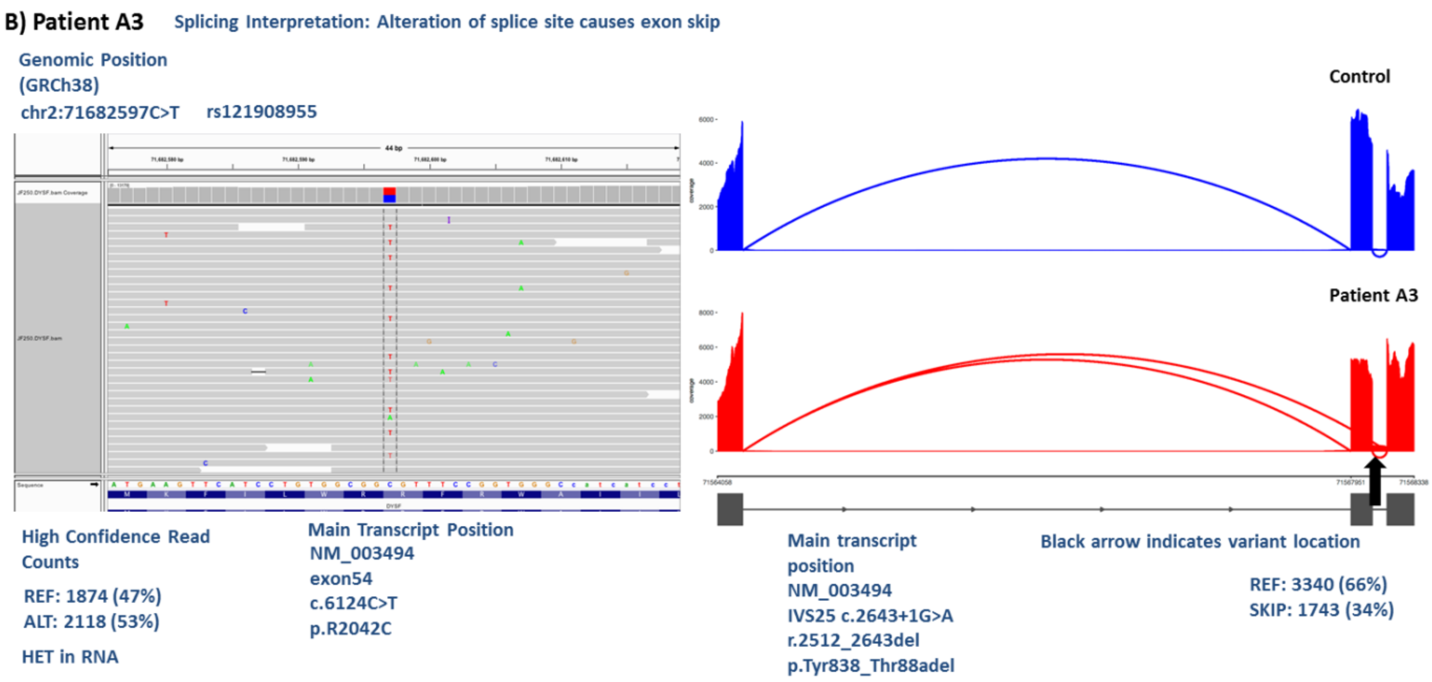

**(B) Patient A3:** IVS25 variant (c.2643+1G>A) results in an exon 25 skip causing an effect at the RNA level of r.2512_2643del (right). This in turn causes an in-frame deletion at the protein level of p.Tyr838_Thr881del. The exon 54 variant (left) in this case does not cause a splice defect or large AEI, but the ratio of AEI is different than that for the IVS25 variant causing exon 25 skip. The AEI ratio (47%:53%) seen in case of the exon 54 variant is not exactly reciprocal to that of the other variant or the unskipped:skipped ratio, since it is possible that some of the reads are captured before nonsense mediated decay or decay due to instability has occurred in them. Also, overall DYSF mRNA expression is reduced compared to controls. Taken together, Dysferlinopathy diagnosis for this case is confirmed and the pathogenic mechanism is understood to make patient clinical trial-ready.

**C) Patient A7: Cryptic Pseudoexon Ex50.1 insertion**

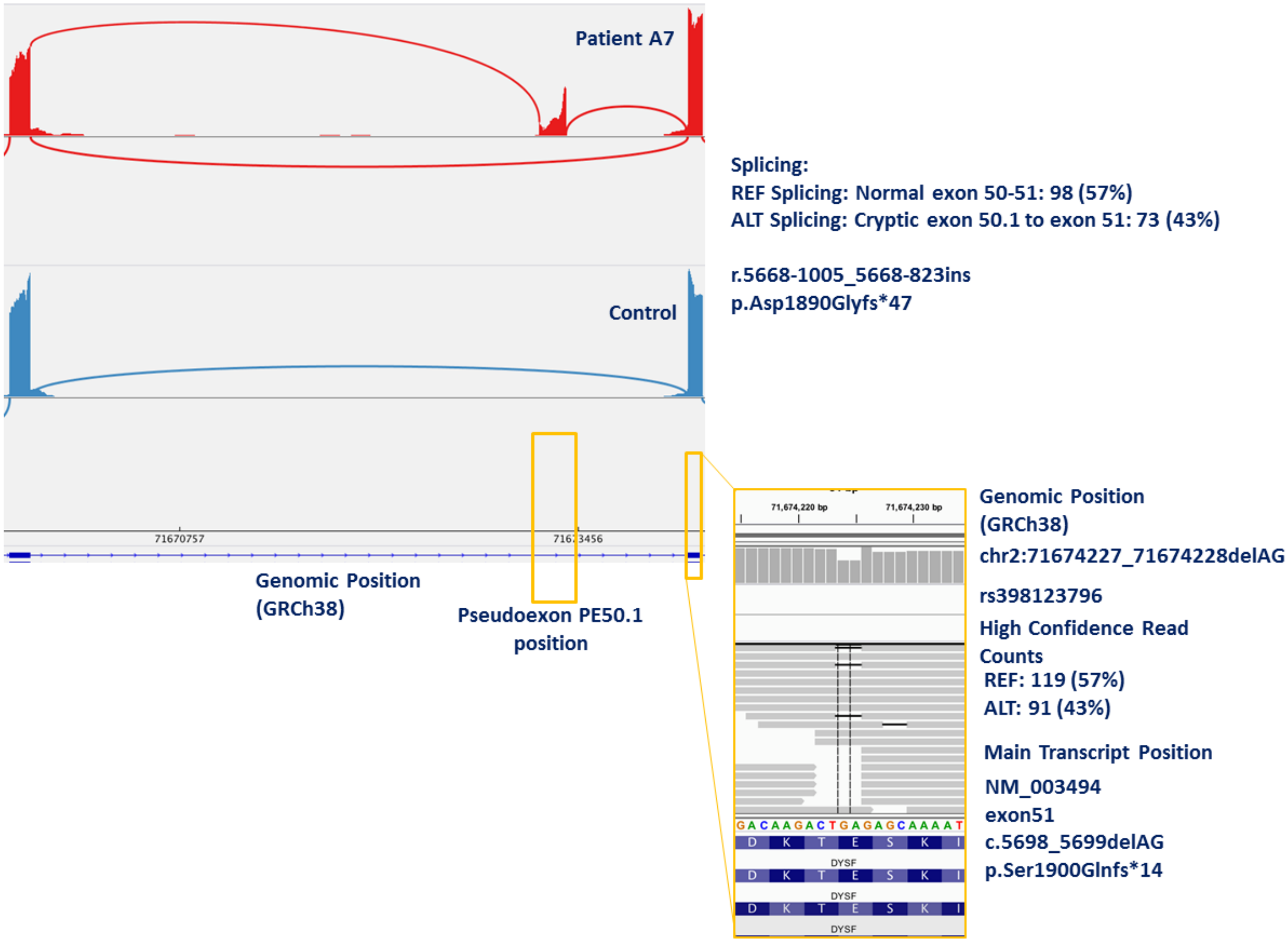

**(C) Patient A7:** A pseudoexon is created between exon 50 and exon 51 due to the deep-intronic IVS50 variant: c.5668-824C>T (top left). This variant results in the creation of a new splice acceptor site in IVS50 causing pseudoexon insertion in 43% of transcripts and, normal exon50-51 in 57% of transcripts. Phasing of the IVS50 and exon 51 variants (bottom right) is confirmed to be *in trans* based on the presence of two pathogenic variants and pathogenic splicing effects along with clinical data and disease-range %DYSF. Dysferlinopathy diagnosis is confirmed.

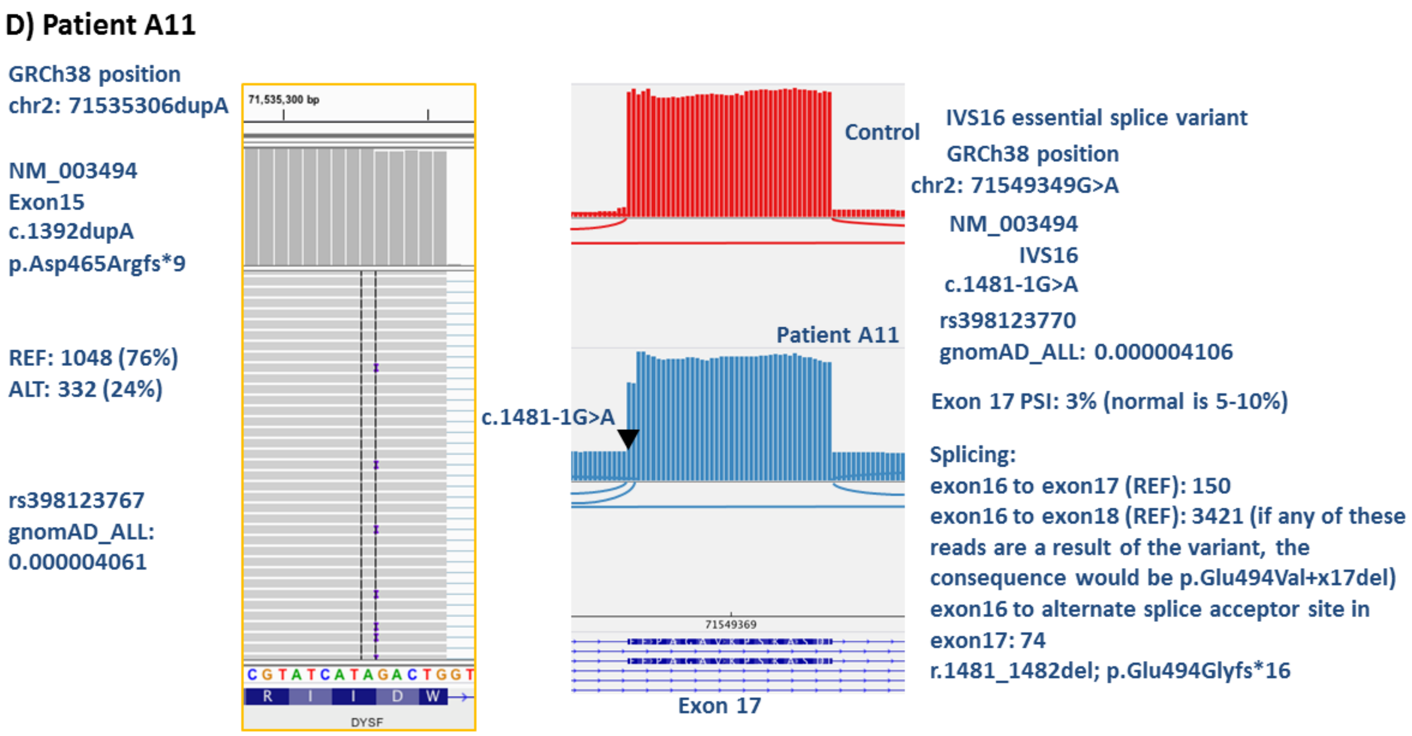

**(D) Patient A11:** RNA analysis shows that the c.1481-1G>A splice variant (right) causes two events to occur: (a) 2 bp deletion in exon 17 occurs during splicing due to the use of an alternate splicing acceptor site in exon 17 (r.1481_1482del) which causes a glutamic acid at position 494 to change to glycine and a downstream frameshift and premature stop codon (p.Glu494Glyfs*16), and (b) likely also leads to complete skip of exon 17 (p.Glu494Val+exon17del) in muscle tissue where exon 17 is present in the RNA transcript; however the skip of exon 17 was not seen in the analysis of the blood RNA-Seq due to the lack of exon 17 of *DYSF* in the majority of blood RNA transcripts. Exon 17 PSI in this case was 3% (normal is 5-10%). Reduced exon 17 PSI indicates this variant is *in trans* with the other *DYSF* variant found in this case (c.1392dupA: left IGV image). Since exon 17 is mostly skipped in healthy conditions in blood, the effect of the IVS16 variant is not prominent in blood but will affect skeletal muscle through the 2 bp frameshift deletion identified.

**Figure S2 (A-Q). Sashimi and/or IGV plots of cases in which splicing events were identified that are related to a single pathogenic event or reclassification of VUS or discovery of a single pathogenic mechanism in *DYSF* that completes the molecular diagnosis (also see Table S5 for reference and details of each patient case)**

**A)-C) Patients B1, B5, B6**

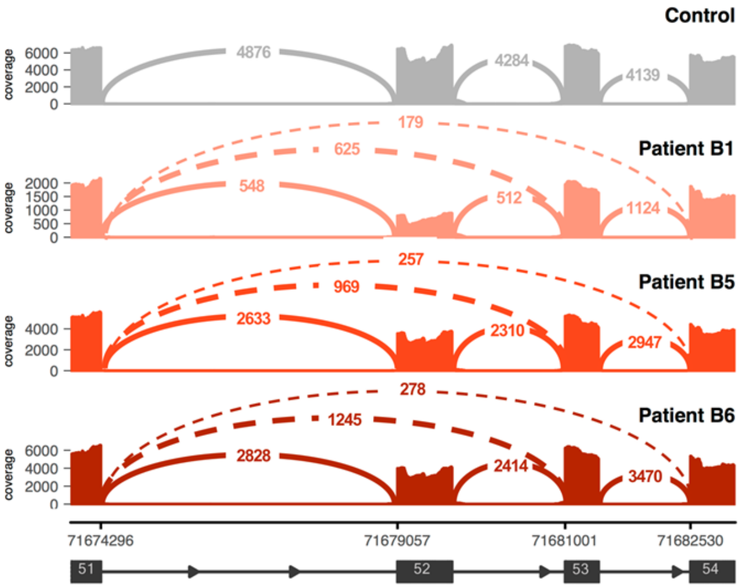

| Patient ID | Normal Splicing : Aberrant Splicing AEI | $2^{nd}$ Variant AEI | Phase |
|---|---|---|---|
| B1 | Normal splicing: 40% Exon 52 skip: 47% Exons 52 & 53 skip: 13% | REF: 67%, ALT: 33% Exon 20 c.1834C>T p.Gln612* | *In trans* |
| B5 | Normal splicing: 68% Exon 52 skip: 25% Exons 52 & 53 skip: 7% | REF: 32% ALT: 68% Exon 19 c.1663C>T p.Arg555Trp | *In trans* |
| B6 | Normal splicing: 64% Exon 52 skip: 29% Exons 52 & 53 skip: 7% | REF: 29%, ALT: 71% Exon 27 c.1663C>T p.Arg959Trp | *In trans* |

Sashimi plots of exon-skipping events seen in three patients (Patients B1, B5 and B6- also see Figure. 3A in main text). Subsequent whole genome sequencing (WGS) or CNV analysis identified the cause to be varying sized large deletions encompassing exon 52. Numbers within continuous and dashed lines indicate number of spliced transcripts.

**(A)** For **patient B1**, normal splicing is only in 40% of the transcripts. Subsequent WGS after RNAseq identified an exon 52 gross deletion: c.5768-51_5946+50del (279 bp). This newly identified deletion variant is classified as pathogenic. The exon 52 deletion variant is *in trans* with the exon 20 nonsense pathogenic variant based on AEI ratios. Significantly reduced DYSF mRNA expression is seen. Dysferlinopathy diagnosis is confirmed.

**(B)** A similar splicing effect in **patient B5** due to a different gross deletion of 2638 bp spanning exon 52 (chr2:71,904,958-71,907,595; hg19) was subsequently identified by CNV analysis. This newly identified deletion variant is classified as pathogenic. The exon 52 deletion variant is *in trans* with the exon 19 pathogenic variant based on AEI ratios. Dysferlinopathy diagnosis is confirmed.

**(C)** A similar splicing effect in **patient B6** due to a different gross deletion of 3673 bp spanning exon 52 (chr2:71,903,191-71,906,863; hg19) was subsequently identified by CNV analysis. This newly identified deletion variant is classified as pathogenic. The exon 52 deletion variant is *in trans* with the exon 27 pathogenic variant based on AEI ratios. Significantly reduced *DYSF* mRNA expression is seen. Dysferlinopathy diagnosis is confirmed.

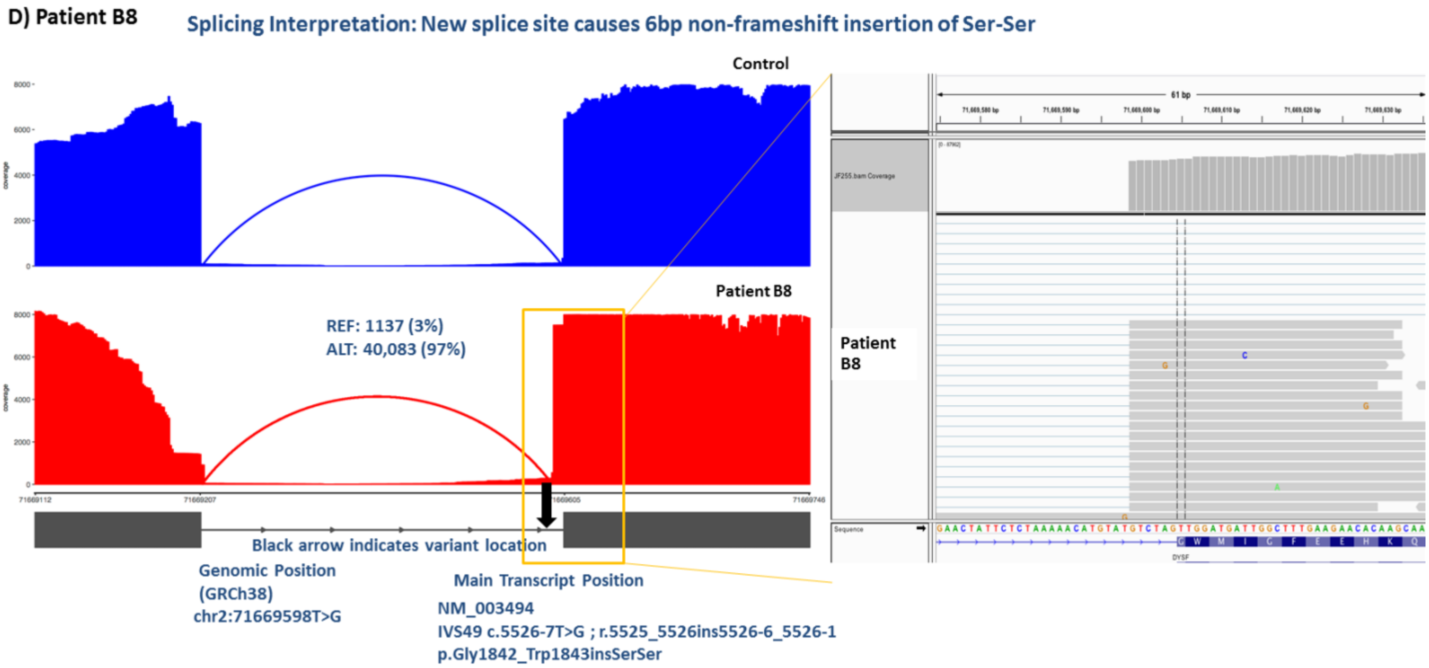

**(D) Patient B8:** IVS49: c.5526-7T>G was previously classified as a VUS and is now reclassified as pathogenic based on the Sashimi plot showing the creation of a new splice acceptor site in IVS49. This causes a 6bp exon extension resulting in an insertion at the RNA level (r.5525_5526ins5526-6_5526-1) and an in-frame insertion of two Serines (p.Gly1842_Trp1843insSerSer). No significant reduction in *DYSF* mRNA expression (p=0.26) is seen meaning the IVS49 homozygous variant is pathogenic at the protein level due to Serine insertions.

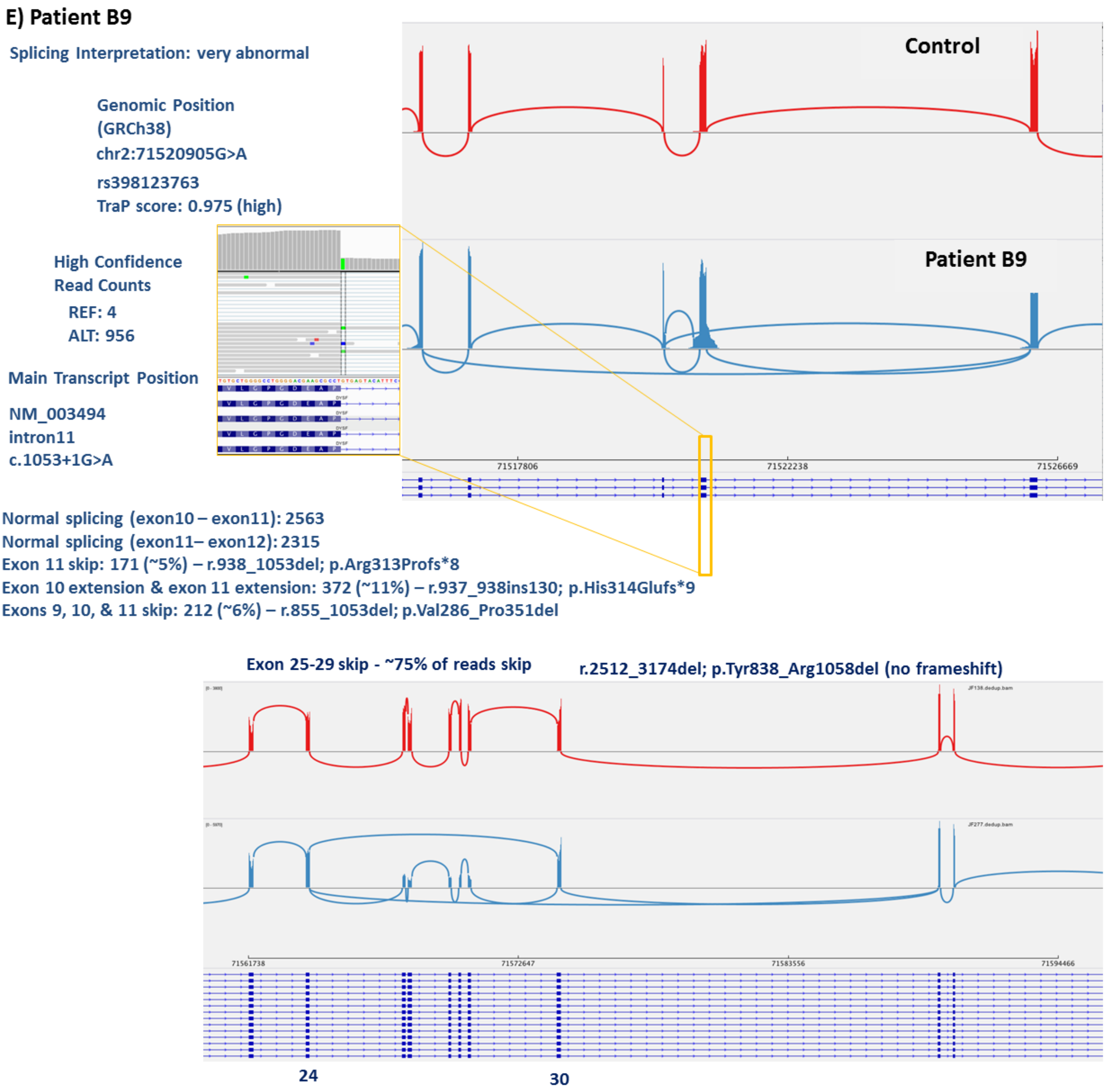

**(E) Patient B9:** IVS11: c.1053+1G>A was previously classified with conflicting interpretations (likely pathogenic/pathogenic). The IVS11 variant causes multiple splicing aberrations (top panel): an exon 11 skip that causes a frameshift and stop codon in 5% of the transcripts (r.938_1053del; p.Arg313Profs*8), an exon 10 and exon 11 extension in 11% of the transcripts causing a frameshift and stop codon (r.937_938ins130; p.His314Glufs*9), and an exon 9, exon 10 and exon 11 combined skip in 6% of the transcripts causing an in-frame deletion (r.855_1053del; p.Val286_Pro351del). Based on this data, the IVS11 variant is now reclassified with confirmation as pathogenic. Also, we identified 75% of the transcripts with a complete skip of exons 25-29 (bottom panel). Subsequent neuromuscular disease panel DNA analysis with deletion/duplication analysis after RNAseq identified an Exon 25-Exon 29 gross deletion of 2650 bp at location Chr2:71795161-71797811, Cytoband: 2p12.3 . This newly identified deletion variant is classified as pathogenic. The exon 25-exon 29 deletion variant is *in trans* with the IVS11 variant based on AEI ratios. Dysferlinopathy diagnosis is confirmed.

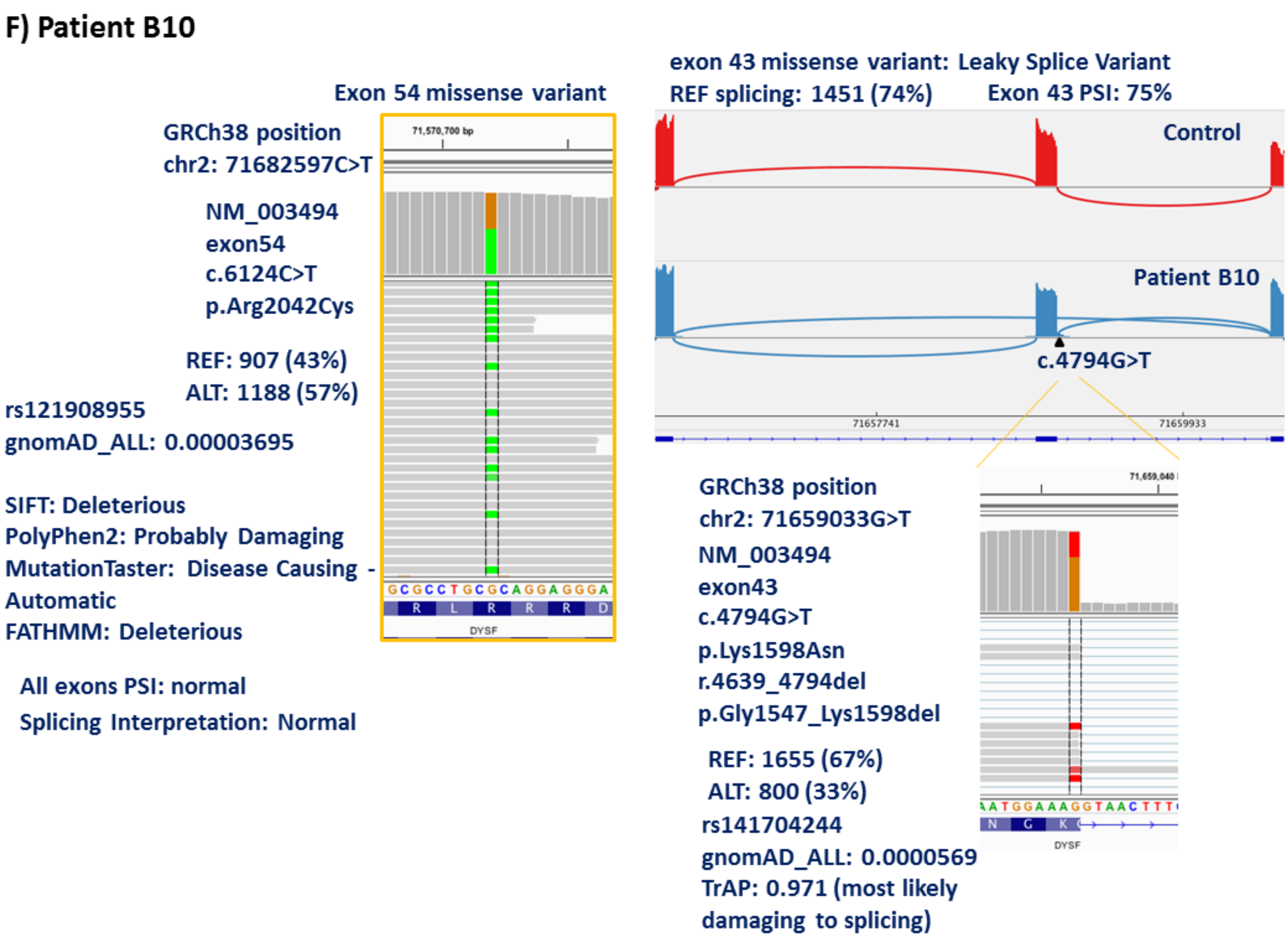

**(F)** Sashimi plots of examples (**Patients B10**; also see Figure. 3C in main text) of exon-skipping caused by a leaky splice variant in exon 43 (c.4794G>T) that was previously classified as a VUS and now we reclassified as pathogenic (right top panel). Right Inset: Variant expression in a minority of reads, which show a normal splice pattern. ~50% normal splicing without the exon 43 variant correlates well with the carrier range (patient B10: >23% DYSF protein) DYSF protein expression observed in this patient using the monocyte assay, and milder clinical symptoms. Missense exon 43 VUS, is reclassified as pathogenic (left) based on leaky splicing pathogenic effect. There are no other reportable variants. The exon 54 variant does not cause a splice defect or large AEI, but the ratio of

AEI is different than that for the exon 43 variant. The AEI ratio (43%:57%) seen in the case of the exon 54 variant is not exactly reciprocal to that of the other variant, since it is possible that some of the reads are captured before nonsense mediated decay or decay due to instability has occurred. Dysferlinopathy diagnosis with milder symptoms is confirmed.

#### G) Patient B12

##### Intron 9 extended splice variant

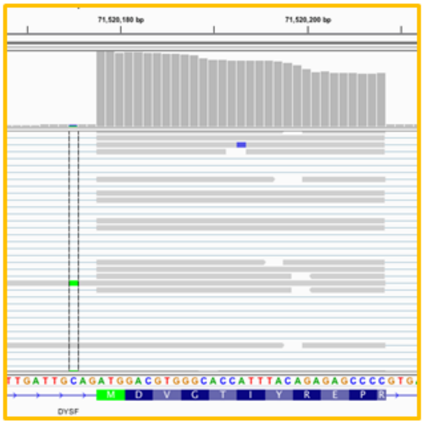

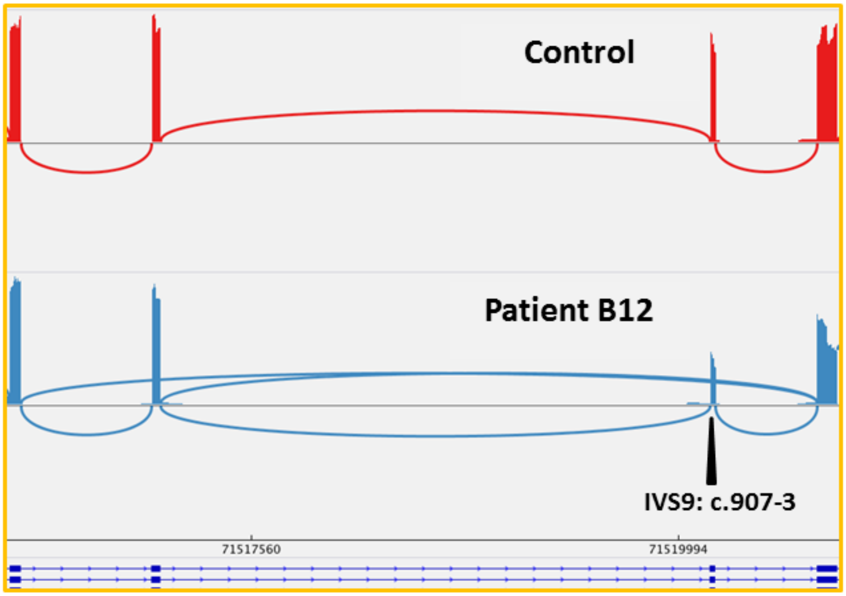

Exon 9 Exon 10

No rs#
gnomAD_ALL: no entry
TrAP: 0.475 (possibly damaging to splicing)
GRCh38 position
chr2: 71520175C>A
NM_003494
Intron9 c.907-3C>A

Splicing: exon skipping event
Exon 10 skip – r.907_937del; p.Met303Glyfs*25 (1091 reads, 49%)
Exon 9&10 skip –r.856_937del; p.Val286Glyfs*25 (223 reads, 10%)

##### Exon 34 frameshift variant

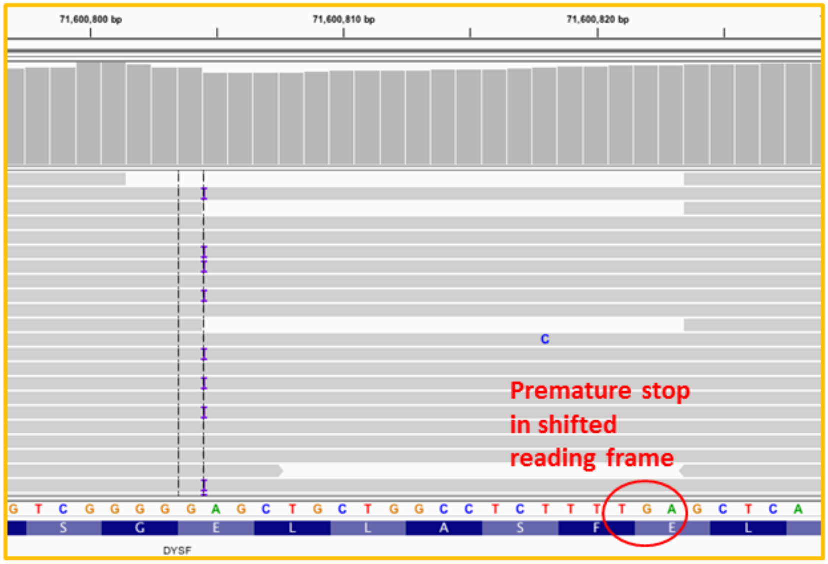

GRCh38 position
chr2: 71600804dupG

Splicing Interpretation: Normal
All exons PSI: normal

NM_003494
exon34
c.3805dupG
p.Glu1269Glyfs*7

REF: 1481 (69%)
ALT: 667 (31%)

rs779407815
gnomAD_ALL: 0.00000817

**(G) Patient B12:** Sashimi plot and IGV image showing that the IVS9, c.907-3C>A variant (top) causes two abnormal splicing events: complete skip of exon 10 in 49% of the transcripts causing a deletion at the RNA level, frameshift and a premature stop codon (r.907_937del; p.Met303Glyfs*25) and complete skip of exons 9 and 10 together in 10% of the transcripts causing a RNA deletion, frameshift, and premature stop codon (r.856_937del; p.Val286Glyfs*25). The IVS9 variant was previously classified as a VUS and now we reclassified as pathogenic. The second *DYSF* Exon 34 frameshift variant (bottom) is *in trans* to the IVS9 variant based on AEI ratios.

**H) Patient B14**
Intron 22 abnormal splice variant

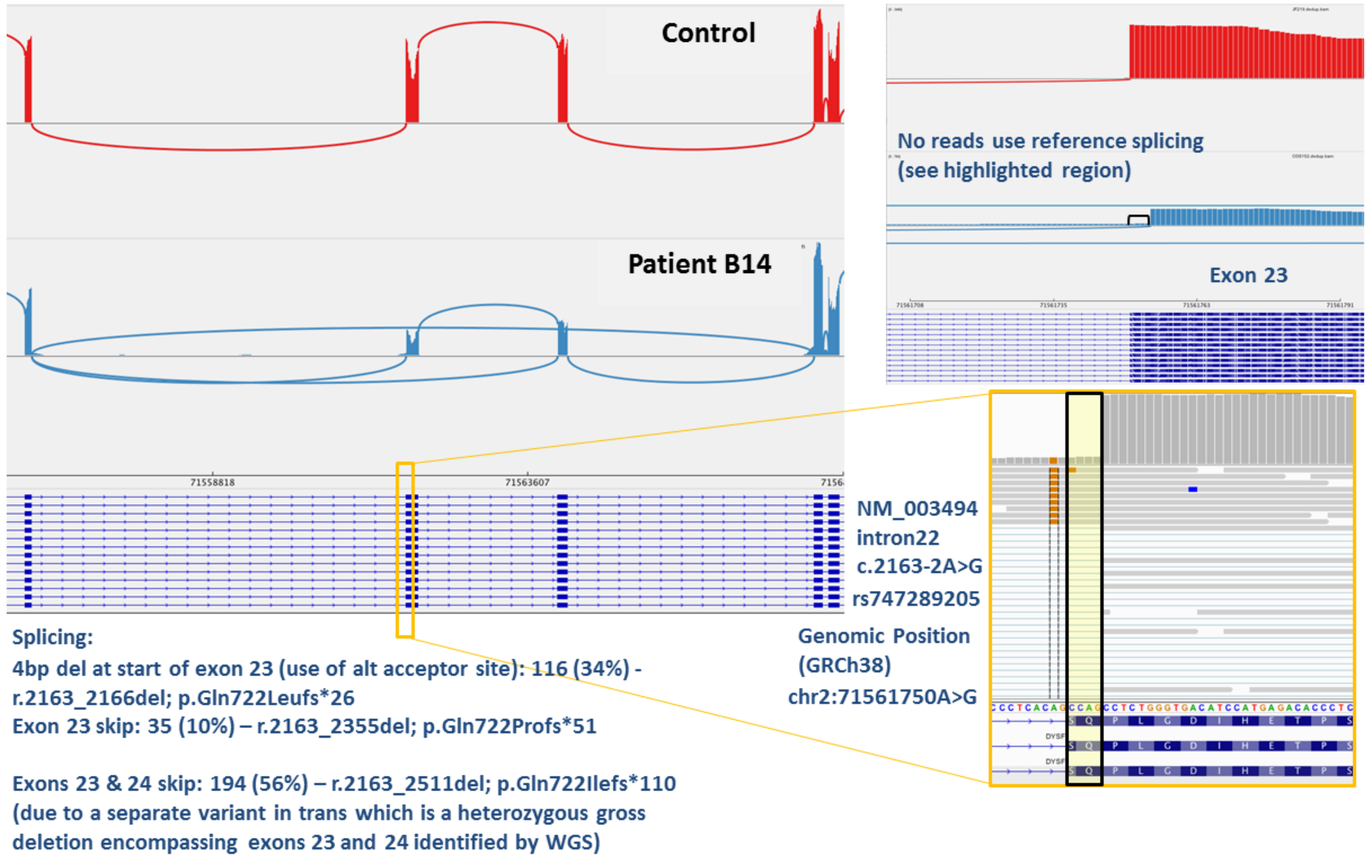

**(H) Patient B14:** IVS22: c.2163-2A>G previously had conflicting classifications (pathogenic/likely pathogenic). No other *DYSF* variant was identified in prior genetic testing in this patient. The c.2163-2A>G variant leads to an exonic splice gain (34%) and an exon 23 skip (10%) for a total of 44% abnormal reads, while the other allele contains a deletion of exons 23 and 24 (56% of reads). No normal spliced transcripts are seen (top right). Gene expression is significantly reduced. Dysferlinopathy diagnosis is confirmed. Subsequent WGS identified a gross heterozygous deletion encompassing exons 23 and 24 (c.2163_2511del) causing the exon 23 and exon 23-24 skip identified by RNA-Seq.

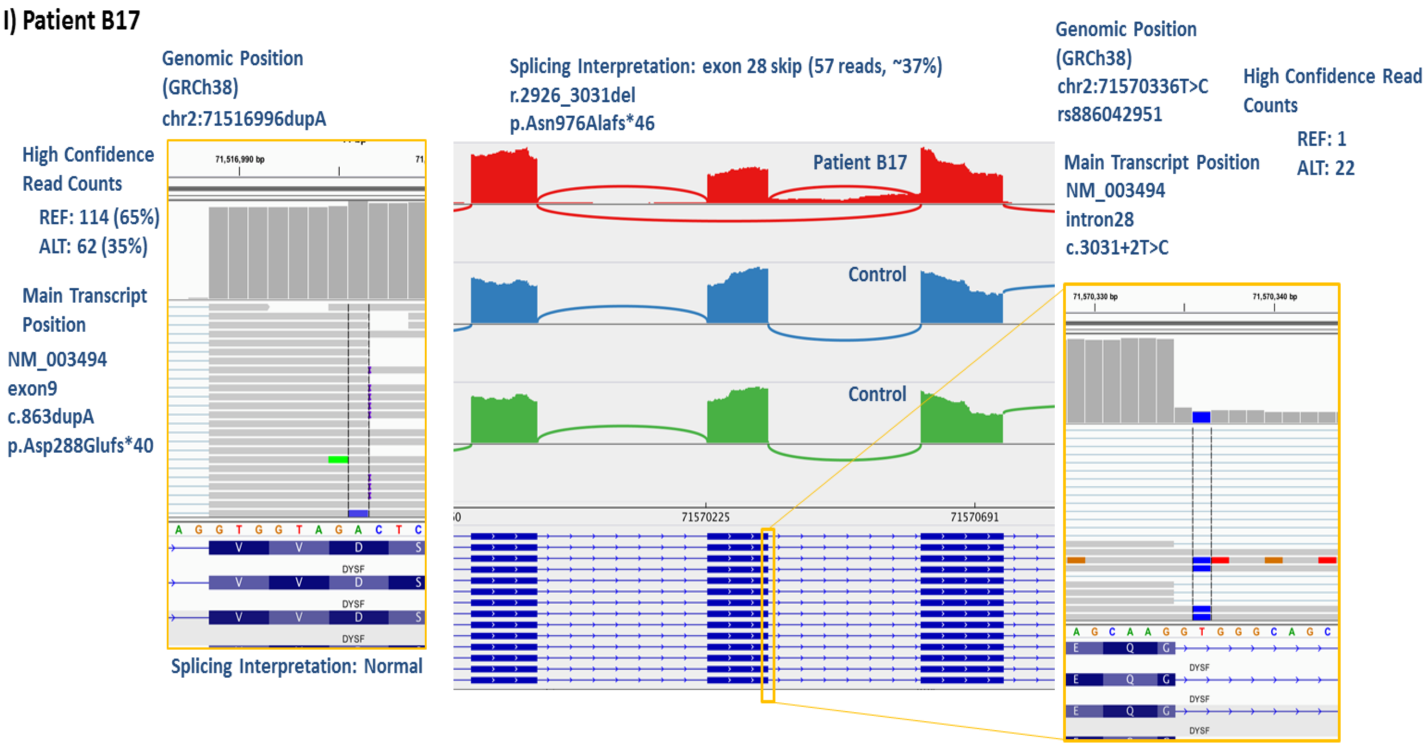

**(I) Patient B17:** Exon 9 frameshift variant was identified (left) and was the only *DYSF* variant found in prior genetic testing. RNA-Seq identified the *DYSF* IVS28: r.3031+2T>C (right inset) in unspliced reads. The c.3031+2T>C is known to be pathogenic. Sashimi plot (right) shows an exon 28 skip during splicing suggesting that the IVS28 pathogenic variant identified is the causal variant for the skip along with the pathogenic exon 9: c.863dupA variant. This explains "reduced" DYSF staining in muscle immunohistochemistry (IHC) / western blot (WB). Dysferlinopathy clinical diagnosis confirmed with two pathogenic variants with elucidation of pathogenic mechanism.

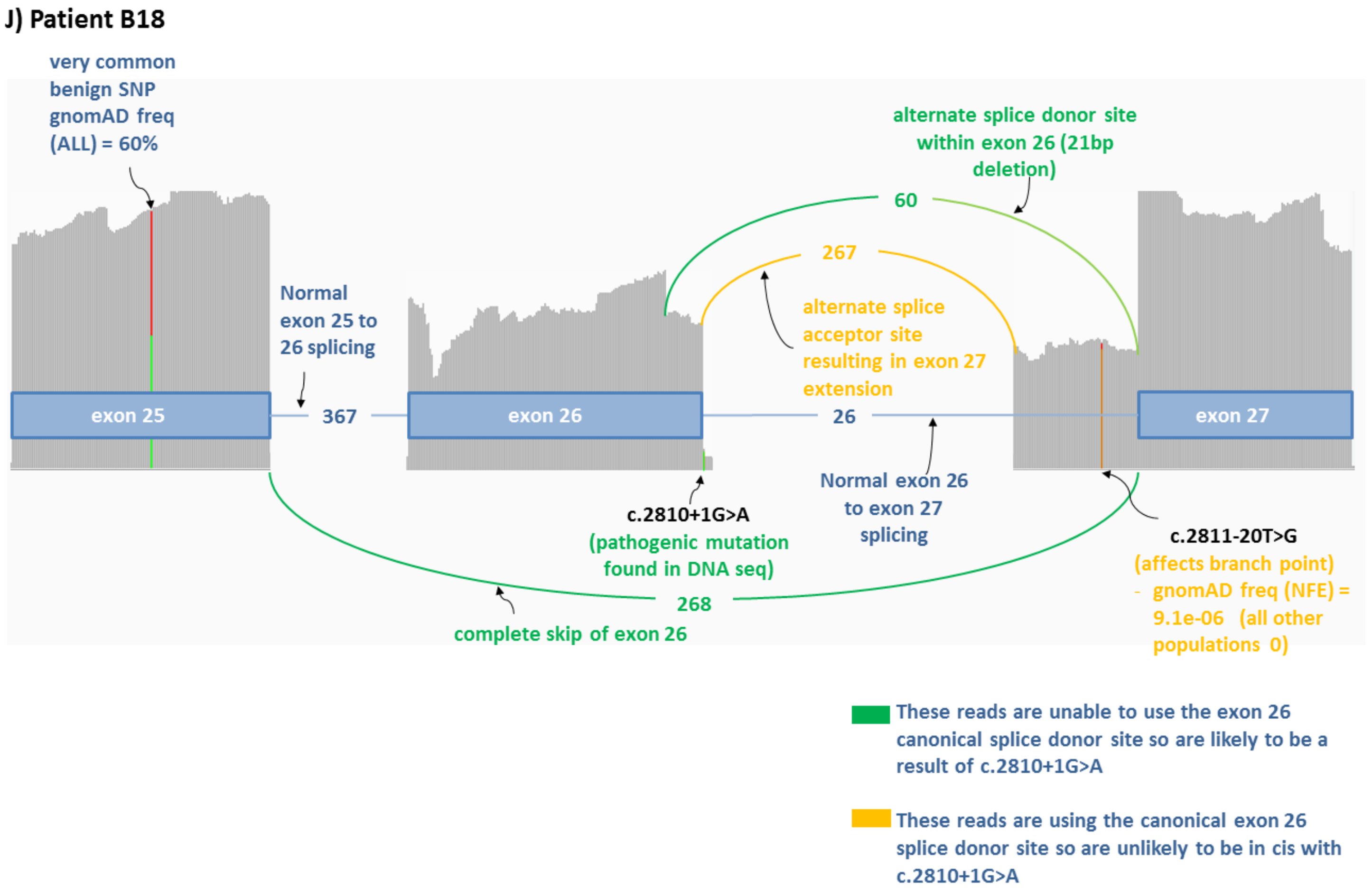

**(J) Patient B18** (also see Figure. 3B in main text): The previously done DNA sequencing only identified a single *DYSF* variant in IVS26 (c.2810+1G>A) which had conflicting interpretations of pathogenic/likely pathogenic. RNA-Seq identified exon 26 skipping caused by the destruction of the splice donor site by the c.2810+1G>A variant (arrow at exon 26-intron26 branchpoint) and exon 27 extension caused by the identification of a novel branch point variant (arrow pointing to the extended exon 27 region). The new variant identified is: IVS26: c.2811-20T>G which is classified as pathogenic. These two variants are found *in trans*. The IVS26 (c.2810+1G>A) variant causes a complete skip of exon 26 (4/5 aberrant reads) and a premature stop codon (r.2644_2810del; p.Tyr882Serfs*4), and the use of an alternate splice donor site in exon 26 (1/5 aberrant reads) resulting in an in-frame deletion (r.2790_2810del; p.Trp930_Thr937del) (green reads). The novel IVS26 (c.2811-20T>G) variant causes the use of an alternate splice acceptor site resulting in exon 27 extension and a frameshift deletion (r.2810_2811ins67; p.Leu938Argfs*3) (yellow reads), and thus is also classified as pathogenic.

**K) Patient B22**

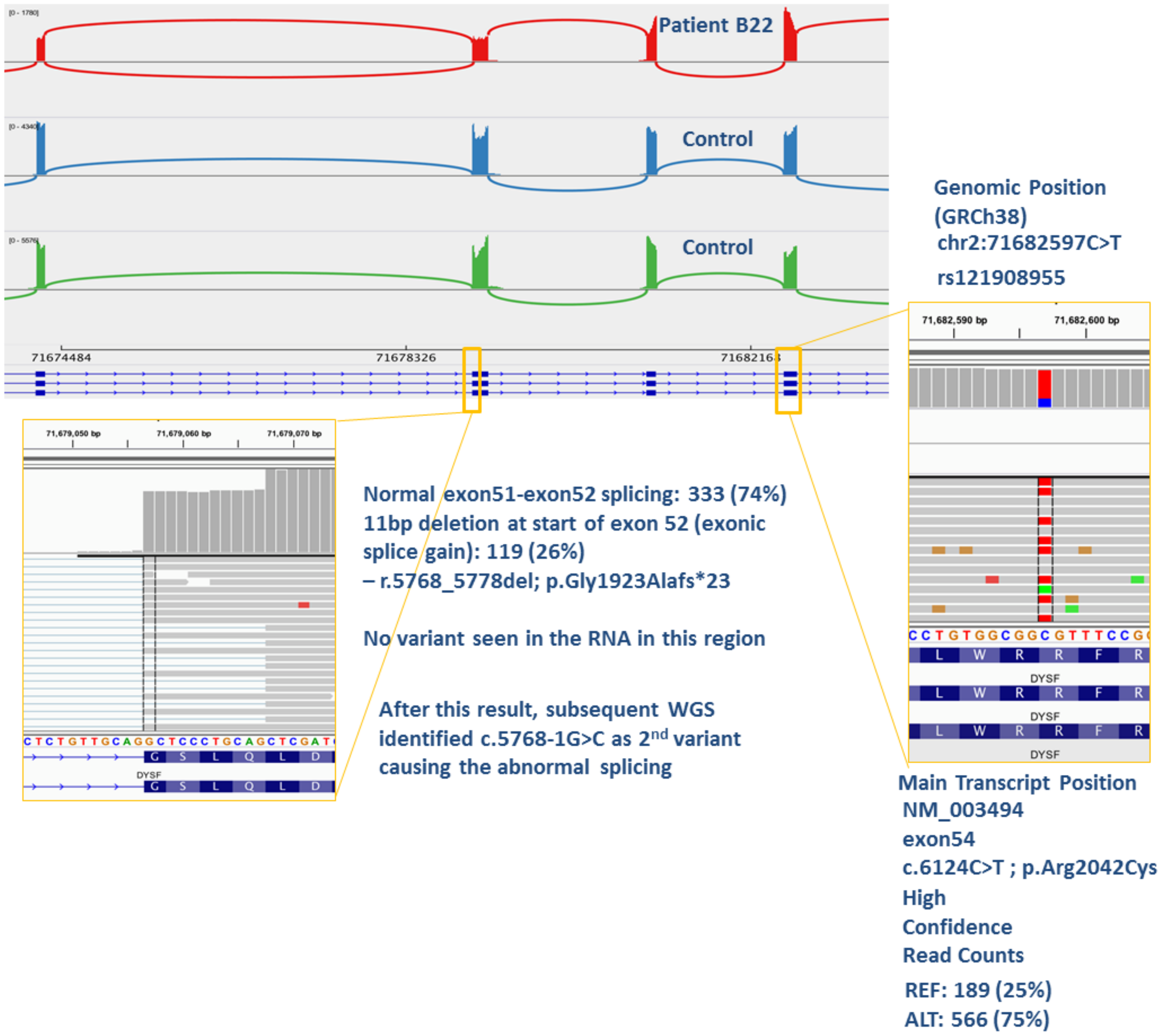

**(K) Patient B22:** In this patient, only the exon54 variant (c.6124C>T), which has conflicting interpretations [pathogenic (P) / likely pathogenic (LP)], was identified by prior DNA-level testing (right inset). RNA-seq identified a skip event (left inset) in exon52: r.5768_5778del causing p.Gly1923Alafs*23 by use of an alternate splice acceptor site causing an 11bp deletion at the start of exon 52 (26% ALT transcripts; 74% REF transcripts). Based on AEI ratios, the splicing event is *in trans* with exon54 variant: c.6124C>T. Subsequent WGS after RNAseq identified the 2nd genomic variant as being IVS51: c.5768-1G>C (previously LP, now we reclassified as P) that causes the abnormal exon 52 splicing. Dysferlinopathy diagnosis is confirmed.

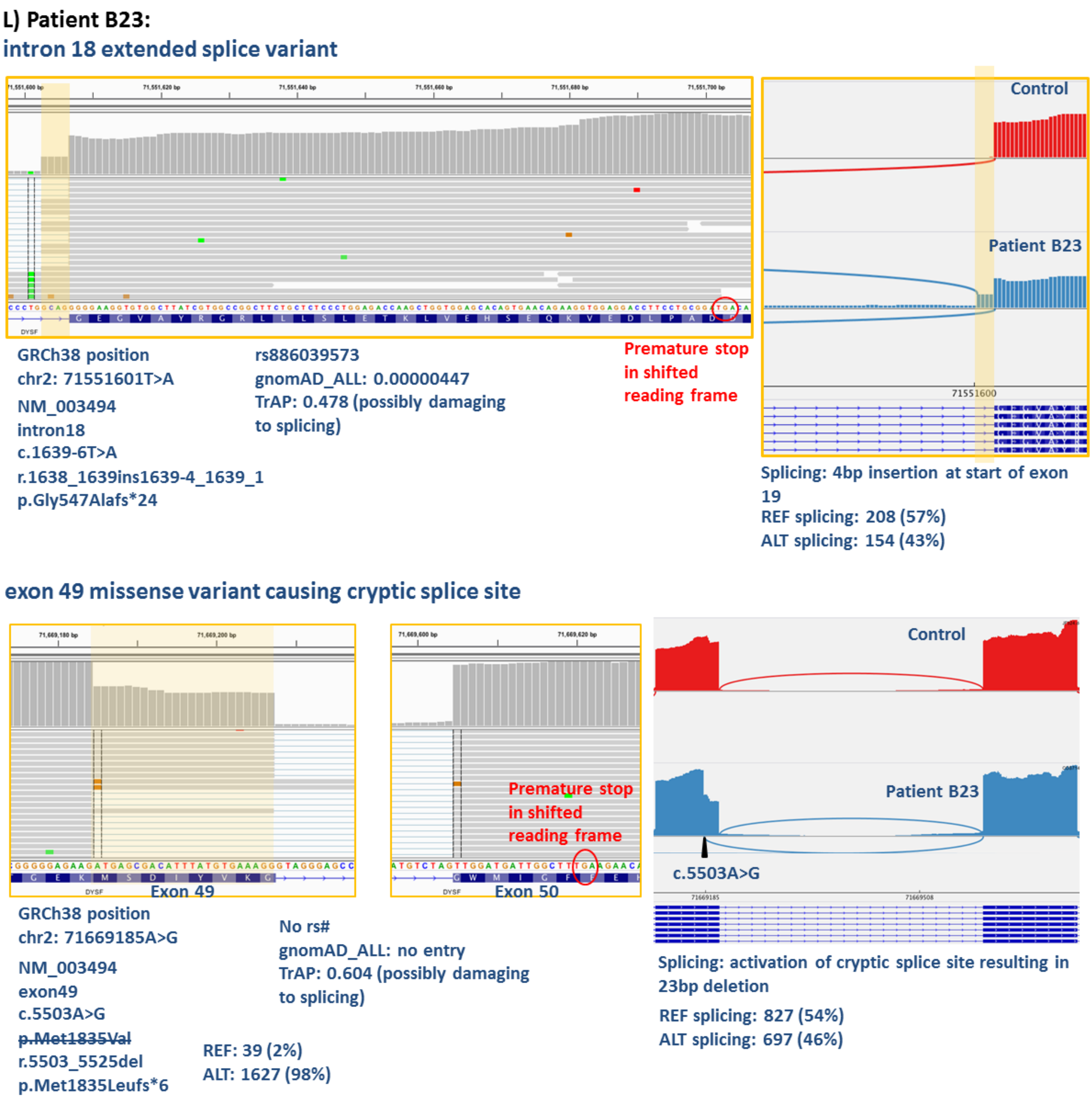

**(L) Patient B23** (also see Figure. 3D in main text): IVS18: c.1639-6T>A variant, which has conflicting classification (P/LP), was identified to create an alternate splice acceptor site in IVS18 causing a 4bp exon extension in the RNA (r.1638_1639ins1639-4_1639-1) that results in a frameshift and a premature stop codon (p.Gly547Alafs*24). The IVS18 variant is therefore reclassified to be a confirmed pathogenic variant. The Exon49: c.5503A>G variant, which was previously classified as a VUS, is identified to cause the activation of a cryptic splice donor site in exon 49 resulting in a 23bp deletion in the RNA (r.5503_5525del) that causes a frameshift and a premature stop codon (p.Met1835Leufs*6). Hence, the exon 49 VUS is now reclassified as pathogenic. *DYSF* mRNA expression is low, and absent DYSF expression in monocytes and muscle suggest the variants are *in trans*. Dysferlinopathy diagnosis is confirmed.

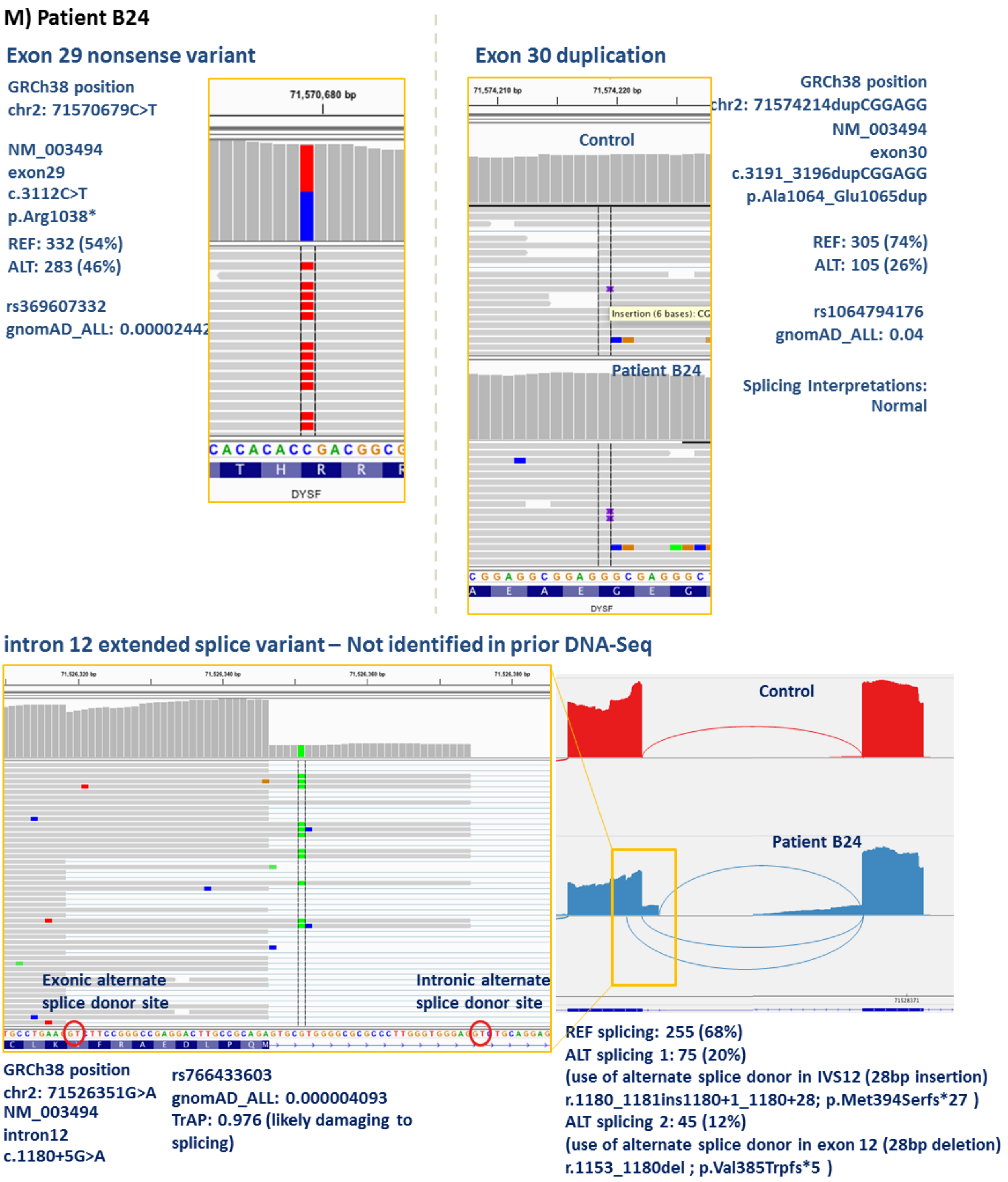

**(M)** **Patient B24:** This patient's prior genetic testing showed an exon 29 c.3112C>T nonsense pathogenic variant (top left), and an exon30: c.3191_3196dupCGGAGG (top right) non-frameshifting duplication VUS with conflicting interpretations (benign (B) /

likely benign (LB) / VUS). RNA-Seq, along with identifying these two variants, also identified a new IVS12: c.1180+5G>A variant which was a VUS with multiple conflicting interpretations (bottom inset). Sashimi plot and IGV shows that the IVS12 variant causes multiple splicing aberrations (bottom): use of alternate splice donor in IVS12 causing a 28bp insertion (r.1180_1181ins1180+1_1180+28) in 20% of the transcripts and results in a frameshift and premature stop codon (p.Met394Serfs*27); use of an alternate splice donor in exon 12 causing a 28bp deletion (r.1153_1180del) in 12% of the transcripts and results in a frameshift and premature stop codon (p.Val385Trpfs*5). The IVS12 variant is now reclassified as pathogenic. The AEI ratios of these splicing events due to the IVS12 pathogenic variant (REF: 68%, ALT:32%) is similar to that of the exon 30 VUS (REF:68%, ALT:32%), but both are different from that of the exon 29 pathogenic variant (REF 54%, ALT 46%). The allelic ratio (54%:46%) seen in the case of the exon 29 variant is not exactly reciprocal to that of the IVS12 variant (REF:ALT=68%:32%), since it is possible that some of the reads are captured before nonsense mediated decay has occurred in them. These findings suggest that the IVS12 and exon 30 variants are *in cis* to each other and both are *in trans* to the exon 29 pathogenic variant, and that the pathogenic variants causing disease are the IVS12 and exon 29 variants. Also based on this data, we have reclassified the exon 30 VUS as benign (B). *DYSF* mRNA expression is very low similar to two truncating variants. Taken together, Dysferlinopathy diagnosis is confirmed.

**N) Patient B25**

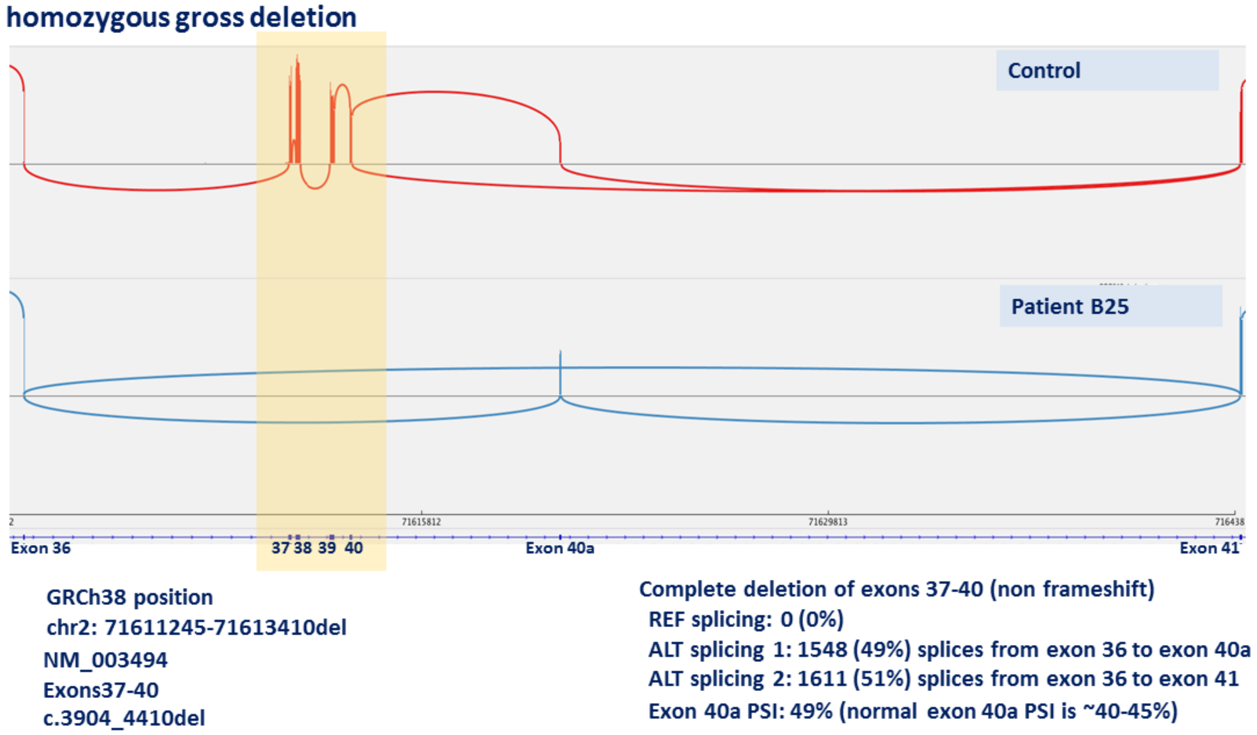

**(N) Patient B25:** RNA-seq identified a novel exon 37-40 gross in-frame deletion variant: c.3904-4410del which was previously classified as a VUS. Sashimi plot shows that the exon 37-40 deletion causes multiple abnormal splicing events. Alternate splicing occurring from exon 36 to exon 40a in 49% transcripts (exon 37-40 skip), and alternate splicing occurring from exon 36 to exon 41 in 51% transcripts (exon 37-40a skip) with no normal reference splicing occurring. The exons 37-40 gross in-frame deletion VUS is now reclassified as pathogenic. DYSF staining is absent in muscle IHC or western blot. Dysferlinopathy diagnosis is confirmed.

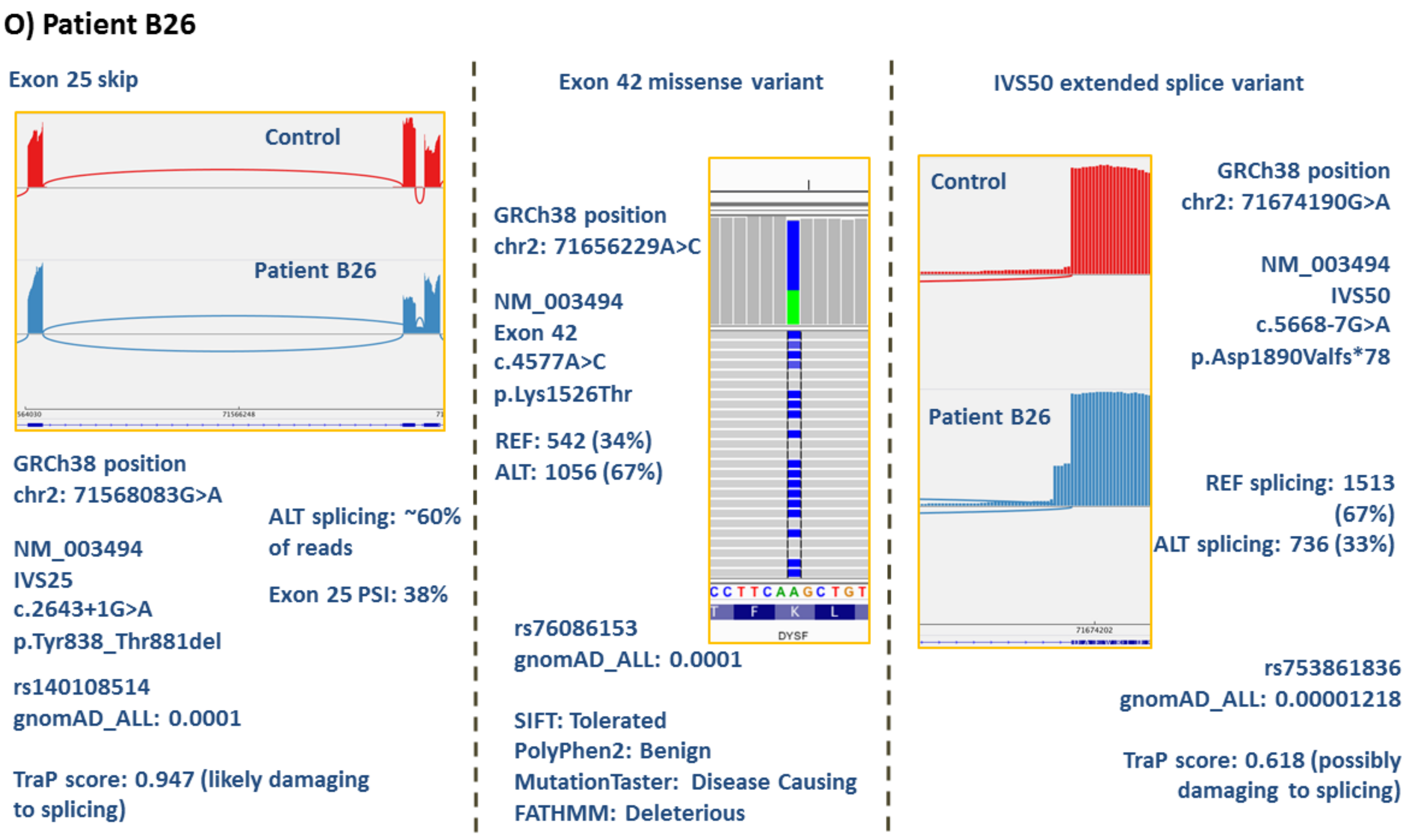

**(O)Patient B26:** IVS25, exon 42 and IVS50 variants were all identified by prior DNA testing, but it was uncertain which variant combination was pathogenic and VUSs or variants with conflicting interpretation needed to be reclassified for that purpose. RNA-Seq identified the IVS25 variant (c.2643+1G>A) (left) which had conflicting interpretations (P/LP). RNA-Seq identified the skipping of exon 25 in 60% of the transcripts due to the IVS25 variant, which causes a non-frameshifting deletion (p.Tyr838_Thr881del). This allows the reclassification of this variant to pathogenic (AEI: REF 40%, ALT 60%). RNA-Seq identified the exon 42 missense VUS (c.4577A>C; middle) with AEI: REF 34%, ALT 67%. RNA-Seq also identified the IVS50 LP variant (c.5668-7G>A; right) with AEI: REF 67%, ALT 33%. The IVS50 variant was found to cause an extension of exon 51 resulting in a frameshift and premature stop codon (p.Asp1890Valfs*78). Therefore, the pathogenic mechanism of IVS50 variant is now elucidated and the IV50 variant can now be classified as pathogenic. RNAseq shows that the pathogenic variants i.e., IVS25 and IVS50 variants are *in trans* based on AEI ratio comparison. *DYSF* mRNA expression is in the low range which fits with what we have seen previously with two truncating variants. The Exon 42 missense VUS is *in cis* with IVS25 variant based on AEI ratios and not pathogenic; hence we have reclassified the exon 42 variant as Benign. Dysferlinopathy diagnosis is confirmed.

**P) Patient B27**

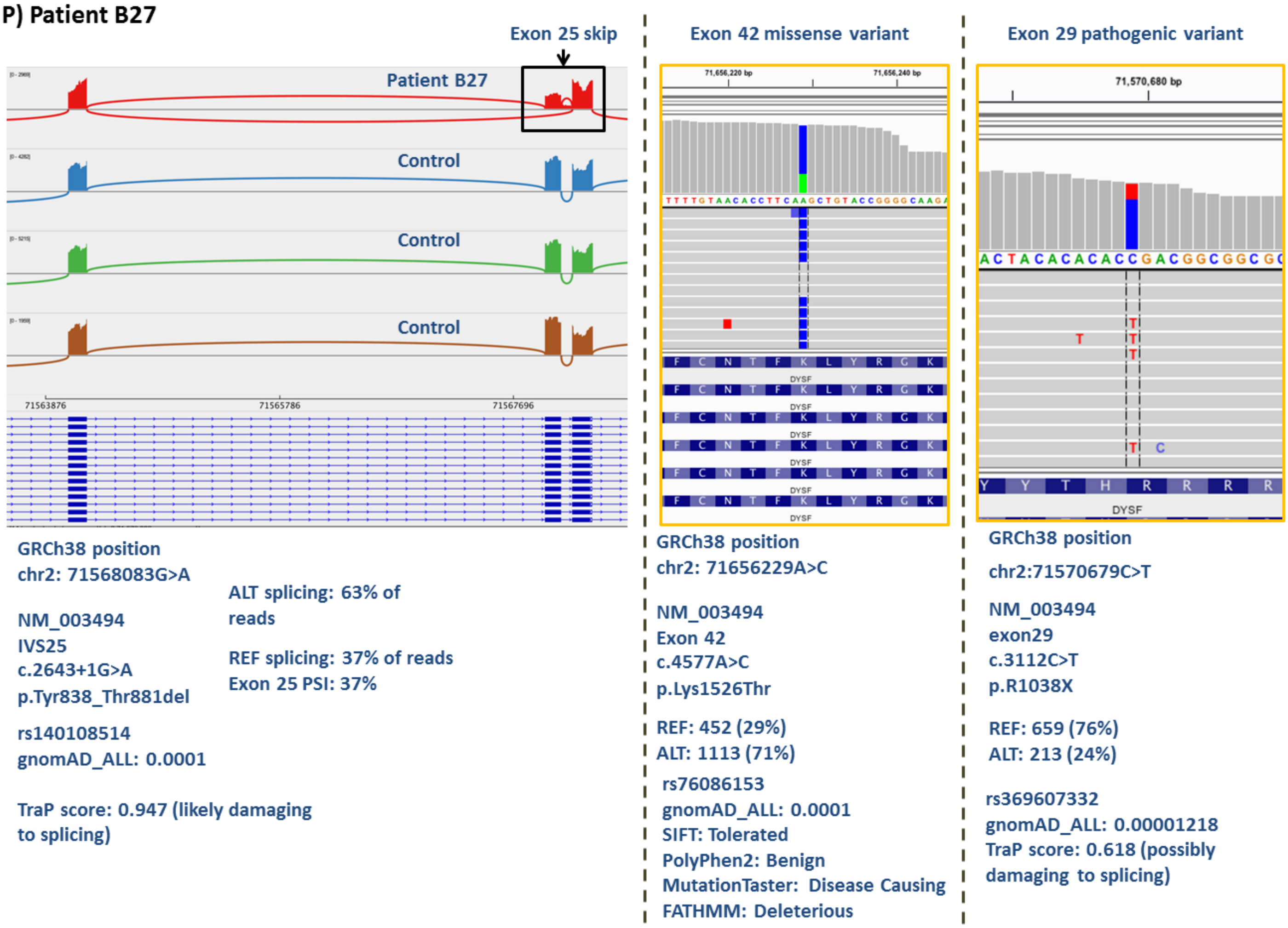

**(P) Patient B27:** IVS25, exon 29, and exon 42 variants were all identified by prior DNA testing, but it was uncertain which variant combination was pathogenic and VUSs or variants with conflicting interpretation needed to be reclassified for that purpose. RNA-

Seq identified the IVS25 variant (c.2643+1G>A) (left) which had conflicting interpretations (P/LP). RNA-Seq identified the skipping of exon 25 in 63% of the transcripts due to the IVS25 variant, which results in a non-frameshift deletion (p.Tyr838_Thr881del). This data allows the IVS25 variant to be reclassified as pathogenic (AEI: REF 37%, ALT 63%). RNA-Seq identified the exon 42 missense VUS (c.4577A>C; middle) with AEI: REF 29%, ALT 71%. RNA-Seq also identified the exon 29 nonsense pathogenic variant (c.3112C>T; right) with AEI: REF 76%, ALT 24%. RNAseq shows that the pathogenic variants i.e., IVS25 and exon 29 variants are *in trans* based on AEI ratio comparison and are pathogenic. *DYSF* mRNA expression is in the low range, which fits with what we have seen previously with two truncating variants. The Exon 42 missense VUS is *in cis* with the IVS25 variant based on AEI ratios and is not pathogenic; hence we have reclassified the exon 42 variant as Benign. Dysferlinopathy diagnosis is confirmed.

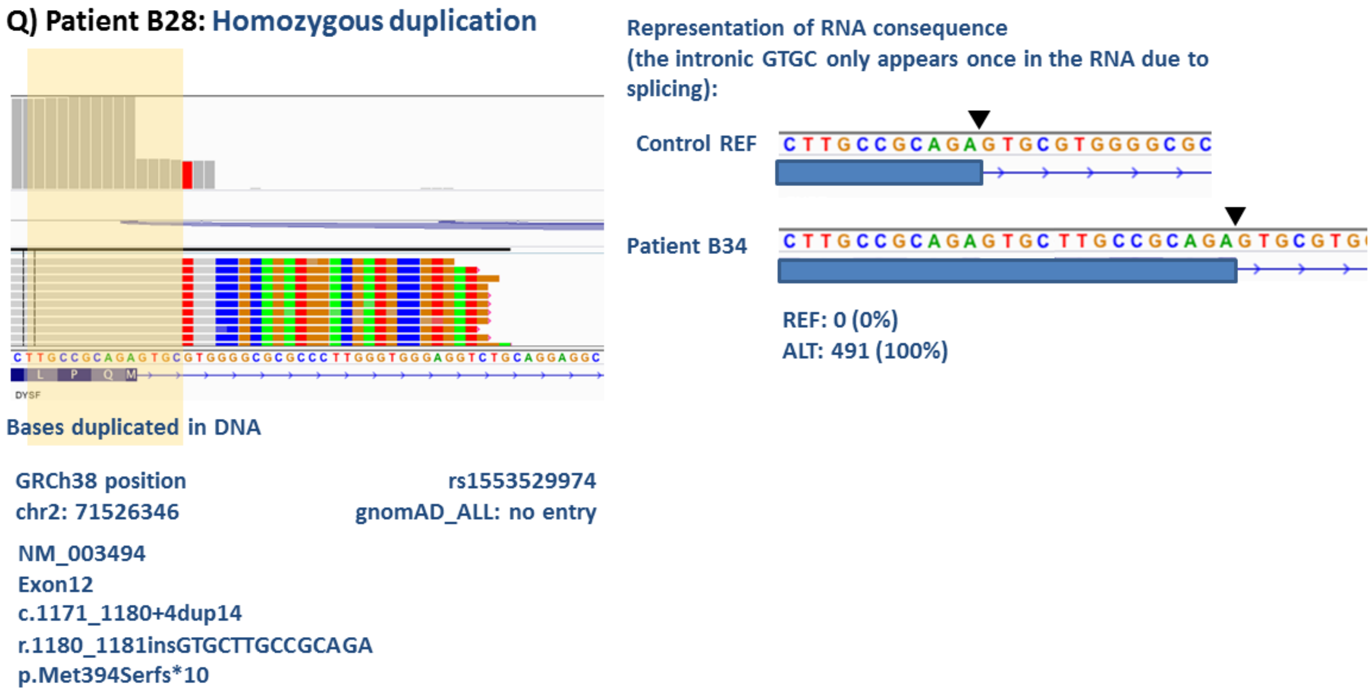

**(Q)Patient B28:** The Exon 12 duplication variant in a homozygous state was found in prior DNA testing with no other *DYSF* reportable variants. The Exon12 duplication variant c.1171_1180+4dup14 was classified as a VUS. IGV image (left) and a blowup representation (right) show that the duplication VUS causes an insertion and subsequent splicing defect by extending exon 12 (r.1180_1181insGTGCTTGCCGCAGA) resulting in a downstream frameshift and a premature stop codon (p.Met394Serfs*10). Subsequent Neuromuscular Disease gene-panel sequencing and deletion/duplication testing confirmed the RNAseq result by identifying the IVS12 variant resulting in the duplication of fourteen nucleotides at positions c.1171 through c.1180 of the *DYSF* gene after the end of exon 12. This VUS is now reclassified as pathogenic. Dysferlinopathy diagnosis is confirmed.

**Figure S3 (A-B). Sashimi and/or IGV plots of cases with *DYSF* splicing events identified that are related to two pathogenic events or reclassification of VUSs or discovery of two pathogenic mechanisms that completes the molecular diagnosis (also see Table S5 for reference and details of each patient case)**

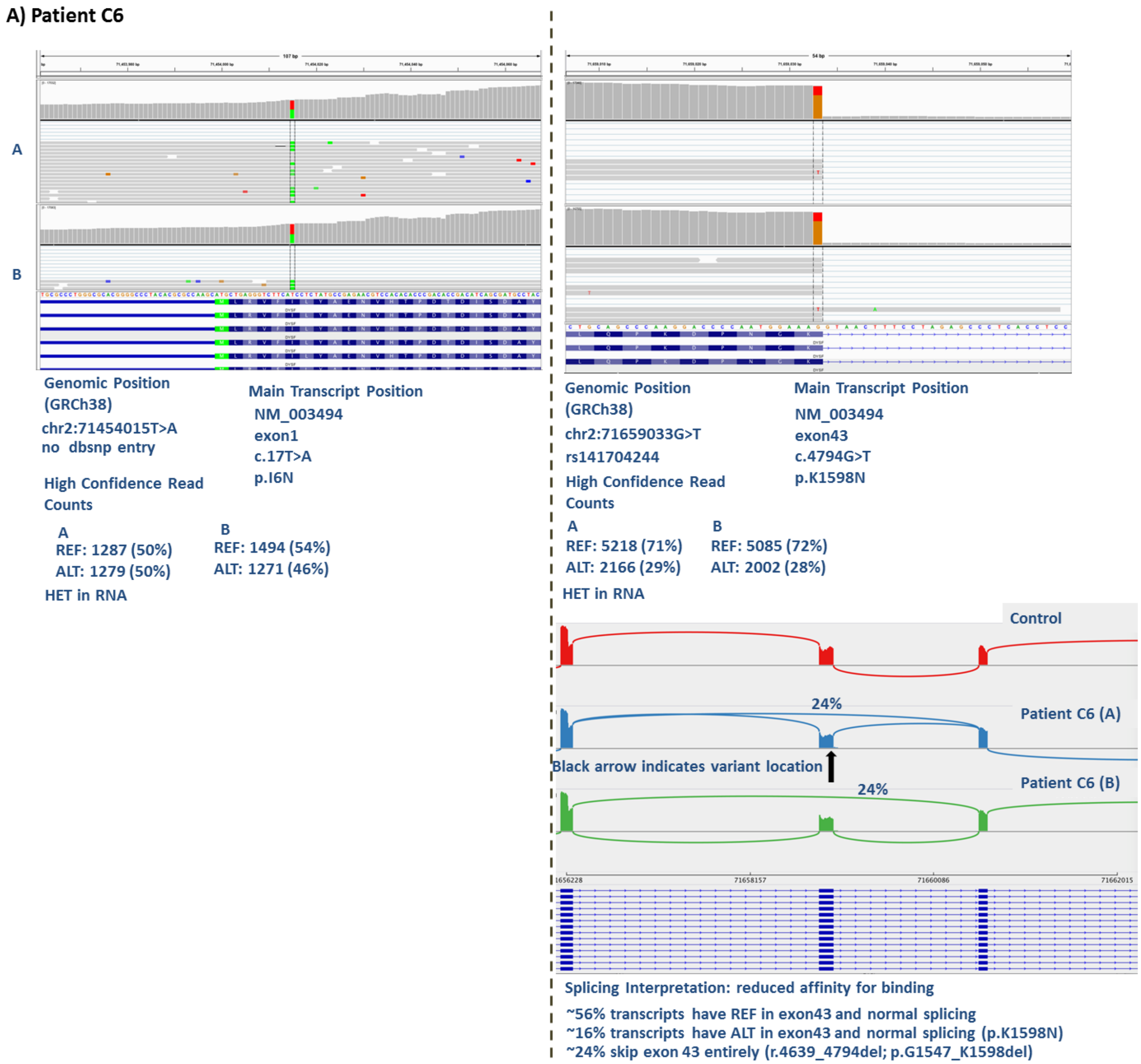

**(A) Patient C6** (also see Figure. 3C in main text): Both exon 1 and exon 43 VUSs were found in prior DNA testing and needed to be reclassified. Exon 1 VUS (c.17T>A)

remains a VUS as RNA-Seq did not identify any RNA-level abnormality. RNA-seq identified the exon 43 variant as a leaky splice variant resulting in 24% of the transcripts skipping exon 43 and 16% of the transcripts harboring the variant but do not skip exon 43 during splicing. Thus, the exon 43 VUS is now reclassified as pathogenic. 56% normal splicing without the exon 43 variant correlates well with carrier range 43% DYSF protein expression in monocyte assay, and milder clinical symptoms. Exon 43 leaky splice variant, carrier range DYSF protein expression, and clinical features together provide genotype-phenotype correlation and patient stratification. Dysferlinopathy diagnosis with milder symptoms is confirmed.

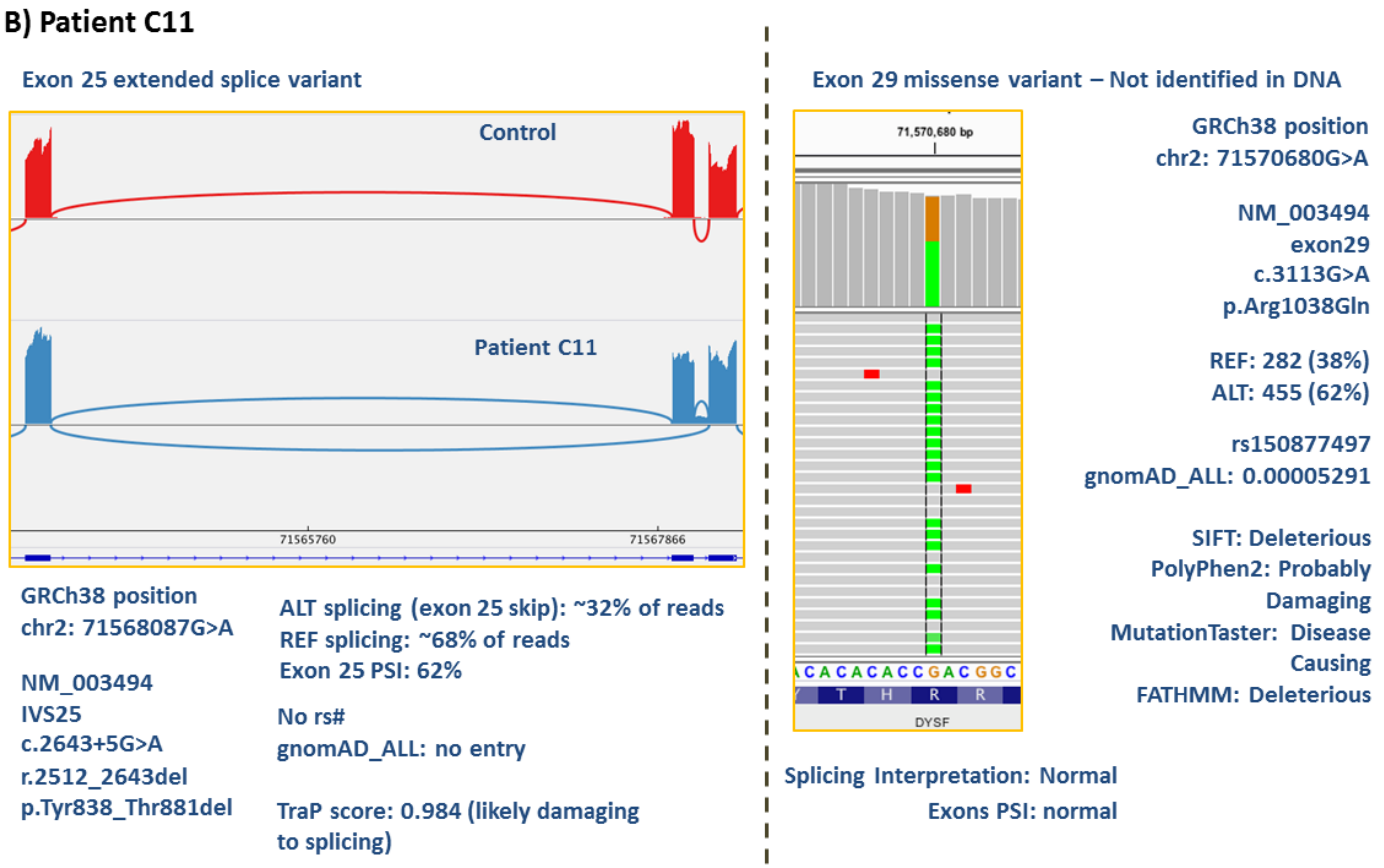

**(B) Patient C11:** Only a novel IVS25 VUS (c.2643+5G>A) was found in prior DNA testing in this patient. RNA-Seq identified that the IVS25 VUS causes skipping of exon 25 in

32% of the transcripts causing a non-frameshifting deletion (p.Tyr838_Thr881del) and hence this variant can be reclassified as pathogenic. For the IVS25 variant, the splicing AEI ratio is: REF 62%; ALT ~32%. RNA-seq identified a second variant in exon 29 (c.3113G>A) which was classified as pathogenic previously. The exon 29 pathogenic variant AEI is: REF 38%, ALT 62%, suggesting the two variants are *in trans*. %DYSF is 0% in monocytes and the patient is clinically suspected of dysferlinopathy. Dysferlinopathy diagnosis is confirmed.

**Figure S4. IGV plots of patient C12 showing identification of a third new *DYSF* variant not previously reported in genetic testing, *in trans* based on AEI, confirming molecular diagnosis (also see Table S5 for reference and details of patient case)**

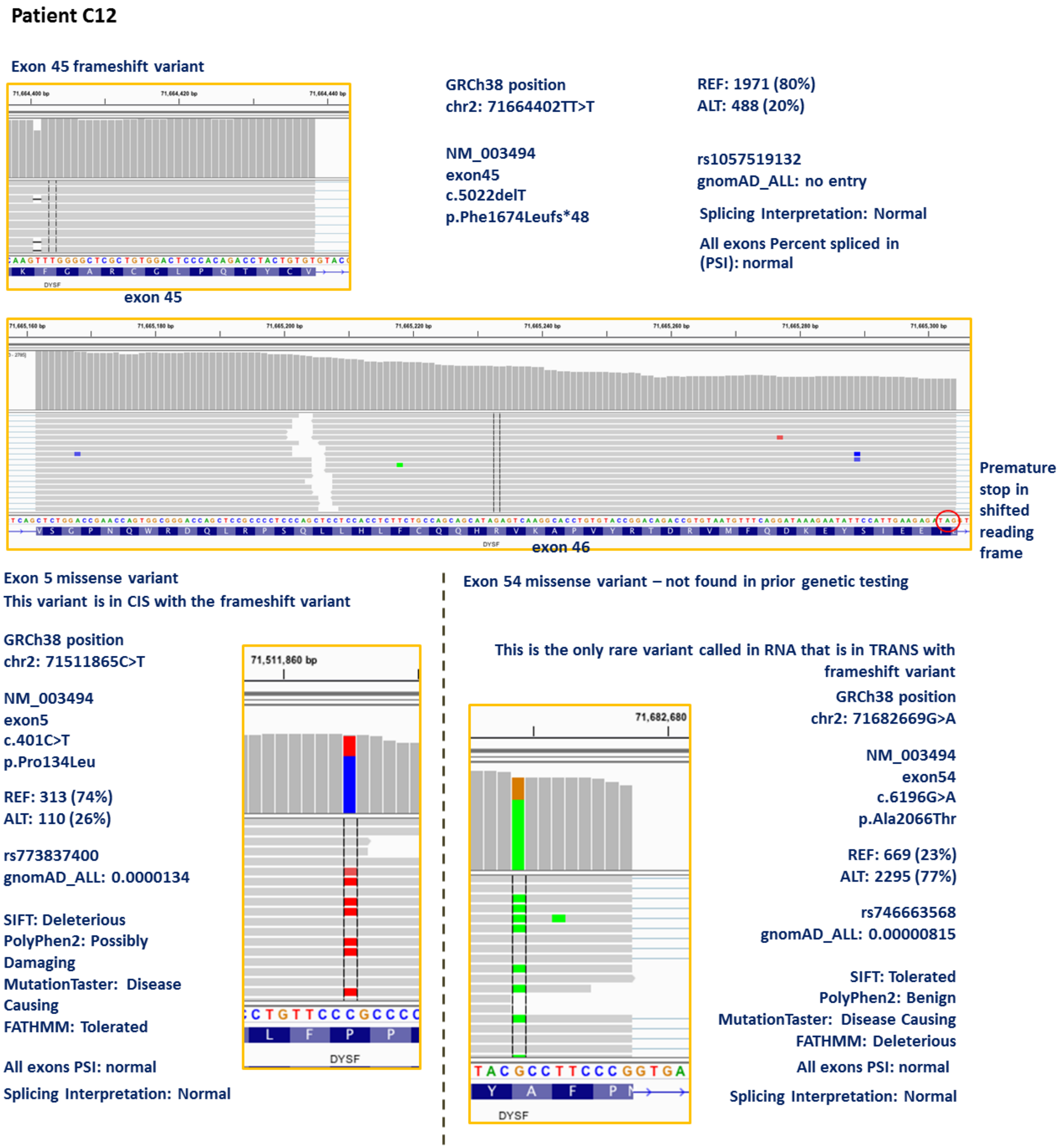

**Figure S4. Patient C12:** This patient's prior DNA-testing only found the exon 45 variant (c.5022delT) which had been previously classified with conflicting interpretations (P/LP), and the exon 5 VUS (c.401C>T) which also had conflicting interpretations (LP/VUS). RNA-Seq identified that the exon 45 variant causes a frameshift and premature stop codon (top and middle) with AEI ratio: REF 80%, ALT 20%, which allows this variant to be reclassified as pathogenic. RNA-Seq identified the exon 5 missense VUS (bottom left) with AEI ratio: REF 74%, ALT 26%; suggesting it is *in cis* with exon 45 pathogenic variant c.5022delT, and thus the exon 5 variant is not pathogenic. RNA-Seq also identified a third new variant that was not reported by the prior DNA testing (bottom right): exon 54 missense VUS (c.6196G>A) with an AEI ratio: REF 23%, ALT 77% suggesting it is *in trans* with the exon 45 frameshift variant c.5022delT and DYSF protein expression is significantly reduced in muscle seen by immunohistochemistry and immunoblotting, and is therefore pathogenic. Thus, the exon 5 missense VUS is now reclassified as benign (B). Dysferlinopathy diagnosis is confirmed.

18. Richards S, Aziz N, Bale S, et al. Standards and guidelines for the interpretation of sequence variants: a joint consensus recommendation of the American College of Medical Genetics and Genomics and the Association for Molecular Pathology. *Genetics*

*in medicine : official journal of the American College of Medical Genetics.* 2015;17(5):405-424.

19. Lek M, Karczewski KJ, Minikel EV, et al. Analysis of protein-coding genetic variation in 60,706 humans. *Nature.* 2016;536(7616):285-291.

20. Robinson JT, Thorvaldsdottir H, Winckler W, et al. Integrative genomics viewer. *Nat Biotech.* 2011;29(1):24-26.

21. Adzhubei I, Jordan DM, Sunyaev SR. Predicting functional effect of human missense mutations using PolyPhen-2. *Curr Protoc Hum Genet.* 2013;Chapter 7:Unit7 20.
